## Supplementary Material for "Comparative analyses of FDA EUA-approved rapid antigen tests and RT-PCR for COVID-19 quarantine and surveillance-based isolation"

**Supplementary methods** 4

Quarantine and testing 4

Frequency of testing to reduce the effective reproduction number 5

Fitting the percent positive agreement 6

False-negative for serial testing strategies 8

Diagnostic sensitivity of the RT-PCR assay 9

Estimation of rapid antigen percent positive agreement with RT-PCR during quarantine. 12

Estimation of offshore prevalence of SARS CoV-2 14

Comparison of the internal and external percent positive agreement data sets 18

**Supplementary tables** 19

**Supplementary figures** 36

### Supplementary methods

*Quarantine and testing*

During the disease time course, the infectivity can be characterized by a functional form

$$r(t) = \begin{cases} R_0 f(t), & t \leq t_E, \\ 0, & t > t_E, \end{cases} ,$$

where $f(t)$ is the relative infectivity over the course of the disease, and $t_E$ is the duration of the disease. The average number of secondary infections in the absence of self-isolation or other interventions is then

$$R_0 = \int_0^\infty r(t)\, \mathrm{d}t.$$

A proportion of infections $p_A$ are asymptomatic. The remainder, $1 - p_A$, will develop symptoms $t_S$ days after infection (i.e. the incubation period). For an asymptomatic individual, we assume the infectivity over time is $r_A(t) = r(t)$. Once an individual exhibits symptoms, they enter isolation and are assumed to no longer transmit the disease. To model the isolation of symptomatic individuals upon onset, we computed their infectivity over time as

$$r_S(t) = \begin{cases} r(t) & 0 \leq t \leq t_s, \\ 0 & t > t_s \end{cases} .$$

Individuals who manifest symptoms are not eligible for quarantine, and instead are isolated.For individuals who enter quarantine randomly over the period in which they do not exhibit symptoms, the expected post-quarantine transmission is

$$R_{q^{\mapsto}}(q) = (1 - p_A) R_{S,q^{\mapsto}}(q) + p_A R_{A,q^{\mapsto}}(q). \quad \text{(S1)}$$

The diagnostic sensitivity $s(t)$ of a test depends on the time of testing post-infection. For a specified duration of quarantine $q$, number of tests administered $N$, delay in obtaining test result $d_t$, and time of testing occurring over the course of quarantine $t_i$, the expected post-quarantine number of secondary infections for a soon-to-be symptomatic case tested for disease at any time $0 \leq t_n \leq q - d_t$ is

$$R_{S,q^{\mapsto}}(q) = \frac{1}{t_s} \int_{u=0}^{t_s} \int_{t=q}^{\infty} r_S(t + u) \cdot \prod_{n=1}^{N} \left(1 - s\left(t_n + u\right)\right) \mathrm{d}t\, \mathrm{d}u. \tag{S2}$$

For asymptomatic carriers,

$$R_{A,q^{\mapsto}}(q) = \frac{1}{t_e} \int_{u=0}^{t_e} \int_{t=q}^{\infty} r(t + u) \cdot \prod_{n=1}^{N} \left(1 - s\left(t_n + u\right)\right) \mathrm{d}t\, \mathrm{d}u. \tag{S3}$$

*Frequency of testing to reduce the effective reproduction number*

The effective reproduction number when conducting serial testing and isolation of positives every θ days is

$$R(\theta) = (1 - p_A)R_S(\theta) + p_A R_A(\theta). \tag{S4}$$

As individuals are not within quarantine during their surveillance, there is potential for an individual to be infected and transmit between tests. Assuming a uniform risk of infection, the probability of being infected between tests is 1/θ. For a soon-to-be symptomatic individual who will isolate upon symptom onset, the effective reproduction number with testing every θ days is

$$R_S(\theta) = \frac{1}{\theta} \int_{\varphi=0}^{\theta} \left( \int_{t=0}^{\varphi} r_S(t)\, \mathrm{d}t + \sum_{d=0}^{N_T(\theta)-2} \left( \prod_{\tau=0}^{d} (1 - s(\varphi + (\theta \cdot \tau))) \cdot \int_{t=\varphi+\theta d}^{\varphi+\theta(d+1)} r_S(t) \ )\, \mathrm{d}t \right) \right) \mathrm{d}\varphi,$$

$$+ \frac{1}{\theta} \int_{\varphi=0}^{\theta} \left( \prod_{\tau=0}^{N_T(\theta)-1} (1 - s(\varphi + (\theta \cdot \tau))) \cdot \int_{t=\varphi+\theta(N_T(\theta)-1)}^{n} r_S(t)\mathrm{d}t \right) \mathrm{d}\varphi, \tag{S5}$$

where φ denotes the offset from infection to the next test (e.g., if $\varphi = 2$, then the initial test is conducted two-days post-infection), and $N_T(\theta)$ denotes the number of tests conducted over the duration of disease for the specified testing frequency.

For an asymptomatic individual, the effective reproduction number with testing every θ days and isolation of positive cases is

$$R_A(\theta) = \frac{1}{\theta}\int_{\varphi=0}^{\theta}\left(\int_{t=0}^{\varphi} r_A(t)\,\mathrm{d}t + \sum_{d=0}^{N_T(\theta)-2}\left(\prod_{\tau=0}^{d}(1 - s(\varphi + (\theta\cdot\tau)))\cdot\int_{t=\varphi+\theta d}^{\varphi+\theta(d+1)} r_A(t)\ \ )\,\mathrm{d}t\right)\right)\mathrm{d}\varphi.$$

$$+\frac{1}{\theta}\int_{\varphi=0}^{\theta}\left(\prod_{\tau=0}^{N_T(\theta)-1}(1 - s(\varphi + (\theta\cdot\tau)))\cdot\int_{t=\varphi+\theta(N_T(\theta)-1)}^{n} r_A(t)\mathrm{d}t\right)\mathrm{d}\varphi. \quad \text{(S6)}$$

*Fitting the percent positive agreement*

We estimated the coefficients of a linear logistic model

$$\ln\left(\frac{p(t)}{1-p(t)}\right) = \beta_0 + \beta_1(t - t_S) \quad \text{(S7)}$$

for the percent positive agreement $p(t)$ between each rapid antigen (RA) test and the RT-PCR result at time $t$ post infection. For each RA test, we fit the model to the data using a log-likelihood function $L$. Data on percent positive agreement for many RA tests were available from the day after symptom onset to 5–14 days after symptom onset, depending on the test. For some RA tests, the percent positive agreement with RT-PCR was calculated by aggregating results over a time span longer than a single day (e.g., 8–10 days after symptom onset). Similarly, the percent positive agreement with RT-PCR for some RA tests was specified for samples only identified as exceeding a threshold time after symptom onset (e.g., six days after symptom onset). We accounted for this heterogeneity in the format of data reporting by constructing a compound log-likelihood function

$$L = L_D + L_S + L_C, \quad \text{(S8)}$$

where the log-likelihood is for the number of positive RA tests given the number of positive RT-PCR tests when reported as occurring on a specific day after symptom onset is $L_D$, when reported as occurring at some point during a specified time span is $L_S$, and when reported as occurring on a day of or subsequent to a threshold day after symptom onset is $L_C$. If a set of percent positive agreement data (e.g., specific day, time span, or threshold) was absent for a RA test, then the corresponding log-likelihood term was set to zero.

The log-likelihood for the number of positive results that were obtained from the RA tests given the number of positive RT-PCR tests on a specified day after symptom onset is

$$L_D = \sum_{i=1}^{N} \log\left(p(t_i + t_S)^{S_i}(1 - p(t_i + t_S))^{F_i}\right), \qquad \text{(S9)}$$

where $S_i$ is the number of successes at time point $t_i$ since symptom onset (i.e., number of infected individuals testing positive for both the RA and the RT-PCR tests at time $t_i$ since symptom onset), $F_i$ is the number of failures (i.e., number of infected individuals testing negative for the RA test and positive for RT-PCR test at time $t_i$ since symptom onset), and $N$ is the number of time points.

The log-likelihood for the number of positive RA tests given the number of positive RT-PCR tests for a specified time span of days after symptom onset is

$$L_S = \sum_{j=1}^{N_S}\left(\frac{1}{1+\tau_j - t_j}\sum_{k=t_j}^{\tau_j} \log\left(p(k + t_S)^{S_j}(1 - p(k + t_S))^{F_j}\right)\right), \qquad \text{(S10)}$$

where $S_j$ is the number of successes within time span $j$ , $F_j$ is the number of failures, and $N_S$ is the number of time spans in the dataset, $t_j$ is the initial time point in the time span, and $\tau_j$ is the final time point in the time span.

The log-likelihood for the number of positive RA tests given the number of positive RT-PCR tests on or beyond a specified threshold day after symptom onset is

$$L_C = \log\left(\sum_{k=S}^{T} p(t + t_S)^{k}(1 - p(t + t_S))^{T-k}\right), \qquad \text{(S11)}$$

where $t$ is the time specified for the threshold day, $S$ is the number of successes at the threshold day, and $T$ is the total number of tests conducted.

We stipulated a domain for $\beta_1 < 0$, reflecting our empirical knowledge that the RA tests become less sensitive than RT-PCR tests as the time of the case with the disease proceeds. [1–3]

To account for uncertainty in the RA test percent positive agreement with RT-PCR, we computed the likelihood values from a grid search of the two parameters from the linear logistic regression model. Using the likelihood values from this grid search, the parameters for the linear logistic regression model were obtained through importance sampling.

The percent positive agreement data for all RA tests—with the exception of the BinaxNOW test—is available for days after the symptom onset either for specific days, ranges, or threshold. The BinaxNOW percent positive agreement data included six individuals that tested positive through RT-PCR on an unspecified day greater than seven days from symptom onset, three of which tested positive through the antigen test. We assumed that these samples were obtained on days not covered by the reported daily ranges of 8–10 days after symptom onset or 11–14 days after symptom onset, and thus were obtained 15 or more days after symptom onset.

None of the percent positive agreement datasets included data spanning the time prior to symptom onset. To infer the percent positive agreement during the incubation period, we constructed a mapping between the relative infectivity function and the percent positive agreement post-symptom onset [4].

We classified the RA tests into three sensitivity categories based on temporal patterns of percent positive agreement that were i) stable, ii) gradually declining, or iii) rapidly declining. If the percent positive agreement at the time of symptom onset decreased less than 1% by 40 days after symptom onset for a test, then the test was classified as stable. A test was classified as rapid declining if the percent positive agreement was less than 1% at 20 days after symptom onset (considered the maximum duration of disease [5–7]). A test was classified as gradually declining if its pattern of percent positive agreement over time did not satisfy either of the criteria for relatively constant or rapid decline.

*False-negative for serial testing strategies*

The specificity of the RT-PCR test is 99.9%.[8] The specificity of RA tests is calculated as the product of the RT-PCR specificity and the negative percent agreement of the RA test (**Table S4**). The uncertainty in the specificity of RT-PCR was obtained via importance sampling specifying a Binomial distribution for the likelihood function (with the number of successes being the reported number of RT-PCR true positives and the number of reported true positives). Similarly, uncertainty in the negative percent agreement was obtained via importance sampling specifying a Binomial distribution for the likelihood function (with the number of successes being the reported number of RT-PCR confirmed RA positives and the number of reported RT-PCR positives).

*Diagnostic sensitivity of the RT-PCR assay*

To determine the temporal diagnostic sensitivity of the RT-PCR assay, we used data on serial testing conducted within a healthcare setting [9]. The corresponding data from Hellewell et al [9] includes symptomaticity status on the testing date, the outcome of the self-administered nasal RT-PCR test, as well as the Ct value. Serology testing was also provided in the dataset, but not utilized in our analysis. We applied the methodology of Hellewell et al [9], with two exceptions: we used the distribution for the duration of the incubation period reported by Zhang et al [10] (to approximate the diagnostic sensitivity for the Delta variant) and we fit a log-Normal distribution for the diagnostic sensitivity function to RT-PCR test results from 27 individuals to determine its shape and scale parameters under the constraint that the timing of the peak of the log-Normal distribution equaled the timing of the peak of the infectivity profile. The parameters of the diagnostic sensitivity curve of the RT-PCR assay were determined by maximizing the log-likelihood

$$L = L_T + L_P, \tag{S12}$$

where $L_T$ is the log-likelihood for the time of infection, and $L_P$ is the log-likelihood of the RT-PCR test results.

Given the date $t_i^L$ at which they were last asymptomatic and the date $t_i^F$ at which they are first symptomatic, the log-likelihood for times of infection for the $N$ individuals is

$$L_T = \sum_{i=1}^{N} \log\Big(F(t_i^F - T_i) - F(t_i^L - T_i)\Big), \tag{S13}$$

where $F$ is the cumulative distribution of the incubation period. This log-likelihood accounts for the censoring of the time symptoms appeared between the testing times to determine the time of infection [9]. For the specified diagnostic sensitivity of the test for all individuals and $M_i$ tests for individual $i$, the likelihood for the test result is expressed by

$$L_P = \sum_{i=1}^{N}\sum_{j=1}^{M_i} \log\left(s(x_{i,j})^{R_{i,j}}(1 - s(x_{i,j}))^{1-R_{i,j}}\right), \tag{S14}$$

where $R_{i,j}$ is the result of test $j$ for individual $i$ (if the test is positive then $R_{i,j} = 1$; otherwise, $R_{i,j} = 0$), and $s(t)$ is the diagnostic sensitivity at time $t$ post-infection.

To estimate the time of infection, we specified the upper bound for the time of infection for an individual to be the minimum of the day of their first positive test, their first day of symptoms, and their first day of a non-zero cycle threshold. The corresponding lower bound was set to 30 days prior to this data-driven upper bound.

The two main differences between the method of Hellewell et al [9] and our approach are the distribution of the incubation period and the function for the diagnostic sensitivity. The inference from Hellewell et al [9] applies the distribution of the incubation period from Lauer et al [11], inferred during the period when wild-type SARS CoV-2 was dominant and has a mean of 5.5 days, while we estimate the time of infection based on the distribution reflective of the Delta variant of concern reported by Zhang et al [10]—based on 47 confirmed cases which resulted in a point estimate mean of 4.4 days. For the diagnostic sensitivity, Hellewell et al [9] used a piecewise logistic regression. Here we modeled the diagnostic sensitivity using the probability density function of a log-Normal distribution. Denoting the scaling coefficient as $C$, the shape coefficient as $z$, and the coefficient determining peak diagnostic sensitivity as $K$, we quantified the continuous probability that the RT-PCR test is positive given a sample from an infected patient (i.e., diagnostic sensitivity) at time $t$ with the log-Normal probability density function

$$s(t) = \frac{C}{tz\sqrt{2\pi}} \exp\left\{\frac{-(\log(t)-K)^2}{2z^2}\right\}, \tag{S15}$$

where $s(0) = 0$. To obtain the criterion that the diagnostic sensitivity peaks at the same time as the infectivity profile, we calculated the coefficient determining peak diagnostic sensitivity

$$K = \ln(t_p) + z^2, \tag{S16}$$

where $t_P$ is the time of the peak in the infectivity profile and $z$ is estimated in the model fitting.

To account for uncertainty in the RT-PCR diagnostic sensitivity curve, we fixed the time of infection for the 27 cases to the maximum likelihood estimated value and computed the likelihood values from a grid search of the scaling coefficient and shape coefficient. Using the likelihood values from this grid search, the scaling coefficient and shape coefficient used in the uncertainty analysis were obtained via importance sampling.

To examine the impact of the incubation period on the RT-PCR diagnostic sensitivity curve, we used the alternative distribution of the incubation period from Ashcroft et al [12], which has a mean of 5.72 days. For this alternative scenario, we also use the distribution of the generation time for the infectivity over the course of disease reported by Ashcroft et al [12].

To examine the impact of the functional form of the RT-PCR diagnostic sensitivity curve, we pursued an alternative approach to the quantification of the continuous probability that at time $t$ the RT-PCR test from an infected individual is positive (i.e., diagnostic sensitivity) with the log-Student's $t$ probability density function

$$s(t) = \frac{C}{\sqrt{z\pi}} \frac{\Gamma\left(\frac{z+1}{2}\right)}{\Gamma\left(\frac{z}{2}\right)} \frac{1}{\left(1+(\ln(Kt))^2/z\right)^{(z+1)/2}}, \quad \text{(S17)}$$

where $s(t) = 0$ for $t \leq 0$. To obtain the criterion that the diagnostic sensitivity peaks at the same time as the infectivity profile, we calculated the coefficient determining peak diagnostic sensitivity

$$K = 1/t_P, \quad \text{(S18)}$$

where $t_P$ is the time of the peak in the infectivity profile.

Our inference of the temporal diagnostic sensitivity of RT-PCR over the course of disease differs from that of Hellewell et al [9]. In our three scenarios in which the temporal diagnostic sensitivity of RT-PCR is determined, we estimate a greater peak in diagnostic sensitivity (100%; 95% Credible Interval [CrI]: 86.5%–100%) ; 87.2% (95% CrI: 69.2%–99.1%); and 96.4%; 95% CrI: 88.0%–99.7%) ;) than Hellewell et al [9] (77%; 95% CrI: 54–88%). The greater diagnostic sensitivity in the peak is compensated by a narrower peak when compared to the breadth of the peak from Hellewell et al [9].

*Estimation of rapid antigen percent positive agreement with RT-PCR during quarantine.*

To compare the results obtained from the paired testing of BD Veritor and an RT-PCR testing in a pre-screened cohort with that of our analytical framework, we computed the probability that both BD Veritor and RT-PCR test produce a positive on entry to quarantine, day three of quarantine, and day four.

Specifying random entry into quarantine in the absence of symptoms, the diagnostic sensitivity of the rapid antigen test $s_A(t)$, and the diagnostic sensitivity of RT-PCR $s(t)$, the probability that both BD Veritor and RT-PCR test produce a positive on entry to quarantine for a soon-to-be symptomatic is

$$Q_{S,0} = \frac{1}{t_S} \int_{u=0}^{t_S} s_A(u) \cdot s(u) \mathrm{d}u, \tag{S19}$$

and for an individual never exhibiting symptoms,

$$Q_{A,0} = \frac{1}{t_e} \int_{u=0}^{t_e} s_A(u) \cdot s(u) \mathrm{d}u. \tag{S20}$$

Individuals that test positive with RT-PCR—irrespective of the result from the rapid antigen test—or exhibit symptoms within quarantine are removed from the quarantine environment and placed in isolation. For a soon-to-be symptomatic individual, the probability that both BD Veritor and RT-PCR test produce a positive on day three of quarantine is

$$Q_{S,3} = \frac{1}{C_{S,0}} \int_{u=0}^{t_S-3} (1 - s(u)) \cdot s_A(u + 3) \cdot s(u + 3) \mathrm{d}u, \tag{S21}$$

where

$$C_{S,0} = \int_{u=0}^{t_S-3} (1 - s(u)) \mathrm{d}u. \tag{S22}$$

For an asymptomatic individual, the probability that both BD Veritor and RT-PCR test produce a positive on day three of quarantine is

$$Q_{A,3} = \frac{1}{C_{A,0}} \int_{u=0}^{t_e} (1 - s(u)) \cdot s_A(u + 3) \cdot s(u + 3) \mathrm{d}u, \tag{S23}$$

where

$$C_{A,0} = \int_{u=0}^{t_e} (1 - s(u))\mathrm{d}u. \tag{S24}$$

For a soon-to-be symptomatic, the probability that both BD Veritor and RT-PCR test produce a positive on day four of quarantine is

$$Q_{S,4} = \frac{1}{C_{S,3}} \int_{u=0}^{t_S-4} (1 - s(u)) \cdot (1 - s(u + 3)) \cdot s_A(u + 4) \cdot s(u + 4)\mathrm{d}u, \tag{S25}$$

where

$$C_{S,3} = \int_{u=0}^{t_S-4} (1 - s(u)) \cdot (1 - s(u + 3))\mathrm{d}u. \tag{S26}$$

For an asymptomatic individual, the probability that both BD Veritor and RT-PCR test produce a positive on day three of quarantine is

$$Q_{A,4} = \frac{1}{C_{A,3}} \int_{u=0}^{t_e} (1 - s(u)) \cdot (1 - s(u + 3)) \cdot s_A(u + 4) \cdot s(u + 4)\mathrm{d}u, \tag{S27}$$

where

$$C_{A,3} = \int_{u=0}^{t_e} (1 - s(u)) \cdot (1 - s(u + 3))\mathrm{d}u. \tag{S28}$$

The probability that both BD Veritor and RT-PCR test produce a positive later in quarantine will be weighted more towards individuals that will never exhibit symptoms, as individuals entering quarantine who are soon-to-be symptomatic are more likely to have exhibited symptoms by the end of the quarantine. Specifying the proportion of asymptomatic infections to be $p_A$ and the duration of the incubation period to be $t_S$, the proportion of asymptomatic infections that will be on day $t$ of quarantine is

$$\tilde{p}_A(t) = p_A \max\left\{\frac{t_S - t}{t_S}, 0\right\} + \left(1 - \max\left\{\frac{t_S - t}{t_S}, 0\right\}\right). \tag{S29}$$

Therefore, the probability that both BD Veritor and RT-PCR test produce a positive on day $t$ of quarantine is

$$Q_t = \left(1 - \tilde{p}_A(t)\right)Q_{S,t} + \tilde{p}_A(t)Q_{A,t}. \tag{S30}$$

*Estimation of offshore prevalence of SARS CoV-2*

We adjusted our framework used to estimate the effective reproductive number during serial testing (**Supplementary Material: Frequency of testing to reduce the effective reproduction number**), to estimate whether there were any additional cases in the offshore environment not detected by the serial testing conducted. Specifically, we use the number of RA positives, number of RA false positives, and the FDA empirical diagnostic sensitivity and specificity for the BD Veritor RA test in this inference. Prior to arriving offshore for serial testing, all individuals were tested with RT-PCR on arrival and with test results returned within approximately 24 h. Asserting no transmission offshore (as no observed outbreak occurred during the observation period), any infected individual must have obtained a false-negative RT-PCR test result. As none of the individuals that tested positive exhibited symptoms at the time of testing, the analysis focuses on asymptomatic cases. We specify that any infected individual needs to be infectious after their release from quarantine (i.e. the 24 h delay from obtaining test results). Thus, these individuals must enter quarantine at least a day prior to the end of their infectiousness.

In the absence of individualized data, and to simplify the calculations, the test result on the second day of the screening offshore is independent of the test outcome on the first day of screening. Similarly, the test result on the third day is independent of the test results on the first and second days of screening offshore.

The number of true-positives requires the individual to first test positive with the rapid antigen test and then positive with RT-PCR. Specifying an uninfected individual, the probability of a double false-positive on a given testing day is

$$P_{U,j} = \left(1 - \zeta_A\right)\left(1 - \zeta_R\right), \tag{S31}$$

where $\zeta_A$ is the specificity of the rapid antigen test and $\zeta_R$ is the specificity of RT-PCR.

Specifying an infected individual, the probability of having an RT-PCR confirmed RA test on the first test day is

$$P_{I,1} = \frac{1}{C} \int_{u=o}^{t_e - q} h(u) s_A(u + q + t_1) s(u + q + t_1)\, \mathrm{d}u, \quad \text{(S32)}$$

where

$$h(u) = (1 - s(u)) \text{ and} \quad \text{(S33)}$$

$$C = \int_{u=o}^{t_e - q} h(u)\, \mathrm{d}u. \quad \text{(S34)}$$

The probability of having an RT-PCR-confirmed RA test on the second test day is

$$P_{I,2} = \frac{1}{C} \int_{u=o}^{t_e - q} h(u) s_A(u + q + t_2) s(u + q + t_2)\, \mathrm{d}u. \quad \text{(S35)}$$

The probability of having an RT-PCR-confirmed RA test on the third and final test day is

$$P_{I,3} = \frac{1}{C} \int_{u=o}^{t_e - q} h(u) s_A(u + q + t_3) s(u + q + t_3)\, \mathrm{d}u. \quad \text{(S36)}$$

The number of false-positive rapid antigen tests requires the individual to first test positive with the rapid antigen test and then negative with RT-PCR. Specifying an uninfected individual, the probability of a false-positive rapid antigen test on any test day is

$$F_{U,j} = \left(1 - \zeta_A\right)\zeta_R. \quad \text{(S37)}$$

Specifying an infected individual, the probability of a false-positive rapid antigen test on the first test day is

$$F_{I,1} = \frac{1}{C} \int_{u=o}^{t_e - q} h(u) s_A(u + q + t_1)\left(1 - s(u + q + t_1)\right) \mathrm{d}u. \quad \text{(S38)}$$

The probability of a false-positive rapid antigen test on the second test day is

$$F_{I,2} = \frac{1}{C} \int_{u=o}^{t_e - q} h(u) s_A(u + q + t_2)\left(1 - s(u + q + t_2)\right) \mathrm{d}u. \quad \text{(S39)}$$

The probability of a false-positive rapid antigen test on the third and final test day is

$$F_{I,3} = \frac{1}{C} \int_{u=o}^{t_e - q} h(u) s_A(u + q + t_3)\left(1 - s(u + q + t_3)\right) \mathrm{d}u. \quad \text{(S40)}$$

As the number of infected individuals in the screening population increases, the probability of obtaining a false-negative from the rapid antigen test in the cohort will decrease. Specifying an infected individual, the probability of a false-negative from the rapid antigen test on the first test day is

$$F_{N,1} = \frac{1}{C} \int_{u=o}^{t_e - q} h(u)\left(1 - s_A(u + q + t_1)\right) \mathrm{d}u. \tag{S41}$$

The probability of a false-positive rapid antigen test on the second test day is

$$F_{N,2} = \frac{1}{C} \int_{u=o}^{t_e - q} h(u)\left(1 - s_A(u + q + t_2)\right) \mathrm{d}u. \tag{S42}$$

The probability of a false-positive rapid antigen test on the third and final test day is

$$F_{N,3} = \frac{1}{C} \int_{u=o}^{t_e - q} h(u)\left(1 - s_A(u + q + t_3)\right) \mathrm{d}u. \tag{S43}$$

We have assumed that there was no transmission offshore. Thus, we need to minimize the probability of post-quarantine transmission for all the infected individuals. Specifying isolation after a positive RT-PCR test, the expected transmission from an asymptomatic infected individual under the specified offshore screening protocol

$$\begin{aligned} R = & \frac{1}{C} \int_{u=0}^{t_e - q} \int_{t=q}^{q+t_1} h(u)\, r(t + u) \mathrm{d}t\, \mathrm{d}u + \frac{1}{C} \int_{u=0}^{t_e - q} \int_{t=q+t_1}^{q+t_2} h(u)\, m_1(u) r(t + u) \mathrm{d}t\, \mathrm{d}u \\ & + \frac{1}{C} \int_{u=0}^{t_e - q} \int_{t=q+t_2}^{q+t_3} h(u) m_1(u) m_2(u) r(t + u) \mathrm{d}t\, \mathrm{d}u \\ & + \frac{1}{C} \int_{u=0}^{t_e - q} \int_{t=q+t_3}^{\infty} h(u)\, m_1(u) m_2(u) m_3(u) r(t + u) \mathrm{d}t\, \mathrm{d}u, \end{aligned} \tag{S44}$$

where

$$m_1(u) = 1 - s_A(u + q + t_1) s(u + q + t_1), \tag{S45}$$

$$m_2(u) = 1 - s_A(u + q + t_2) s(u + q + t_2), \tag{S46}$$

and

$$m_3(u) = 1 - s_A(u + q + t_3)s(u + q + t_3). \tag{S47}$$

Over the course of the data collection, the number of participants on each testing day was not constant (e.g. 124 tested on day two, and 96 tested on day eight). To account for changes in the cohort size, we specify the same reduction in the number of infected individuals. Denoting the number of people tested in the $i$th round of testing by $N_i$ and specifying $T_I$ infected individuals, the number of infected individuals that have remained offshore is $\frac{N_i}{N_1}T_I$.

Specifying an initial $T_U$ uninfected individuals and $T_I$ infected individuals within the cohort for testing strategy $j$, the expected number of false-positives on testing day $i$ is

$$F_{i,j} = \frac{N_i}{N_1}\left(T_U F_{U,1} + T_I F_{I,1}\right), \tag{S48}$$

and the number of RT-PCR confirmed positives is

$$P_{i,j} = \frac{N_i}{N_1}\left(T_U P_{U,1} + T_I P_{I,1}\right). \tag{S49}$$

and the probability of false-negatives from a rapid antigen test for those not detected in the screening on the first test day is expressed as

$$F_{N,i}^{\;N_i T_I (1-P_{I,i}-F_{I,i})/N_1}. \tag{S50}$$

The likelihood of the outcomes that occurred on the first testing day is

$$L_{i,j} = f(\overline{F}_i | F_i) \cdot f(\overline{P}_i | P_i) \cdot F_{N,i}^{\;N_i T_I (1-P_{I,i}-F_{I,i})/N_1} \tag{S51}$$

where $\overline{F}_i$ is the number of observed false-positives on the testing day $i$, $\overline{P}_i$ is the number of RT-PCR confirmed positives on testing day $i$, and $f(X|Y)$ is the Poisson distribution with mean $Y$.

To estimate that the number of infected individuals that were offshore with a probability of onward transmission $p$ for the two testing strategies—computed from the expected transmission as in Eq. S44—we maximized

$$\prod_{j=1}^{2}\left(1-p_j\right)^{T_{I,j}}\prod_{i=1}^{3}L_{i,j}. \quad (S52)$$

The effectiveness of serial testing conducted offshore in identifying asymptomatic cases is

$$\min\left\{\frac{\sum_{j=1}^{2}\bar{P}_{i,j}}{\sum_{j=1}^{2}T_{I,j}}, 1\right\}. \quad (S53)$$

*Comparison of the internal and external percent positive agreement data sets*

To determine if the internal (i.e., USA FDA) dataset differs significantly from the external (i.e. independently conducted outside a controlled setting), we used the likelihood ratio test. We calculated the log-likelihood for the linear logistic regression curve for the i) the internal dataset ($L_I$ ), ii) the external dataset ($L_E$ ), and iii) from combining the two datasets ($L_{IE}$ ). The internal and external datasets are significantly different from each other if

$$L_I + L_E - L_{IE} \geq \chi^2_{0.95,\,2}/2\,,$$

where $\chi^2_{0.95,\,2}$ is the 95[th] percentile of the Chi-squared distribution with two degrees of freedom.

Otherwise, if

$$L_I + L_E - L_{IE} < \chi^2_{0.95,\,2}/2\,,$$

then the two datasets are not significantly different from each other.

### Supplementary tables

**Table S1.** The percent positive agreement data for the rapid antigen tests based on the day of symptom onset

<table>
<tr><th>Test name</th><th colspan="16">Days since symptom onset</th></tr>
<tr><td></td><td>0</td><td>1</td><td>2</td><td>3</td><td>4</td><td>5</td><td>6</td><td>7</td><td>8</td><td>9</td><td>10</td><td>11</td><td>12</td><td>13</td><td>14</td><td>>14</td></tr>
<tr><td>BD Veritor [a,b,c]</td><td>•</td><td>7/8</td><td>10/12</td><td>1/2</td><td>5/5</td><td>3/4</td><td>2/3</td><td>1/4</td><td>•</td><td>•</td><td>•</td><td>•</td><td>•</td><td>•</td><td>•</td><td>•</td></tr>
<tr><td>BinaxNOW [b,c]</td><td>•</td><td>10/12</td><td>18/22</td><td>13/16</td><td>9/13</td><td>13/15</td><td>12/12</td><td>24/27</td><td colspan="3">19/27</td><td colspan="4">8/17</td><td>3/6</td></tr>
<tr><td>BinaxNOW [c,d]</td><td>7/8</td><td>30/33</td><td>30/32</td><td>22/22</td><td>8/9</td><td>6/6</td><td>7/7</td><td>2/2</td><td colspan="8">12/17</td></tr>
<tr><td>CareStart [b,c]</td><td>3/3</td><td>7/8</td><td>11/13</td><td>8/9</td><td>3/4</td><td>2/2</td><td>•</td><td>•</td><td>•</td><td>•</td><td>•</td><td>•</td><td>•</td><td>•</td><td>•</td><td>•</td></tr>
<tr><td>CareStart [b,e]</td><td>•</td><td>•</td><td>7/7</td><td>8/8</td><td>7/8</td><td>8/9</td><td>•</td><td>•</td><td>•</td><td>•</td><td>•</td><td>•</td><td>•</td><td>•</td><td>•</td><td>•</td></tr>
<tr><td>CareStart [c,d]</td><td>4/5</td><td>7/9</td><td>8/9</td><td>11/12</td><td>6/8</td><td>3/3</td><td>3/4</td><td>1/1</td><td colspan="8">2/9</td></tr>
<tr><td>Celltrion DiaTrust [b,e]</td><td>•</td><td>3/3</td><td>6/6</td><td>5/6</td><td>9/9</td><td>2/2</td><td>1/1</td><td>2/3</td><td>•</td><td>•</td><td>•</td><td>•</td><td>•</td><td>•</td><td>•</td><td>•</td></tr>
<tr><td>Clip COVID [b,c]</td><td>•</td><td>4/5</td><td>12/12</td><td>8/8</td><td>6/6</td><td>1/1</td><td>•</td><td>•</td><td>•</td><td>•</td><td>•</td><td>•</td><td>•</td><td>•</td><td>•</td><td>•</td></tr>
<tr><td>Ellume [b,f]</td><td colspan="2">7/7</td><td>6/6</td><td>5/5</td><td>3/3</td><td>1/1</td><td>3/3</td><td>0/1</td><td>•</td><td>•</td><td>•</td><td>•</td><td>•</td><td>•</td><td>•</td><td>•</td></tr>
<tr><td>Liaison [b,c]</td><td>6/6</td><td>5/6</td><td>2/2</td><td>2/2</td><td>0/0</td><td>2/2</td><td>4/4</td><td>1/1</td><td>4/4</td><td>4/4</td><td>•</td><td>•</td><td>•</td><td>•</td><td>•</td><td>•</td></tr>
<tr><td>Liaison [b,e]</td><td>8/9</td><td>9/9</td><td>4/4</td><td>10/10</td><td>5/6</td><td>3/3</td><td>3/3</td><td>3/3</td><td>1/1</td><td>2/2</td><td>1/1</td><td>•</td><td>•</td><td>•</td><td>•</td><td>•</td></tr>
<tr><td>LumiraDx [b,c]</td><td>6/6</td><td>6/6</td><td>16/16</td><td>9/9</td><td>17/18</td><td>6/6</td><td>6/6</td><td>6/6</td><td>2/2</td><td>0/0</td><td>2/2</td><td>3/3</td><td>2/3</td><td>•</td><td>•</td><td>•</td></tr>
<tr><td>LumiraDx [b,e]</td><td>2/2</td><td>4/4</td><td>3/3</td><td>8/8</td><td>5/5</td><td>1/1</td><td>3/3</td><td>8/8</td><td>2/2</td><td>0/0</td><td>2/3</td><td>1/1</td><td>•</td><td>•</td><td>•</td><td>•</td></tr>
<tr><td>Omnia [b,c]</td><td>1/1</td><td>5/5</td><td>7/9</td><td>11/13</td><td>19/21</td><td>2/2</td><td>6/6</td><td>•</td><td>•</td><td>•</td><td>•</td><td>•</td><td>•</td><td>•</td><td>•</td><td>•</td></tr>
<tr><td>SCoV-2 Ag Detect [b,c]</td><td>•</td><td>7/8</td><td>5/6</td><td>14/16</td><td>10/11</td><td>3/4</td><td>•</td><td>•</td><td>•</td><td>•</td><td>•</td><td>•</td><td>•</td><td>•</td><td>•</td><td>•</td></tr>
<tr><td>Simoa [b,e]</td><td>8/8</td><td>14/15</td><td>11/11</td><td>10/10</td><td>9/9</td><td>4/4</td><td>4/4</td><td>8/8</td><td colspan="7">18/19</td><td>•</td></tr>
<tr><td>Sofia [b,c]</td><td>•</td><td>5/5</td><td>11/12</td><td>3/3</td><td>5/5</td><td>2/2</td><td>2/2</td><td>1/1</td><td>•</td><td>•</td><td>•</td><td>•</td><td>•</td><td>•</td><td>•</td><td>•</td></tr>
<tr><td rowspan="2">Sofia [c,d]</td><td>1/1</td><td>6/6</td><td>5/6</td><td>7/7</td><td>1/2</td><td>•</td><td>•</td><td>•</td><td>3/3</td><td>•</td><td>•</td><td>•</td><td>•</td><td>•</td><td>•</td><td>•</td></tr>
<tr><td colspan="6">3/9</td><td colspan="10">5/5</td></tr>
<tr><td>Sofia 2 Flu + SARS [b,e]</td><td>0/0</td><td>14/14</td><td>10/11</td><td>4/4</td><td>10/10</td><td>2/3</td><td>•</td><td>•</td><td>•</td><td>•</td><td>•</td><td>•</td><td>•</td><td>•</td><td>•</td><td>•</td></tr>
<tr><td>Status COVID-19/Flu [b,e]</td><td>16/18</td><td>13/13</td><td>6/6</td><td>6/6</td><td>5/5</td><td>0/1</td><td>•</td><td>•</td><td>•</td><td>•</td><td>•</td><td>•</td><td>•</td><td>•</td><td>•</td><td>•</td></tr>
</table>

| | | | | | | | | | | | | | | | | |
|---|---|---|---|---|---|---|---|---|---|---|---|---|---|---|---|---|
| Vitros [b,e] | 4/8 | 2/2 | 4/4 | 6/6 | 0/0 | 2/2 | 3/4 | 3/4 | • | • | • | • | • | • | • | • |

• Indicates that there was no data available for the specified time period
[a] Peer-reviewed
[b] Data from EUA submission
[c] Anterior nasal swab
[d] Data from community testing
[e] Nasopharyngeal swab
[f] Mid-turbinate swab

**Table S2.** Estimated coefficients and 95% credible interval (generated from 1,000 samples) for the logistic regression models describing the percent positive agreement curve for each of the rapid antigen tests

| Test name | $\beta_0$ - | $\beta_1$ |
|---|---|---|
| BD Veritor [a,b,c] | 2.55<br>(1.1–4.68) | $-0.379$<br>($-0.852$–$-0.0609$) |
| BinaxNOW [b,c] | 1.95<br>(1.25–2.85) | $-0.1$<br>($-0.214$–$-0.0066$) |
| BinaxNOW [c,d] | 2.91<br>(2.2–3.81) | $-1.93 \times 10^{-9}$<br>($-1.03 \times 10^{-8}$–$3.57 \times 10^{-13}$) |
| BinaxNOW [c,e] | 2.5<br>(1.92–3.17) | $-0.152$<br>($-0.247$–$-0.0612$) |
| CareStart [b,c] | 2.2<br>(1.39–4.02) | $-0.123$<br>($-0.616$–$-0.00178$) |
| CareStart [b,f] | 6.77<br>(2.16–20) | $-0.998$<br>($-3.7$–0) |
| CareStart [c,d] | 1.87<br>(1.18–2.69) | $-2.19 \times 10^{-11}$<br>($-1.16 \times 10^{-10}$–$2.22 \times 10^{-13}$) |
| CareStart [c,e] | 1.89<br>(1.35–2.51) | $-4.9 \times 10^{-10}$<br>($-2.67 \times 10^{-9}$–$-2.04 \times 10^{-11}$) |
| Celltrion DiaTrust [b,f] | 4.81<br>(2.19–11.5) | $-0.518$<br>($-1.66$–$-0.0217$) |
| Clip COVID [b,c] | 3.43<br>(1.77–6.74) | $-1.28 \times 10^{-8}$<br>($-8.08 \times 10^{-8}$–$-2.59 \times 10^{-9}$) |
| Ellume [b,g] | 100<br>(100–490) | $-15.3$<br>($-59.3$–$-3.6$) |
| Liaison [b,c] | 3.47<br>(1.85–6.82) | $-1.15 \times 10^{-9}$<br>($-6.05 \times 10^{-9}$–$3.33 \times 10^{-13}$) |
| Liaison [b,f] | 3.2<br>(1.95–5.23) | $-1.73 \times 10^{-9}$<br>($-9.13 \times 10^{-9}$–$3.61 \times 10^{-13}$) |
| LumiraDx [b,c] | 5.64<br>(1.33–10.9) | $-0.327$<br>($-0.943$–$-0.0382$) |
| LumiraDx [b,f] | 12.1<br>(3.25–47.5) | $-1.05$<br>($-4.52$–$-0.0284$) |
| Omnia [b,c] | 2.14 | $-1.81 \times 10^{-9}$ |

| | | |
|---|---|---|
| | (1.35–3.19) | $(-2.88 \times 10^{-8}$–$-4.02 \times 10^{-10})$ |
| SCoV-2 Ag Detect [b,c] | 2.03<br>(1.34–3.42) | $-0.0537$<br>$(-0.289$–$0)$ |
| Simoa [b,f] | 4.17<br>(3.06–7.28) | $-0.0778$<br>$(-0.352$–$0)$ |
| Sofia [b,c] | 3.37<br>(1.61–6.71) | $-8.89 \times 10^{-10}$<br>$(-1.12 \times 10^{-8}$–$-1.53 \times 10^{-10})$ |
| Sofia [c,d] | 1.35<br>(0.646–2.22) | $-9.23 \times 10^{-10}$<br>$(-1.12 \times 10^{-8}$–$-2.27 \times 10^{-10})$ |
| Sofia [c,e] | 1.9<br>(1.27–2.71) | $-8.98 \times 10^{-10}$<br>$(-4.68 \times 10^{-9}$–$3.4 \times 10^{-13})$ |
| Sofia 2 Flu + SARS [b,c] | 4.77<br>(2.19–11.4) | $-0.6$<br>$(-2.1$–$0)$ |
| Status COVID-19/Flu [b,f] | 2.93<br>(2.02–4.99) | $-0.135$<br>$(-0.667$–$0)$ |
| Vitros [b,f] | 1.39<br>(0.453–2.36) | $-8 \times 10^{-10}$<br>$(-4.23 \times 10^{-9}$–$-2.33 \times 10^{-12})$ |

[a] Peer-reviewed EUA data
[b] Data from EUA submission
[c] Anterior nasal swab
[d] Data from community testing
[e] Data from EUA submission and community testing
[f] Nasopharyngeal swab
[g] Mid-turbinate swab

**Table S3.** Specifying a 4.4-day incubation period, a log-Normal distribution for the temporal RT-PCR diagnostic sensitivity, a basic reproductive number of 3.2, and 35.1% of infections being asymptomatic, the required quarantine durations, serial testing frequencies, and probabilities of false-positives associated with the serial testing frequency.

| Rapid antigen test | Quarantine required | | Serial testing required | |
|---|---|---|---|---|
| | Exit test [a] | Entry and exit test [a] | Frequency [b] | Prob. of a false positive [c] |
| BD Veritor [d,e] | 8 (8–8) | 7 (7–8) | 2 (2–2) | 0.0389 (0.00845–0.152) |
| BinaxNOW [e,f] | 8 (8–8) | 7 (7–7) | 2 (2–2) | 0.104 (0.0428–0.21) |
| BinaxNOW [g,f] | 8 (8–8) | 7 (7–7) | 2 (2–3) | 0.0474 (0.0264–0.0751) |
| BinaxNOW [h,f] | 8 (8–8) | 7 (7–7) | 2 (2–2) | 0.0558 (0.0364–0.0833) |
| CareStart [e,f] | 8 (8–8) | 7 (7–7) | 2 (2–2) | 0.00675 (0.00407–0.33) |
| CareStart [e,i] | 8 (8–8) | 6 (6–7) | 3 (2–3) | 0.0355 (0.00716–0.188) |
| CareStart [f,g] | 8 (8–8) | 7 (7–7) | 2 (2–2) | 0.117 (0.0771–0.17) |
| CareStart [f,h] | 8 (8–8) | 7 (7–7) | 2 (2–2) | 0.112 (0.0739–0.164) |
| Celltrion DiaTrust [e,i] | 8 (8–8) | 6 (6–7) | 3 (2–3) | 0.0488 (0.00846–0.26) |
| Clip COVID [e,f] | 8 (8–8) | 7 (6–7) | 3 (2–3) | 0.00451 (0.00315–0.144) |
| Ellume [e,j] | 8 (8–8) | 6 (6–7) | 3 (2–3) | 0.141 (0.0628–0.357) |
| Liaison [e,f] | 8 (8–8) | 7 (6–7) | 3 (2–3) | 0.00451 (0.00325–0.175) |
| Liaison [e,i] | 8 (8–8) | 7 (6–7) | 2 (2–3) | 0.0575 (0.00863–0.221) |
| LumiraDX [e,f] | 8 (8–8) | 6 (6–7) | 3 (2–3) | 0.155 (0.0767–0.374) |
| LumiraDX [e,i] | 8 | 6 | 3 | 0.108 |

| | | | | |
|---|---|---|---|---|
| | (8–8) | (6–7) | (2–3) | (0.0473–0.28) |
| Omnia [e,f] | 8<br>(8–8) | 7<br>(7–7) | 2<br>(2–2) | 0.00675<br>(0.00419–0.478) |
| SCoV-2 Ag Detect [e,f] | 8<br>(8–8) | 7<br>(7–7) | 2<br>(2–2) | 0.00675<br>(0.00387–0.0857) |
| Simoa [e,i] | 8<br>(8–8) | 6<br>(6–7) | 3<br>(2–3) | 0.00451<br>(0.00323–0.393) |
| Sofia [e.f] | 8<br>(8–8) | 7<br>(6–7) | 3<br>(2–3) | 0.00451<br>(0.00329–0.113) |
| Sofia [g,f] | 8<br>(8–8) | 7<br>(7–8) | 2<br>(1–2) | 0.109<br>(0.0637–0.241) |
| Sofia [h,f] | 8<br>(8–8) | 7<br>(7–7) | 2<br>(2–2) | 0.0944<br>(0.0586–0.145) |
| Sofia 2 Flu+SARS [e,f] | 8<br>(8–8) | 7<br>(6–7) | 3<br>(2–3) | 0.00451<br>(0.00308–0.15) |
| Status COVID-19/Flu [e,i] | 8<br>(8–8) | 7<br>(6–7) | 2<br>(2–3) | 0.00675<br>(0.00363–0.243) |
| Vitros [e,i] | 8<br>(8–8) | 7<br>(7–8) | 2<br>(1–2) | 0.00675<br>(0.00402–0.284) |

[a] Quarantine durations that are equivalent or better than a 7-day quarantine with an RT-PCR test conducted 24 h before exit.
[b] The minimum required testing frequency for serial testing such that the effective reproductive number is less than one.
[c] The probability of at least one false positive in a two-week period of serial testing under the minimum required testing frequency.
[d] Peer-reviewed
[e] Data from EUA submission
[f] Anterior nasal swab
[g] Data from community testing
[h] Combined data from EUA submission and community testing
[i] Nasopharyngeal swab
[j] Mid-turbinate swab

**Table S4.** Specifying a 4.4 day incubation period, a log-Student's *t* distribution for the temporal RT-PCR diagnostic sensitivity , a basic reproductive number of 3.2, and 35.1% of infections being asymptomatic, the required quarantine durations, serial testing frequencies, and probabilities of false-positives associated with the serial testing frequency.

| Rapid antigen test | Quarantine required | | Serial testing required | |
|---|---|---|---|---|
| | Exit test [a] | Entry and exit test [a] | Frequency [b] | Prob. of a false positive [c] |
| BD Veritor [d,e] | 8 (8–8) | 7 (7–8) | 2 (1–2) | 0.0389 ( 0.00815–0.159) |
| BinaxNOW [e,f] | 8 (8–8) | 7 (7–7) | 2 (1–2) | 0.104 (0.0387–0.244) |
| BinaxNOW [g,f] | 8 (8–8) | 7 (7–7) | 2 (2–3) | 0.0474 (0.0245–0.0761) |
| BinaxNOW [h,f] | 8 (8–8) | 7 (7–7) | 2 (2–3) | 0.0558 (0.0319–0.0882) |
| CareStart [e,f] | 8 (8–8) | 7 (7–7) | 2 (1–2) | 0.00675 (0.00403–0.345) |
| CareStart [e,i] | 8 (8–8) | 7 (6–7) | 2 (2–3) | 0.0528 (0.00736–0.196) |
| CareStart [f,g] | 8 (8–8) | 7 (7–7) | 2 (1–2) | 0.117 (0.0718–0.236) |
| CareStart [f,h] | 8 (8–8) | 7 (7–7) | 2 (1–2) | 0.112 (0.0686–0.223) |
| Celltrion DiaTrust [e,i] | 8 (8–8) | 7 (6–7) | 2 (2–3) | 0.0723 (0.00852–0.269) |
| Clip COVID [e,f] | 8 (8–8) | 7 (6–7) | 2 (2–3) | 0.00675 (0.00324–0.145) |
| Ellume [e,j] | 8 (8–8) | 7 (6–7) | 2 (2–3) | 0.204 (0.0669–0.371) |
| Liaison [e,f] | 8 (8–8) | 7 (6–7) | 2 (2–3) | 0.00675 (0.00316–0.174) |
| Liaison [e,i] | 8 (8–8) | 7 (6–7) | 2 (2–3) | 0.0575 (0.00851–0.222) |
| LumiraDX [e,f] | 8 (8–8) | 7 (6–7) | 2 (2–3) | 0.223 (0.0808–0.385) |
| LumiraDX [e,i] | 8 (8–8) | 6 (6–7) | 2 (2–3) | 0.158 (0.0499–0.291) |
| Omnia [e,f] | 8 (8–8) | 7 (7–7) | 2 (1–2) | 0.00675 (0.00439–0.497) |

| | | | | |
|---|---|---|---|---|
| SCoV-2 Ag Detect [e,f] | 8 (8–8) | 7 (7–7) | 2 (1–2) | 0.00675 (0.0038–0.0925) |
| Simoa [e,i] | 8 (8–8) | 7 (6–7) | 2 (2–3) | 0.00675 (0.00336–0.401) |
| Sofia [e.f] | 8 (8–8) | 7 (6–7) | 2 (2–3) | 0.00675 (0.00325–0.113) |
| Sofia [h,f] | 8 (8–8) | 7 (7–8) | 2 (1–2) | 0.109 (0.0678–0.273) |
| Sofia [g,f] | 8 (8–8) | 7 (7–7) | 2 (1–2) | 0.0944 (0.0534–0.19) |
| Sofia 2 Flu+SARS [e,f] | 8 (8–8) | 7 (6–7) | 2 (2–3) | 0.00675 (0.00309–0.155) |
| Status COVID-19/Flu [e,i] | 8 (8–8) | 7 (6–7) | 2 (2–3) | 0.00675 (0.0035–0.242) |
| Vitros [e,i] | 8 (8–8) | 7 (7–8) | 2 (1–2) | 0.00675 (0.00409–0.337) |

[a] Quarantine durations that are equivalent or better than a 7-day quarantine with an RT-PCR test conducted 24 h before exit.
[b] The minimum required testing frequency for serial testing such that the effective reproductive number is less than one.
[c] The probability of at least one false positive in a two-week period of serial testing under the minimum required testing frequency.
[d] Peer-reviewed
[e] Data from EUA submission
[f] Anterior nasal swab
[g] Data from community testing
[h] Combined data from EUA submission and community testing
[i] Nasopharyngeal swab
[j] Mid-turbinate swab

**Table S5.** Specifying a 5.72-day incubation period, a log-Normal distribution for the temporal RT-PCR diagnostic sensitivity , a basic reproductive number of 3.2, and 35.1% of infections being asymptomatic, the required quarantine durations, serial testing frequencies, and probabilities of false-positives associated with the serial testing frequency.

| Rapid antigen test | Quarantine required | | Serial testing required | |
|---|---|---|---|---|
| | Exit test [a] | Entry and exit test [a] | Frequency [b] | Prob. of a false positive [c] |
| BD Veritor [d,e] | 9<br>(8–9) | 9<br>(8–9) | 2<br>(2–2) | 0.0389<br>(0.00847–0.155) |
| BinaxNOW [e,f] | 9<br>(8–9) | 9<br>(8–9) | 2<br>(2–2) | 0.104<br>(0.0415–0.217) |
| BinaxNOW [g,f] | 9<br>(8–9) | 8<br>(8–9) | 2<br>(2–3) | 0.0474<br>(0.0255–0.0751) |
| BinaxNOW [h,f] | 9<br>(8–9) | 9<br>(8–9) | 2<br>(2–2) | 0.0558<br>(0.0357–0.0841) |
| CareStart [e,f] | 9<br>(8–9) | 9<br>(8–9) | 2<br>(2–2) | 0.00675<br>(0.00408–0.333) |
| CareStart [e,i] | 8<br>(8–9) | 8<br>(8–9) | 3<br>(2–3) | 0.0355<br>(0.00729–0.192) |
| CareStart [f,g] | 9<br>(8–9) | 9<br>(8–9) | 2<br>(2–2) | 0.117<br>(0.0752–0.173) |
| CareStart [f,h] | 9<br>(8–9) | 9<br>(8–9) | 2<br>(2–2) | 0.112<br>(0.0729–0.166) |
| Celltrion DiaTrust [e,i] | 8<br>(8–9) | 8<br>(8–9) | 2<br>(2–3) | 0.0723<br>(0.009–0.267) |
| Clip COVID [e,f] | 8<br>(8–9) | 8<br>(8–9) | 2<br>(2–3) | 0.00675<br>(0.00331–0.145) |
| Ellume [e,j] | 8<br>(8–9) | 8<br>(8–9) | 3<br>(2–3) | 0.141<br>(0.0642–0.363) |
| Liaison [e,f] | 8<br>(8–9) | 8<br>(8–9) | 2<br>(2–3) | 0.00675<br>(0.00314–0.174) |
| Liaison [e,i] | 8<br>(8–9) | 8<br>(8–9) | 2<br>(2–3) | 0.0575<br>(0.00868–0.222) |
| LumiraDX [e,f] | 8<br>(8–9) | 8<br>(8–9) | 3<br>(2–3) | 0.155<br>(0.0796–0.381) |
| LumiraDX [e,i] | 8<br>(8–9) | 8<br>(8–9) | 3<br>(2–3) | 0.108<br>(0.0489–0.287) |

| | | | | |
|---|---|---|---|---|
| Omnia [e,f] | 9 (8–9) | 9 (8–9) | 2 (2–2) | 0.00675 (0.00444–0.481) |
| SCoV-2 Ag Detect [e,f] | 9 (8–9) | 9 (8–9) | 2 (2–2) | 0.00675 (0.00389–0.0871) |
| Simoa [e,i] | 8 (8–9) | 8 (8–9) | 2 (2–3) | 0.00675 (0.00337–0.398) |
| Sofia [e.f] | 8 (8–9) | 8 (8–9) | 2 (2–3) | 0.00675 (0.00323–0.113) |
| Sofia [h,f] | 9 (8–9) | 9 (8–9) | 2 (1–2) | 0.109 (0.0658–0.261) |
| Sofia [g,f] | 9 (8–9) | 9 (8–9) | 2 (2–2) | 0.0944 (0.0576–0.147) |
| Sofia 2 Flu+SARS [e,f] | 8 (8–9) | 8 (8–9) | 2 (2–3) | 0.00675 (0.00314–0.155) |
| Status COVID-19/Flu [e,i] | 9 (8–9) | 9 (8–9) | 2 (2–3) | 0.00675 (0.00362–0.243) |
| Vitros [e,i] | 9 (8–9) | 9 (8–9) | 2 (1–2) | 0.00675 (0.00406–0.308) |

[a] Quarantine durations that are equivalent or better than a 7-day quarantine with an RT-PCR test conducted 24 h before exit.
[b] The minimum required testing frequency for serial testing such that the effective reproductive number is less than one.
[c] The probability of at least one false positive in a two-week period of serial testing under the minimum required testing frequency.
[d] Peer-reviewed
[e] Data from EUA submission
[f] Anterior nasal swab
[g] Data from community testing
[h] Combined data from EUA submission and community testing
[i] Nasopharyngeal swab
[j] Mid-turbinate swab

**Table S6.** The change in the required quarantine durations and serial testing frequencies when there is a specified threshold in which the rapid antigen test can return a positive test result relative to the results in Table S1 for the maximum likelihood estimates.

| Rapid antigen test | Threshold based on infectivity at 5.6 days after symptom onset | | | Threshold based on infectivity at 10 days after symptom onset | | |
|---|---|---|---|---|---|---|
| | Exit test [a] | Entry and exit test [a] | Frequency [b] | Exit test [a] | Entry and exit test [a] | Frequency [b] |
| BD Veritor [d,e] | 0 | 0 | 0 | 0 | 0 | 0 |
| BinaxNOW [e,f] | 0 | 0 | 0 | 0 | 0 | 0 |
| BinaxNOW [g,f] | 0 | 0 | 0 | 0 | 0 | 0 |
| BinaxNOW [h,f] | 0 | 0 | 0 | 0 | 0 | 0 |
| CareStart [e,f] | 0 | 0 | 0 | 0 | 0 | 0 |
| CareStart [e,i] | 0 | 1 | 0 | 0 | 0 | 0 |
| CareStart [f,g] | 0 | 0 | 0 | 0 | 0 | 0 |
| CareStart [f,h] | 0 | 0 | 0 | 0 | 0 | 0 |
| Celltrion DiaTrust [e,i] | 0 | 1 | 0 | 0 | 0 | 0 |
| Clip COVID [e,f] | 0 | 0 | - 1 | 0 | 0 | 0 |
| Ellume [e,j] | 0 | 0 | 0 | 0 | 0 | 0 |
| Liaison [e,f] | 0 | 0 | - 1 | 0 | 0 | 0 |
| Liaison [e,i] | 0 | 0 | 0 | 0 | 0 | 0 |
| LumiraDX [e,f] | 0 | 1 | 0 | 0 | 0 | 0 |
| LumiraDX [e,i] | 0 | 0 | 0 | 0 | 0 | 0 |
| Omnia [e,f] | 0 | 0 | 0 | 0 | 0 | 0 |
| SCoV-2 Ag Detect [e,f] | 0 | 0 | 0 | 0 | 0 | 0 |
| Simoa [e,i] | 0 | 1 | - 1 | 0 | 0 | 0 |
| Sofia [e.f] | 0 | 0 | - 1 | 0 | 0 | 0 |
| Sofia [h,f] | 0 | 0 | 0 | 0 | 0 | 0 |

| | | | | | | |
|---|---|---|---|---|---|---|
| Sofia [g,f] | 0 | 0 | 0 | 0 | 0 | 0 |
| Sofia 2 Flu+SARS [e,f] | 0 | 0 | - 1 | 0 | 0 | 0 |
| Status COVID-19/Flu [e,i] | 0 | 0 | 0 | 0 | 0 | 0 |
| Vitros [e,i] | 0 | 0 | 0 | 0 | 0 | 0 |

[a] Quarantine durations that are equivalent or better than a 7-day quarantine with an RT-PCR test conducted 24 h before exit.
[b] The minimum required testing frequency for serial testing such that the effective reproductive number is less than one.
[c] The probability of at least one false positive in a two-week period of serial testing under the minimum required testing frequency.
[d] Peer-reviewed
[e] Data from EUA submission
[f] Anterior nasal swab
[g] Data from community testing
[h] Combined data from EUA submission and community testing
[i] Nasopharyngeal swab
[j] Mid-turbinate swab

**Table S7.** The specificity of RT-PCR and the EUA rapid antigen tests and 95% credible interval constructed from 1000 samples.

| Test | Fraction of RA tests in agreement with negative RT-PCR | Specificity (95% CrI) | Data Source | Reference |
|---|---|---|---|---|
| RT-PCR | N/A | 99.90% (99.84%–99.95%) (12392/12404) | Community testing | 8 |
| BD Veritor [a] | 212/213 | 99.43% (97.58%–99.91%) | EUA submission [b] | 13 |
| BinaxNOW [a] | 338/343 | 98.45% (96.60%–99.43%) | EUA submission | 14 |
| BinaxNOW [a] | 2004/2016 | 99.31% (98.91%–99.59%) | Community testing | 15 |
| BinaxNOW [a] | 2342/2359 | 99.18% (98.75%–99.49%) | EUA submission and community testing | 14,15 |
| CareStart [a] | 53/53 | 99.90% (94.40%–99.92%) | EUA submission | 16 |
| CareStart [d] | 147/148 | 99.23% (96.50%–99.90%) | EUA submission | 16 |
| CareStart [a] | 1243/1264 | 98.24% (97.38%–98.90%) | Community testing | 17 |
| CareStart [a] | 1296/1317 | 98.31% (97.52%–98.89%) | EUA submission and community testing | 16,17 |
| Celltrion [d] | 102/103 | 98.93% (95.06%–99.86%) | EUA submission | 18 |
| Clip COVID [a] | 134/134 | 99.90% (97.52%–99.96%) | EUA submission | 19 |
| Ellume [e] | 156/161 | 96.80% (93.11%–98.74%) | EUA submission | 20 |
| Liaison [a] | 108/108 | 99.90% (96.84%–99.96%) | EUA submission | 21 |
| Liaison [d] | 133/134 | 99.16% (96.16%–99.87%) | EUA submission | 21 |

| | | | | |
|---|---|---|---|---|
| LumiraDx [a] | 168/174 | 96.46% (92.83%–98.57%) | EUA submission | 22 |
| LumiraDx [d] | 210/215 | 97.58% (94.73%–99.08%) | EUA submission | 23 |
| Omnia [a] | 32/32 | 99.90% (90.58%–99.92%) | EUA submission | 24 |
| SCoV-2 Ag Detect [a] | 257/257 | 99.90% (98.72%–99.95%) | EUA submission | 25 |
| Simoa [d] | 38/38 | 99.90% (92.14%–99.92%) | EUA submission | 26 |
| Sofia [a] | 179/179 | 99.90% (98.09%–99.96%) | EUA submission | 27 |
| Sofia [a] | 1025/1041 | 98.37% (97.46%–99.03%) | Community testing | 28 |
| Sofia [a] | 1204/1220 | 98.59% (97.77%–99.16%) | EUA submission and community testing | 27,28 |
| Sofia 2 Flu + SARS [a] | 122/122 | 99.90% (97.55%–99.96%) | EUA submission | 29 |
| Status COVID-19/Flu [d] | 76/76 | 99.90% (95.70%–99.96%) | EUA submission | 30 |
| VITROS [d] | 75/75 | 99.90% (95.45%–99.97%) | EUA submission | 31 |

[a] Anterior nasal swab
[b] Peer-reviewed EUA data
[c] Calculated based on both symptomatic and asymptomatic individuals
[d] Nasopharyngeal swab
[e] Mid-turbinate swab

**Table S8.** Cycle times in rapid antigen test false negatives and true positives

| Paired sampling outcome | Day of quarantine | Gene | | |
|---|---|---|---|---|
| | | *N* | ORF1ab | *S* |
| *RA − / RT-PCR +* | | | | |
| Sample 1 | 0 | 31 | 30 | 32 |
| Sample 2 | 0 | 32 | 31 | 30 |
| Sample 3 | 0 | 28 | 28 | 28 |
| Sample 4 | 0 | 30 | 31 | 36 |
| Sample 5 | 0 | 32 | 34 | —[a] |
| Sample 6 | 0 | 24 | 22 | 23 |
| Sample 7 | 0 | 32 | 31 | 35 |
| Sample 8 | 0 | 32 | 0[a] | 0[a] |
| Sample 9 | 0 | 30 | 33 | 34 |
| Sample 10 | 3 | 32 | 30 | —[a] |
| Sample 11 | 3 | 30 | 30 | 31 |
| Sample 12 | 3 | 29 | 28 | 28 |
| Sample 13 | 3 | —[a] | 31 | 33 |
| Sample 14 | 4 | 32 | 31 | 33 |
| Sample 15 | 4 | 27 | 25 | 26 |
| Sample 16 | 4 | 33 | —[a] | —[a] |
| *RA + / RT-PCR +* | | | | |
| Sample 1 | 0 | 17 | 17 | 18 |
| Sample 2 | 0 | —[a] | 34 | 34 |
| Sample 3 | 0 | 19 | 18 | 18 |
| Sample 4 | 3 | 13 | 11 | 12 |

[a] Result was negative (cycle time threshold: >36)

**Table S9.** Comparison of the internal and external percent positive agreement datasets for three rapid antigen tests

| Rapid Antigen Test | Log-likelihood | | | | Are the datasets significantly different |
|---|---|---|---|---|---|
| | Internal | External | Combined | Difference [a] | |
| BinaxNOW | - 83.64 | - 27.53 | - 124.62 | 13.45 | Yes |
| CareStart (AS) | - 14.88 | - 26.55 | - 41.49 | 0.05 | No |
| Sofia | - 4.38 | - 19.79 | - 26.72 | 2.54 | No |

[a] The difference may not reflect the difference based on values presented in the table due to rounding in the presentation of the log-likelihood values

**Table S10.** The estimated and 95% credible interval for the temporal RT-PCR diagnostic sensitivity curve

| Incubation period (days) | Functional form | Parameter determining time of peak sensitivity (*K*) | Parameter determining the shape of the curve (*z*) | Scaling parameter (*C*) | Peak diagnostic sensitivity |
|---|---|---|---|---|---|
| 4.4 [10] | log-Normal | 1.81<br>(1.69–2) | 0.822<br>(0.749–0.932) | 8.95<br>(7.2–10.5) | 1<br>(0.865–1) |
| 4.4 [10] | log-Student's *t* | 0.322<br>(0.322–0.322) | $9.99 \times 10^7$<br>($52.2–10^8$) | 2.18<br>(1.81–2.46) | 0.872<br>(0.692–0.991) |
| 5.72 [12] | log-Normal | 2.04<br>(1.97–2.13) | 0.581<br>(0.52–0.658) | 9.1<br>(7.52–10.4) | 0.964<br>(0.88–0.997) |

### Supplementary figures

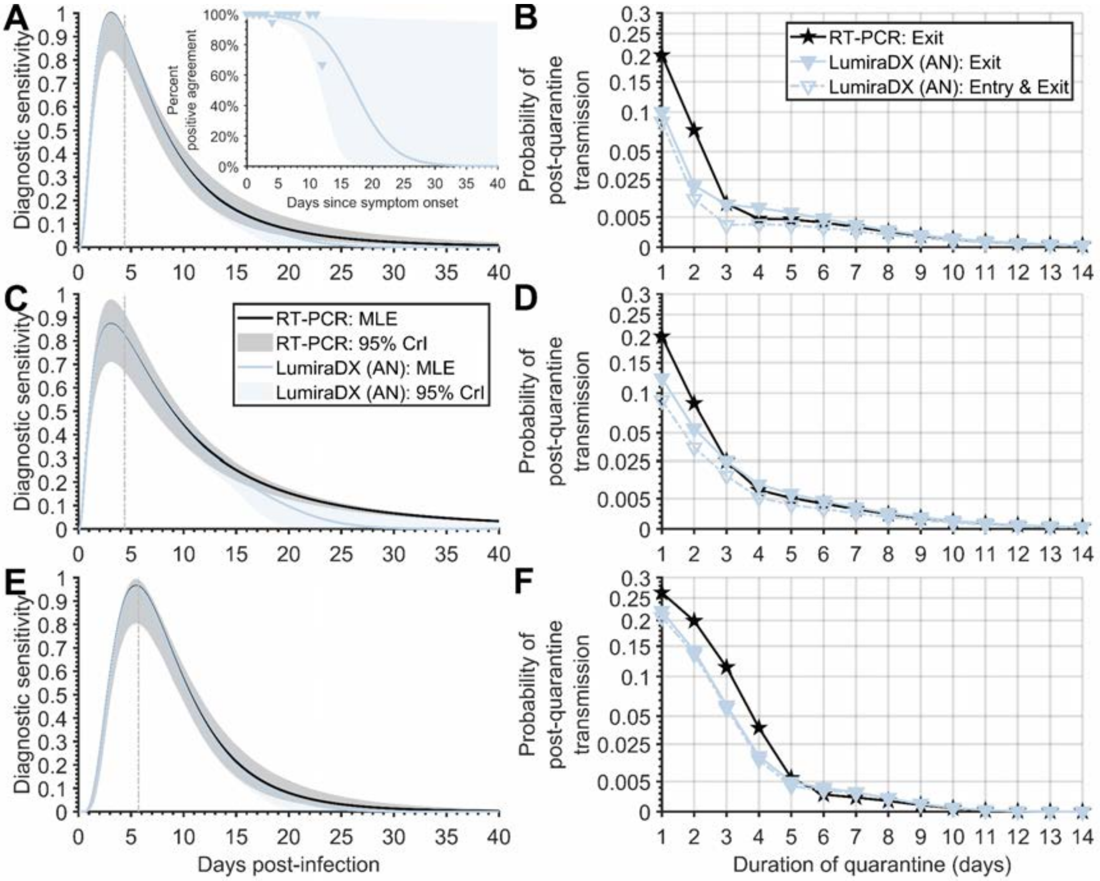


**Figure S1. The diagnostic sensitivity curve and probability of post-quarantine transmission for LumiraDx anterior nasal swab.** Specifying a 4.4-day incubation period, 35.1% of infections being asymptomatic, self-isolation upon symptom onset, (**A**) the RT-PCR diagnostic sensitivity curve (black) informed by data from Hellewell et al [9] using a log-Normal distribution and logistic regression model for the diagnostic sensitivity of the rapid antigen test (blue, solid line) informed by percent positive agreement data (blue triangles); (**B**) the probability of post-quarantine transmission for a RT-PCR test conducted 24 h before exit from quarantine (black stars), a rapid antigen test on exit (filled blue triangles), as well as a rapid antigen test conducted on both entry to and exit from quarantine (open blue triangles, dashed line); (**C**) the RT-PCR diagnostic sensitivity curve informed by data from Hellewell et al [9] using a log-Student's *t* distribution and logistic regression model for the diagnostic sensitivity of the rapid antigen test; (**D**) the probability of post-quarantine transmission for a RT-PCR test conducted 24 h before exit from quarantine, a rapid antigen test on exit, as well as a rapid antigen test conducted on both entry to and exit from quarantine. Specifying an 5.72-day incubation period, 35.1% of infections being asymptomatic, self-isolation upon symptom onset, (**E**) the RT-PCR diagnostic sensitivity curve informed by data from Hellewell et al [9] using a log-Normal distribution and logistic regression model for the diagnostic sensitivity of the rapid antigen test; and (**F**) the probability of post-quarantine transmission for a RT-PCR test conducted 24 h before exit from quarantine, a rapid antigen test on exit, as well as a rapid antigen test conducted on both entry to and exit from quarantine. The colored area denotes the 2.5 and 97.5 percentile constructed from 1,000 samples of the RT-PCR diagnostic sensitivity curve and the rapid antigen tests percent positive agreement curve.

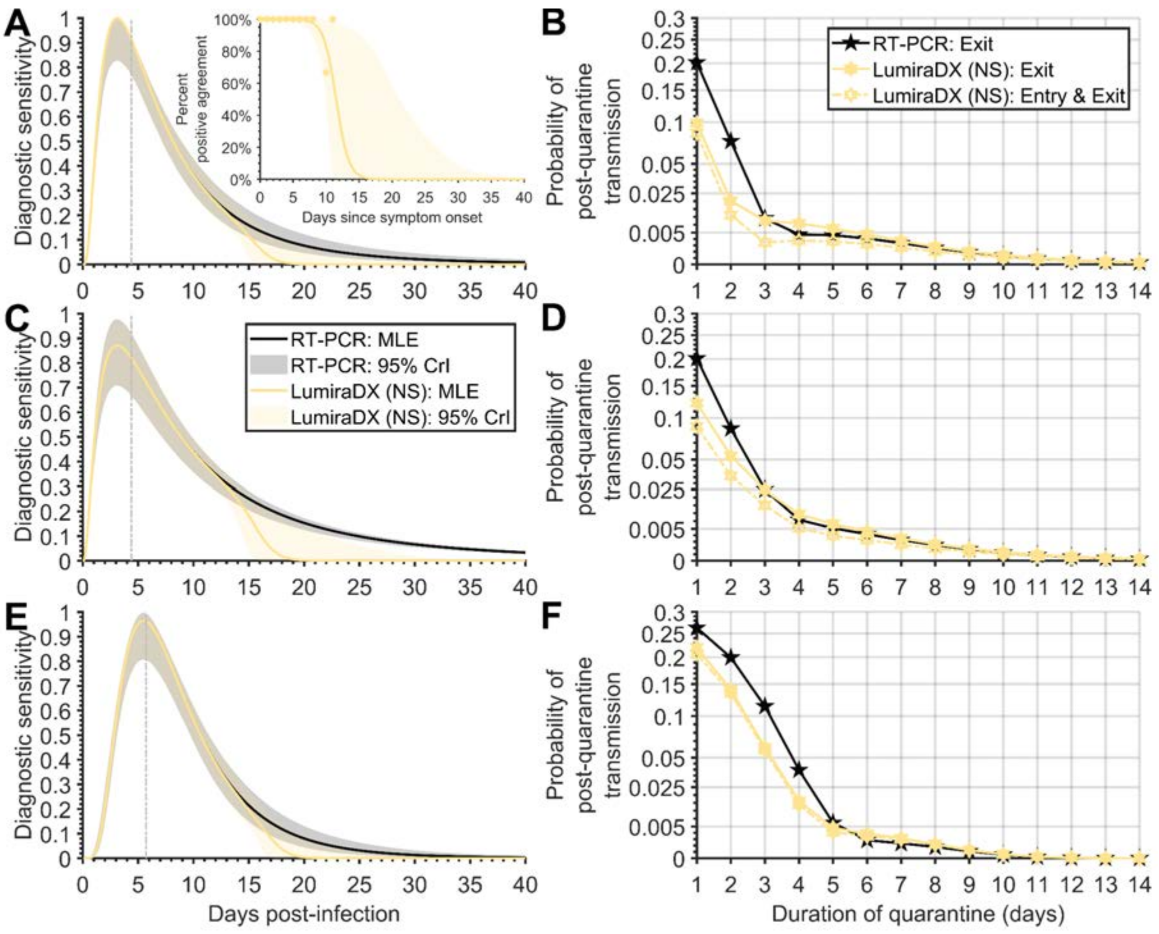


**Figure S2. The diagnostic sensitivity curve and probability of post-quarantine transmission for LumiraDx nasopharyngeal nasal swab.** Specifying a 4.4-day incubation period, 35.1% of infections being asymptomatic, self-isolation upon symptom onset, (**A**) the RT-PCR diagnostic sensitivity curve (black) informed by data from Hellewell et al [9] using a log-Normal distribution and logistic regression model for the diagnostic sensitivity of the rapid antigen test (yellow, solid line) informed by percent positive agreement data (yellow hexagrams); (**B**) the probability of post-quarantine transmission for a RT-PCR test conducted 24 h before exit from quarantine (black stars), a rapid antigen test on exit (filled yellow hexagrams), as well as a rapid antigen test conducted on both entry to and exit from quarantine (open yellow hexagrams, dashed line); (**C**) the RT-PCR diagnostic sensitivity curve informed by data from Hellewell et al [9] using a log-Student's *t* distribution and logistic regression model for the diagnostic sensitivity of the rapid antigen test; (**D**) the probability of post-quarantine transmission for a RT-PCR test conducted 24 h before exit from quarantine, a rapid antigen test on exit, as well as a rapid antigen test conducted on both entry to and exit from quarantine. Specifying an 5.72-day incubation period, 35.1% of infections being asymptomatic, self-isolation upon symptom onset, (**E**) the RT-PCR diagnostic sensitivity curve informed by data from Hellewell et al [9] using a log-Normal distribution and logistic regression model for the diagnostic sensitivity of the rapid antigen test; and (**F**) the probability of post-quarantine transmission for a RT-PCR test conducted 24 h before exit from quarantine, a rapid antigen test on exit, as well as a rapid antigen test conducted on both entry to and exit from quarantine. The colored area denotes the 2.5 and 97.5 percentile constructed from 1,000 samples of the RT-PCR diagnostic sensitivity curve and the rapid antigen tests percent positive agreement curve.

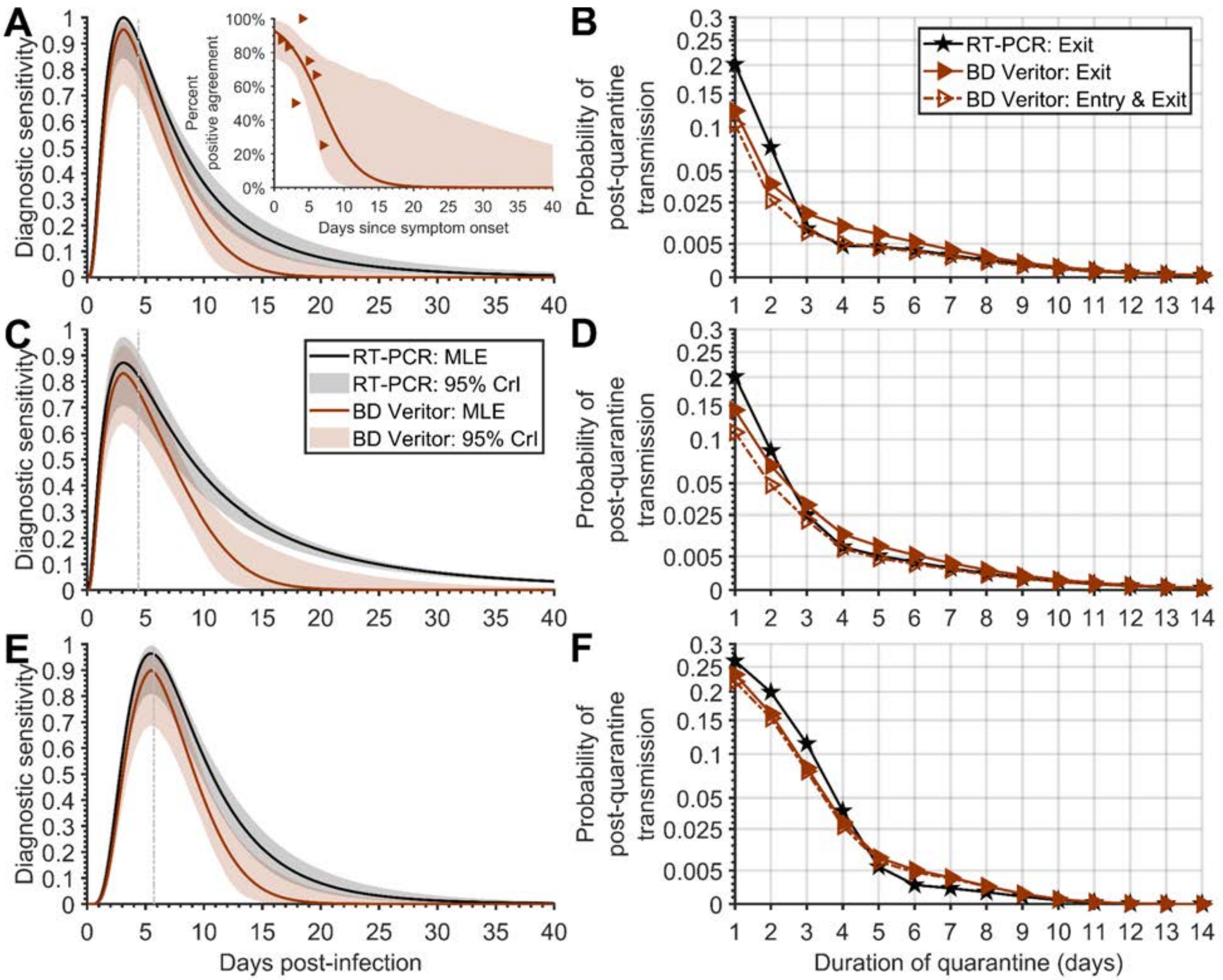


**Figure S3. The diagnostic sensitivity curve and probability of post-quarantine transmission for BD Veritor.** Specifying a 4.4-day incubation period, 35.1% of infections being asymptomatic, self-isolation upon symptom onset, (**A**) the RT-PCR diagnostic sensitivity curve (black) informed by data from Hellewell et al [9] using a log-Normal distribution and logistic regression model for the diagnostic sensitivity of the rapid antigen test (red, solid line) informed by percent positive agreement data (red triangles); (**B**) the probability of post-quarantine transmission for a RT-PCR test conducted 24 h before exit from quarantine (black stars), a rapid antigen test on exit (filled red triangles), as well as a rapid antigen test conducted on both entry to and exit from quarantine (open red triangles, dashed line); (**C**) the RT-PCR diagnostic sensitivity curve informed by data from Hellewell et al [9] using a log-Student's *t* distribution and logistic regression model for the diagnostic sensitivity of the rapid antigen test; (**D**) the probability of post-quarantine transmission for a RT-PCR test conducted 24 h before exit from quarantine, a rapid antigen test on exit, as well as a rapid antigen test conducted on both entry to and exit from quarantine. Specifying an 5.72-day incubation period, 35.1% of infections being asymptomatic, self-isolation upon symptom onset, (**E**) the RT-PCR diagnostic sensitivity curve informed by data from Hellewell et al [9] using a log-Normal distribution and logistic regression model for the diagnostic sensitivity of the rapid antigen test; and (**F**) the probability of post-quarantine transmission for a RT-PCR test conducted 24 h before exit from quarantine, a rapid antigen test on exit, as well as a rapid antigen test conducted on both entry to and exit from quarantine. The colored area denotes the 2.5 and 97.5 percentile constructed from 1,000 samples of the RT-PCR diagnostic sensitivity curve and the rapid antigen tests percent positive agreement curve.

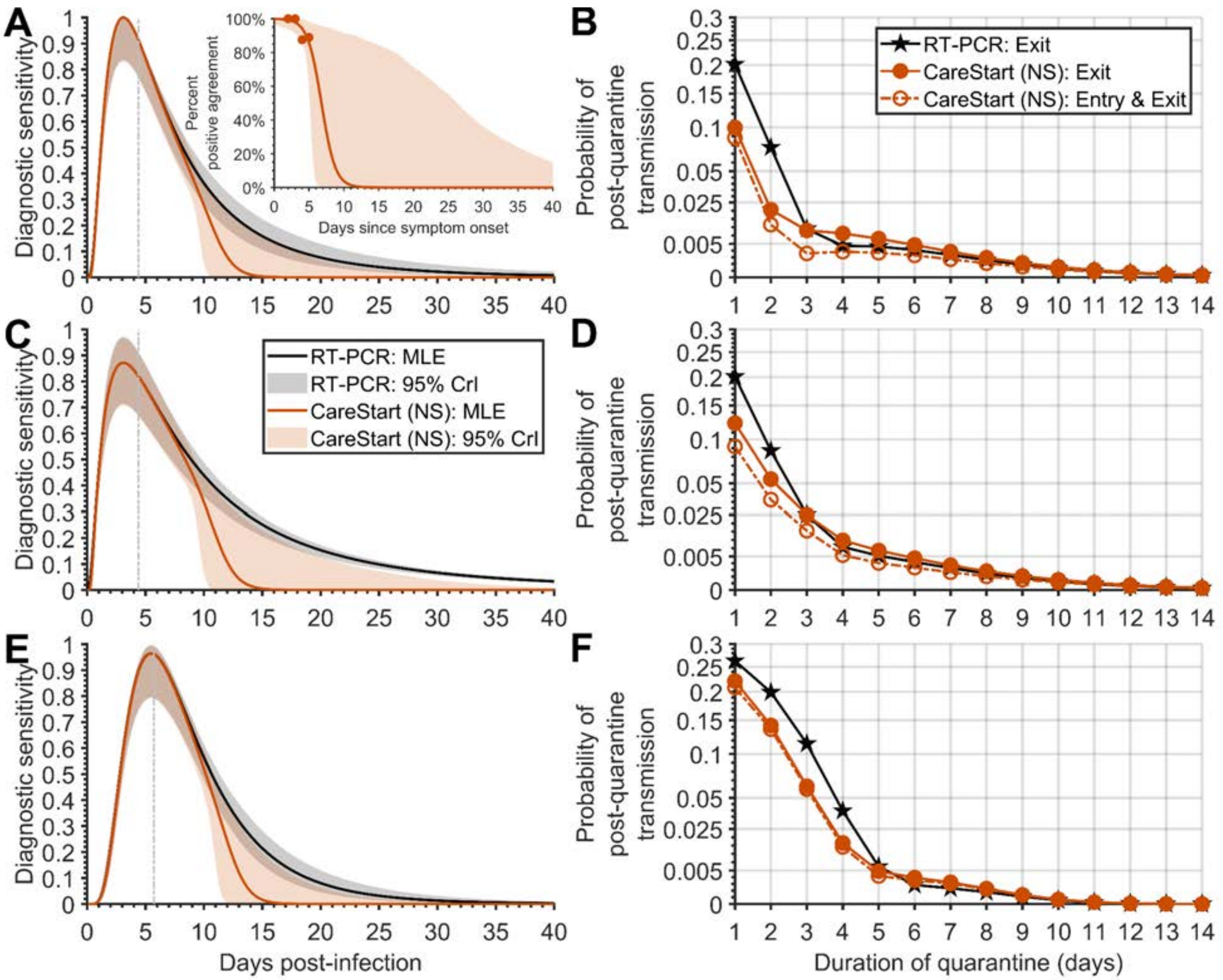


**Figure S4. The diagnostic sensitivity curve and probability of post-quarantine transmission for CareStart nasopharyngeal nasal swab.** Specifying a 4.4-day incubation period, 35.1% of infections being asymptomatic, self-isolation upon symptom onset, (**A**) the RT-PCR diagnostic sensitivity curve (black) informed by data from Hellewell et al [9] using a log-Normal distribution and logistic regression model for the diagnostic sensitivity of the rapid antigen test (orange, solid line) informed by percent positive agreement data (orange circles); (**B**) the probability of post-quarantine transmission for a RT-PCR test conducted 24 h before exit from quarantine (black stars), a rapid antigen test on exit (filled orange circles), as well as a rapid antigen test conducted on both entry to and exit from quarantine (open orange circles, dashed line); (**C**) the RT-PCR diagnostic sensitivity curve informed by data from Hellewell et al [9] using a log-Student's *t* distribution and logistic regression model for the diagnostic sensitivity of the rapid antigen test; (**D**) the probability of post-quarantine transmission for a RT-PCR test conducted 24 h before exit from quarantine, a rapid antigen test on exit, as well as a rapid antigen test conducted on both entry to and exit from quarantine. Specifying an 5.72-day incubation period, 35.1% of infections being asymptomatic, self-isolation upon symptom onset, (**E**) the RT-PCR diagnostic sensitivity curve informed by data from Hellewell et al [9] using a log-Normal distribution and logistic regression model for the diagnostic sensitivity of the rapid antigen test; and (**F**) the probability of post-quarantine transmission for a RT-PCR test conducted 24 h before exit from quarantine, a rapid antigen test on exit, as well as a rapid antigen test conducted on both entry to and exit from quarantine. The colored area denotes the 2.5 and 97.5 percentile constructed from 1,000 samples of the RT-PCR diagnostic sensitivity curve and the rapid antigen tests percent positive agreement curve.

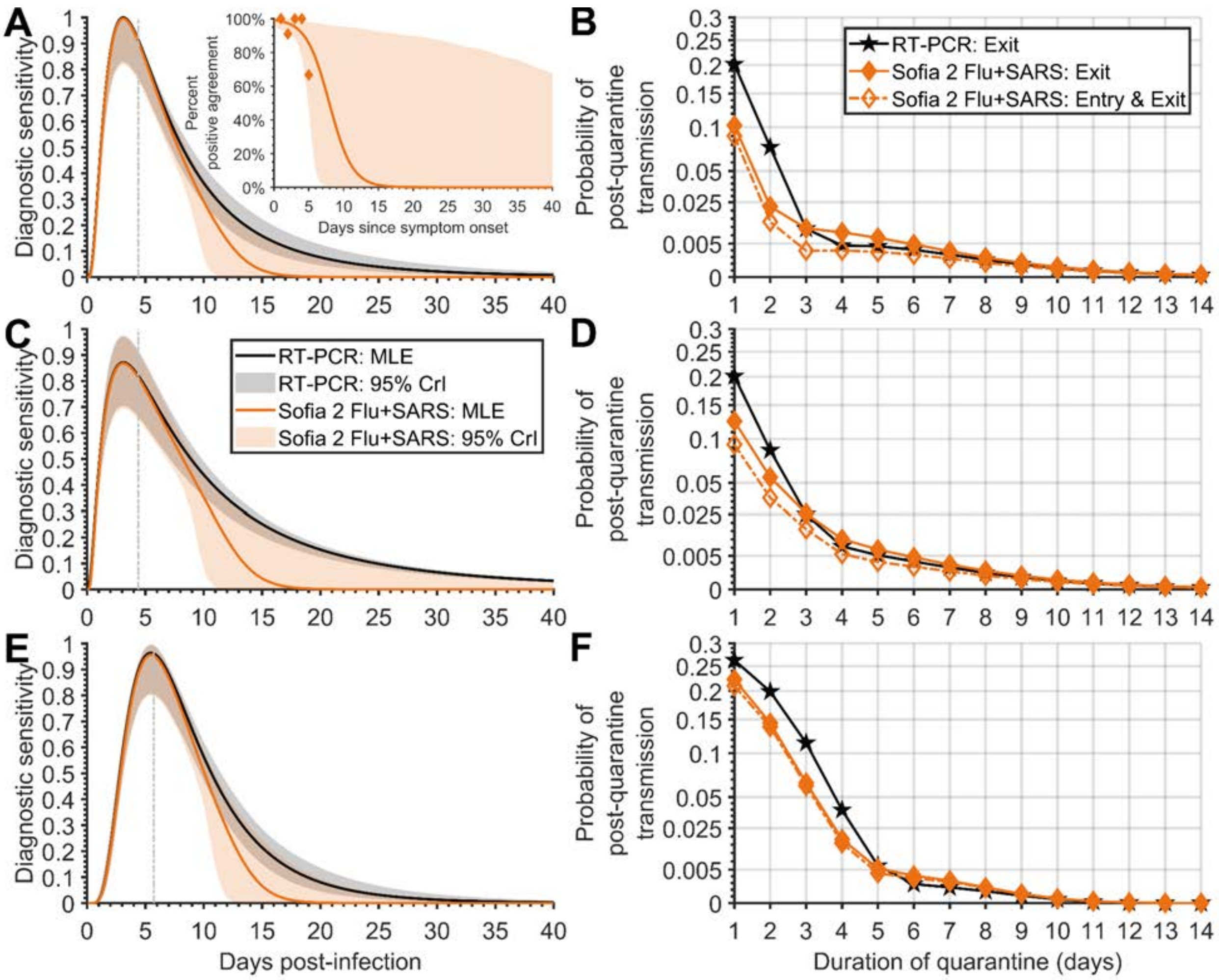


**Figure S5. The diagnostic sensitivity curve and probability of post-quarantine transmission for Sofia 2 Flu + SARS.** Specifying a 4.4-day incubation period, 35.1% of infections being asymptomatic, self-isolation upon symptom onset, (**A**) the RT-PCR diagnostic sensitivity curve (black) informed by data from Hellewell et al [9] using a log-Normal distribution and logistic regression model for the diagnostic sensitivity of the rapid antigen test (orange, solid line) informed by percent positive agreement data (orange diamonds); (**B**) the probability of post-quarantine transmission for a RT-PCR test conducted 24 h before exit from quarantine (black stars), a rapid antigen test on exit (filled orange diamonds), as well as a rapid antigen test conducted on both entry to and exit from quarantine (open orange diamonds, dashed line); (**C**) the RT-PCR diagnostic sensitivity curve informed by data from Hellewell et al [9] using a log-Student's *t* distribution and logistic regression model for the diagnostic sensitivity of the rapid antigen test; (**D**) the probability of post-quarantine transmission for a RT-PCR test conducted 24 h before exit from quarantine, a rapid antigen test on exit, as well as a rapid antigen test conducted on both entry to and exit from quarantine. Specifying an 5.72-day incubation period, 35.1% of infections being asymptomatic, self-isolation upon symptom onset, (**E**) the RT-PCR diagnostic sensitivity curve informed by data from Hellewell et al [9] using a log-Normal distribution and logistic regression model for the diagnostic sensitivity of the rapid antigen test; and (**F**) the probability of post-quarantine transmission for a RT-PCR test conducted 24 h before exit from quarantine, a rapid antigen test on exit, as well as a rapid antigen test conducted on both entry to and exit from quarantine. The colored area denotes the 2.5 and 97.5 percentile constructed from 1,000 samples of the RT-PCR diagnostic sensitivity curve and the rapid antigen tests percent positive agreement curve.

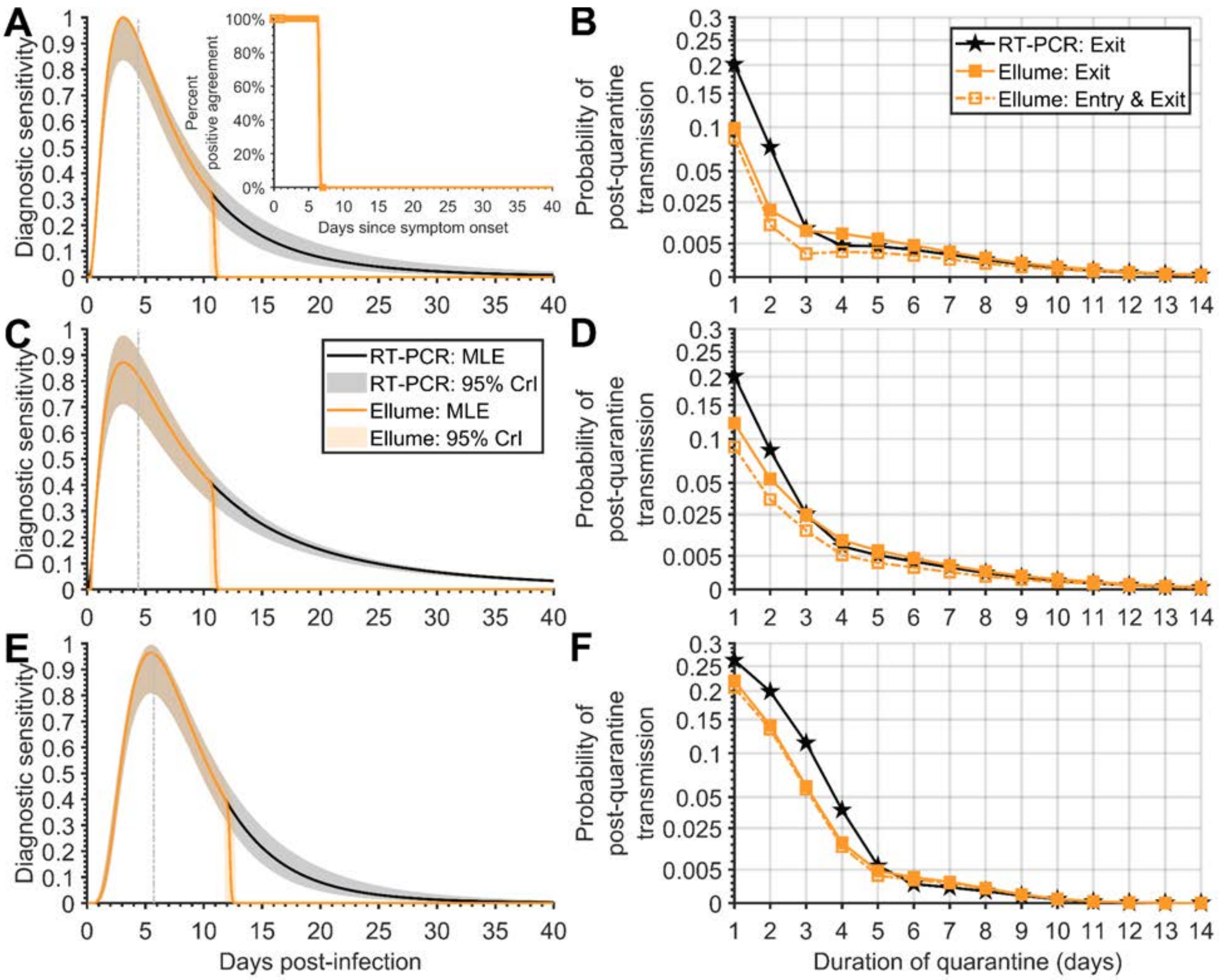


**Figure S6. The diagnostic sensitivity curve and probability of post-quarantine transmission for Ellume.** Specifying a 4.4-day incubation period, 35.1% of infections being asymptomatic, self-isolation upon symptom onset, (**A**) the RT-PCR diagnostic sensitivity curve (black) informed by data from Hellewell et al [9] using a log-Normal distribution and logistic regression model for the diagnostic sensitivity of the rapid antigen test (orange, solid line) informed by percent positive agreement data (orange squares); (**B**) the probability of post-quarantine transmission for a RT-PCR test conducted 24 h before exit from quarantine (black stars), a rapid antigen test on exit (filled orange squares), as well as a rapid antigen test conducted on both entry to and exit from quarantine (open orange squares, dashed line); (**C**) the RT-PCR diagnostic sensitivity curve informed by data from Hellewell et al [9] using a log-Student's *t* distribution and logistic regression model for the diagnostic sensitivity of the rapid antigen test; (**D**) the probability of post-quarantine transmission for a RT-PCR test conducted 24 h before exit from quarantine, a rapid antigen test on exit, as well as a rapid antigen test conducted on both entry to and exit from quarantine. Specifying an 5.72-day incubation period, 35.1% of infections being asymptomatic, self-isolation upon symptom onset, (**E**) the RT-PCR diagnostic sensitivity curve informed by data from Hellewell et al [9] using a log-Normal distribution and logistic regression model for the diagnostic sensitivity of the rapid antigen test; and (**F**) the probability of post-quarantine transmission for a RT-PCR test conducted 24 h before exit from quarantine, a rapid antigen test on exit, as well as a rapid antigen test conducted on both entry to and exit from quarantine. The colored area denotes the 2.5 and 97.5 percentile constructed from 1,000 samples of the RT-PCR diagnostic sensitivity curve and the rapid antigen tests percent positive agreement curve.

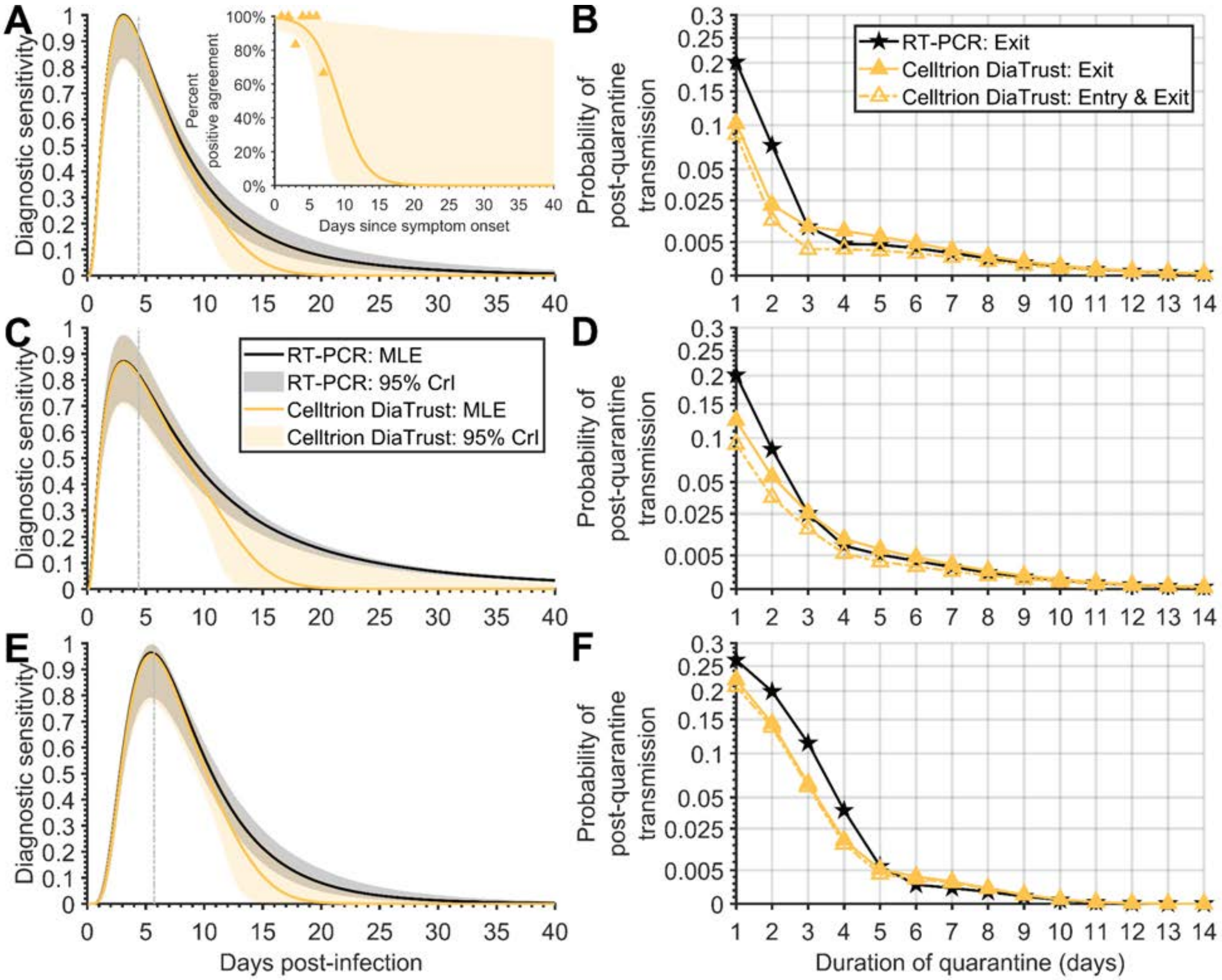


**Figure S7. The diagnostic sensitivity curve and probability of post-quarantine transmission for Celltrion DiaTrust.** Specifying a 4.4-day incubation period, 35.1% of infections being asymptomatic, self-isolation upon symptom onset, (**A**) the RT-PCR diagnostic sensitivity curve (black) informed by data from Hellewell et al [9] using a log-Normal distribution and logistic regression model for the diagnostic sensitivity of the rapid antigen test (yellow, solid line) informed by percent positive agreement data (yellow triangles); (**B**) the probability of post-quarantine transmission for a RT-PCR test conducted 24 h before exit from quarantine (black stars), a rapid antigen test on exit (filled yellow triangles), as well as a rapid antigen test conducted on both entry to and exit from quarantine (open yellow triangles, dashed line); (**C**) the RT-PCR diagnostic sensitivity curve informed by data from Hellewell et al [9] using a log-Student's *t* distribution and logistic regression model for the diagnostic sensitivity of the rapid antigen test; (**D**) the probability of post-quarantine transmission for a RT-PCR test conducted 24 h before exit from quarantine, a rapid antigen test on exit, as well as a rapid antigen test conducted on both entry to and exit from quarantine. Specifying an 5.72-day incubation period, 35.1% of infections being asymptomatic, self-isolation upon symptom onset, (**E**) the RT-PCR diagnostic sensitivity curve informed by data from Hellewell et al [9] using a log-Normal distribution and logistic regression model for the diagnostic sensitivity of the rapid antigen test; and (**F**) the probability of post-quarantine transmission for a RT-PCR test conducted 24 h before exit from quarantine, a rapid antigen test on exit, as well as a rapid antigen test conducted on both entry to and exit from quarantine. The colored area denotes the 2.5 and 97.5 percentile constructed from 1,000 samples of the RT-PCR diagnostic sensitivity curve and the rapid antigen tests percent positive agreement curve.

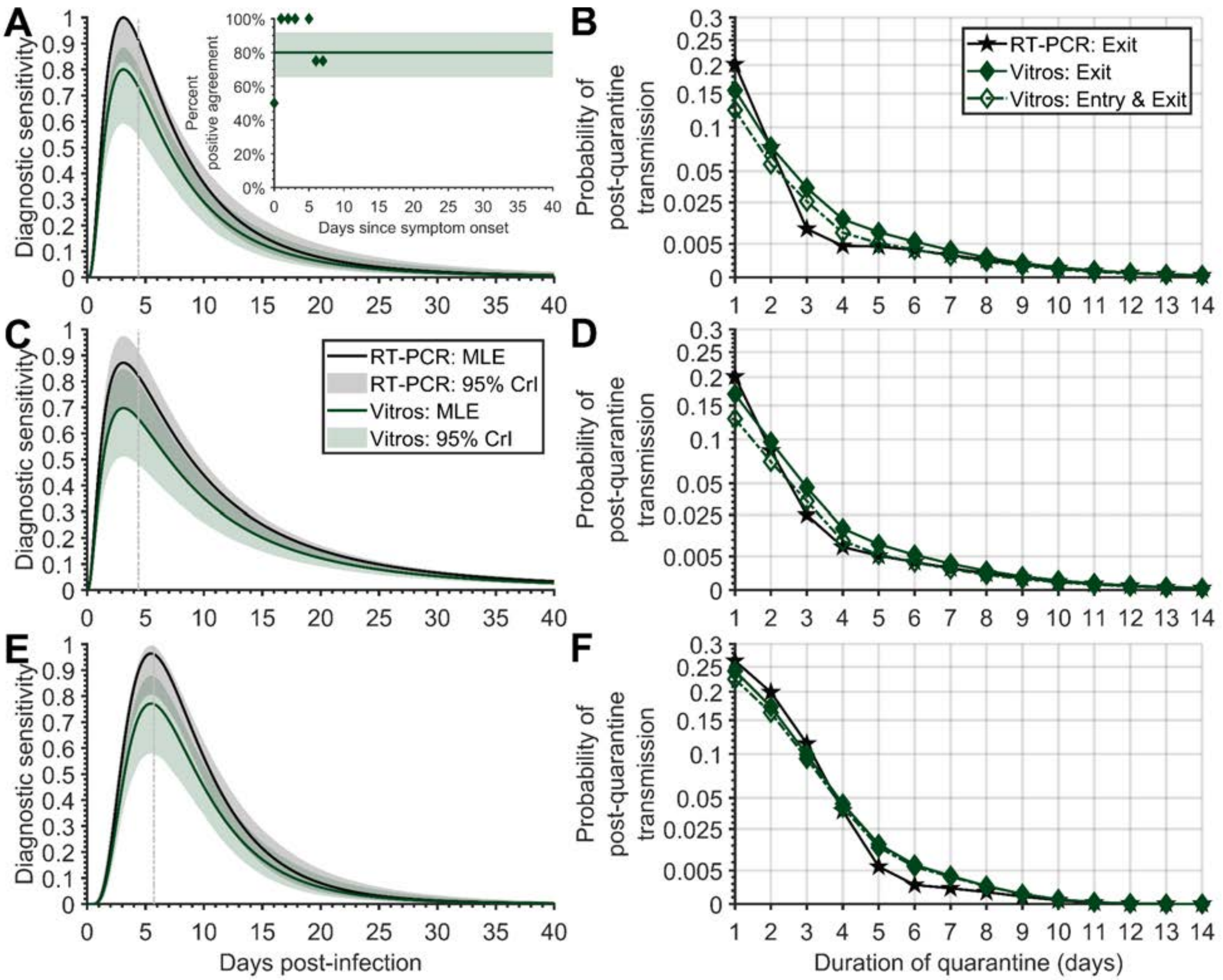


**Figure S8. The diagnostic sensitivity curve and probability of post-quarantine transmission for Vitros.** Specifying a 4.4-day incubation period, 35.1% of infections being asymptomatic, self-isolation upon symptom onset, (**A**) the RT-PCR diagnostic sensitivity curve (black) informed by data from Hellewell et al [9] using a log-Normal distribution and logistic regression model for the diagnostic sensitivity of the rapid antigen test (green, solid line) informed by percent positive agreement data (green diamonds); (**B**) the probability of post-quarantine transmission for a RT-PCR test conducted 24 h before exit from quarantine (black stars), a rapid antigen test on exit (filled green diamonds), as well as a rapid antigen test conducted on both entry to and exit from quarantine (open green diamonds, dashed line); (**C**) the RT-PCR diagnostic sensitivity curve informed by data from Hellewell et al [9] using a log-Student's *t* distribution and logistic regression model for the diagnostic sensitivity of the rapid antigen test; (**D**) the probability of post-quarantine transmission for a RT-PCR test conducted 24 h before exit from quarantine, a rapid antigen test on exit, as well as a rapid antigen test conducted on both entry to and exit from quarantine. Specifying an 5.72-day incubation period, 35.1% of infections being asymptomatic, self-isolation upon symptom onset, (**E**) the RT-PCR diagnostic sensitivity curve informed by data from Hellewell et al [9] using a log-Normal distribution and logistic regression model for the diagnostic sensitivity of the rapid antigen test; and (**F**) the probability of post-quarantine transmission for a RT-PCR test conducted 24 h before exit from quarantine, a rapid antigen test on exit, as well as a rapid antigen test conducted on both entry to and exit from quarantine. The colored area denotes the 2.5 and 97.5 percentile constructed from 1,000 samples of the RT-PCR diagnostic sensitivity curve and the rapid antigen tests percent positive agreement curve.

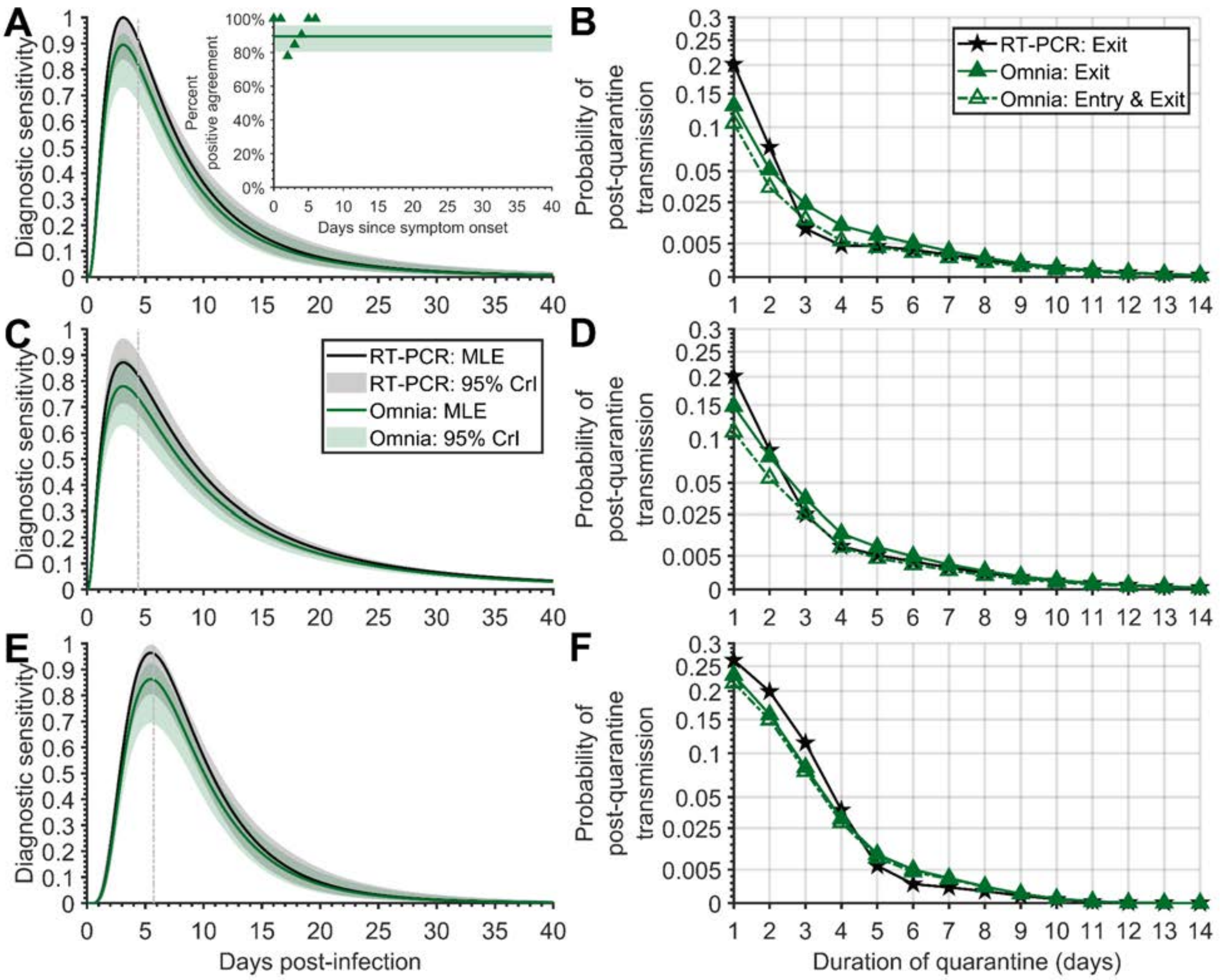


**Figure S9. The diagnostic sensitivity curve and probability of post-quarantine transmission for Omnia.** Specifying a 4.4-day incubation period, 35.1% of infections being asymptomatic, self-isolation upon symptom onset, (**A**) the RT-PCR diagnostic sensitivity curve (black) informed by data from Hellewell et al [9] using a log-Normal distribution and logistic regression model for the diagnostic sensitivity of the rapid antigen test (green, solid line) informed by percent positive agreement data (green triangles); (**B**) the probability of post-quarantine transmission for a RT-PCR test conducted 24 h before exit from quarantine (black stars), a rapid antigen test on exit (filled green triangles), as well as a rapid antigen test conducted on both entry to and exit from quarantine (open green triangles, dashed line); (**C**) the RT-PCR diagnostic sensitivity curve informed by data from Hellewell et al [9] using a log-Student's *t* distribution and logistic regression model for the diagnostic sensitivity of the rapid antigen test; (**D**) the probability of post-quarantine transmission for a RT-PCR test conducted 24 h before exit from quarantine, a rapid antigen test on exit, as well as a rapid antigen test conducted on both entry to and exit from quarantine. Specifying an 5.72-day incubation period, 35.1% of infections being asymptomatic, self-isolation upon symptom onset, (**E**) the RT-PCR diagnostic sensitivity curve informed by data from Hellewell et al [9] using a log-Normal distribution and logistic regression model for the diagnostic sensitivity of the rapid antigen test; and (**F**) the probability of post-quarantine transmission for a RT-PCR test conducted 24 h before exit from quarantine, a rapid antigen test on exit, as well as a rapid antigen test conducted on both entry to and exit from quarantine. The colored area denotes the 2.5 and 97.5 percentile constructed from 1,000 samples of the RT-PCR diagnostic sensitivity curve and the rapid antigen tests percent positive agreement curve.

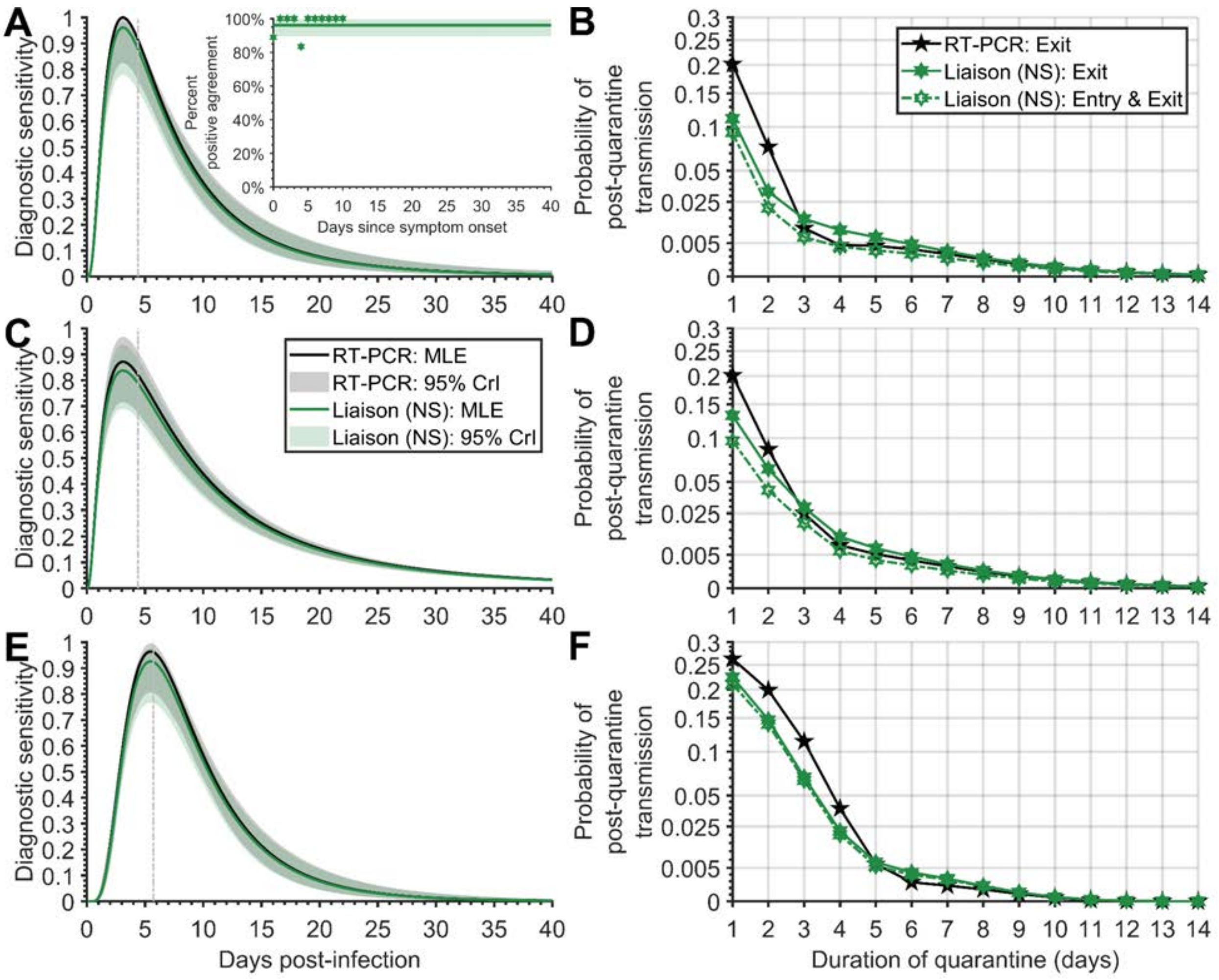


**Figure S10. The diagnostic sensitivity curve and probability of post-quarantine transmission for Liaison nasopharyngeal nasal swab.** Specifying a 4.4-day incubation period, 35.1% of infections being asymptomatic, self-isolation upon symptom onset, (**A**) the RT-PCR diagnostic sensitivity curve (black) informed by data from Hellewell et al [9] using a log-Normal distribution and logistic regression model for the diagnostic sensitivity of the rapid antigen test (green, solid line) informed by percent positive agreement data (green hexagrams); (**B**) the probability of post-quarantine transmission for a RT-PCR test conducted 24 h before exit from quarantine (black stars), a rapid antigen test on exit (filled green hexagrams), as well as a rapid antigen test conducted on both entry to and exit from quarantine (open green hexagrams, dashed line); (**C**) the RT-PCR diagnostic sensitivity curve informed by data from Hellewell et al [9] using a log-Student's *t* distribution and logistic regression model for the diagnostic sensitivity of the rapid antigen test; (**D**) the probability of post-quarantine transmission for a RT-PCR test conducted 24 h before exit from quarantine, a rapid antigen test on exit, as well as a rapid antigen test conducted on both entry to and exit from quarantine. Specifying an 5.72-day incubation period, 35.1% of infections being asymptomatic, self-isolation upon symptom onset, (**E**) the RT-PCR diagnostic sensitivity curve informed by data from Hellewell et al [9] using a log-Normal distribution and logistic regression model for the diagnostic sensitivity of the rapid antigen test; and (**F**) the probability of post-quarantine transmission for a RT-PCR test conducted 24 h before exit from quarantine, a rapid antigen test on exit, as well as a rapid antigen test conducted on both entry to and exit from quarantine. The colored area denotes the 2.5 and 97.5 percentile constructed from 1,000 samples of the RT-PCR diagnostic sensitivity curve and the rapid antigen tests percent positive agreement curve.

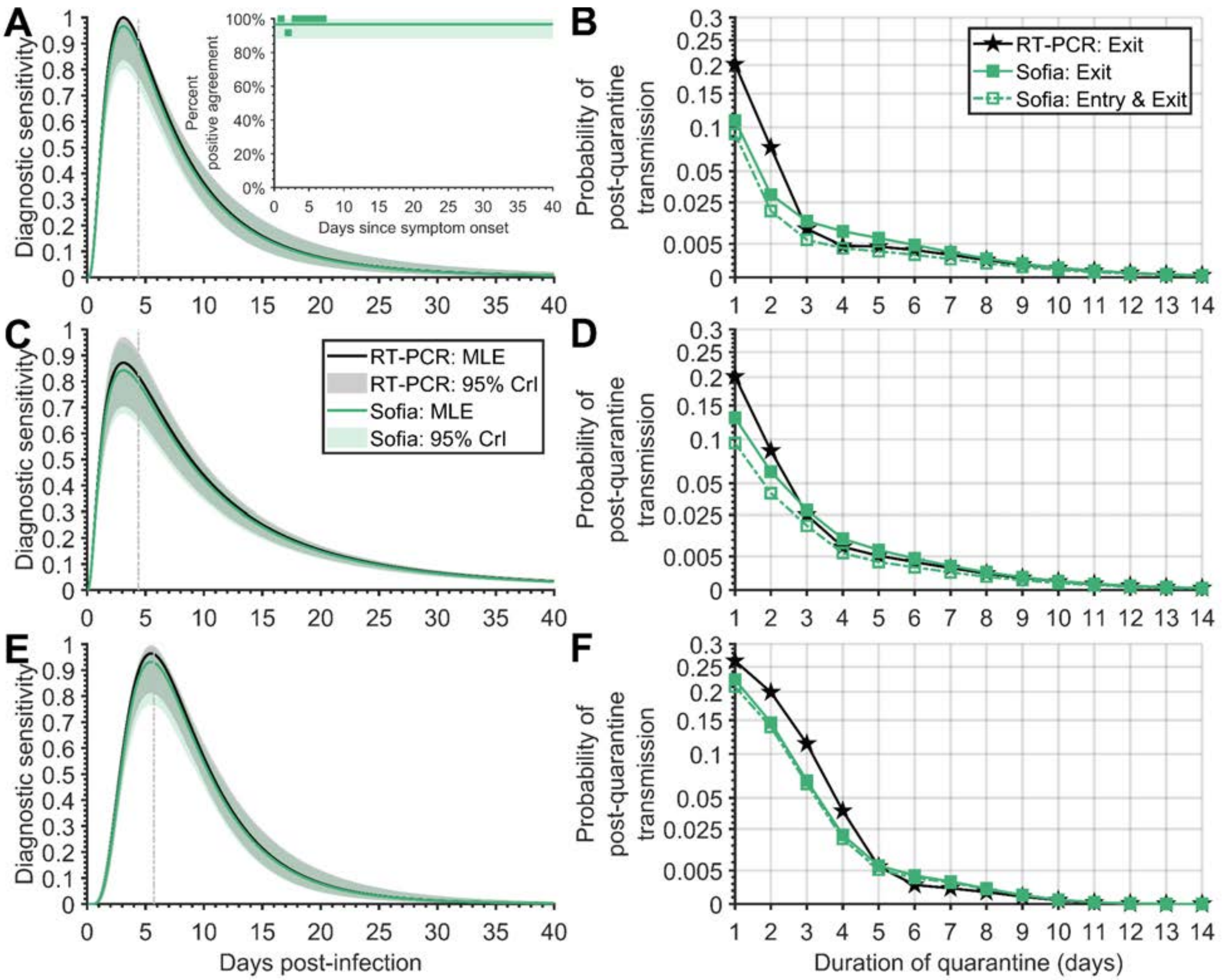


**Figure S11. The diagnostic sensitivity curve and probability of post-quarantine transmission for Sofia.** Specifying a 4.4-day incubation period, 35.1% of infections being asymptomatic, self-isolation upon symptom onset, (**A**) the RT-PCR diagnostic sensitivity curve (black) informed by data from Hellewell et al [9] using a log-Normal distribution and logistic regression model for the diagnostic sensitivity of the rapid antigen test (green, solid line) informed by percent positive agreement data (green squares); (**B**) the probability of post-quarantine transmission for a RT-PCR test conducted 24 h before exit from quarantine (black stars), a rapid antigen test on exit (filled green squares), as well as a rapid antigen test conducted on both entry to and exit from quarantine (open green squares, dashed line); (**C**) the RT-PCR diagnostic sensitivity curve informed by data from Hellewell et al [9] using a log-Student's *t* distribution and logistic regression model for the diagnostic sensitivity of the rapid antigen test; (**D**) the probability of post-quarantine transmission for a RT-PCR test conducted 24 h before exit from quarantine, a rapid antigen test on exit, as well as a rapid antigen test conducted on both entry to and exit from quarantine. Specifying an 5.72-day incubation period, 35.1% of infections being asymptomatic, self-isolation upon symptom onset, (**E**) the RT-PCR diagnostic sensitivity curve informed by data from Hellewell et al [9] using a log-Normal distribution and logistic regression model for the diagnostic sensitivity of the rapid antigen test; and (**F**) the probability of post-quarantine transmission for a RT-PCR test conducted 24 h before exit from quarantine, a rapid antigen test on exit, as well as a rapid antigen test conducted on both entry to and exit from quarantine. The colored area denotes the 2.5 and 97.5 percentile constructed from 1,000 samples of the RT-PCR diagnostic sensitivity curve and the rapid antigen tests percent positive agreement curve.

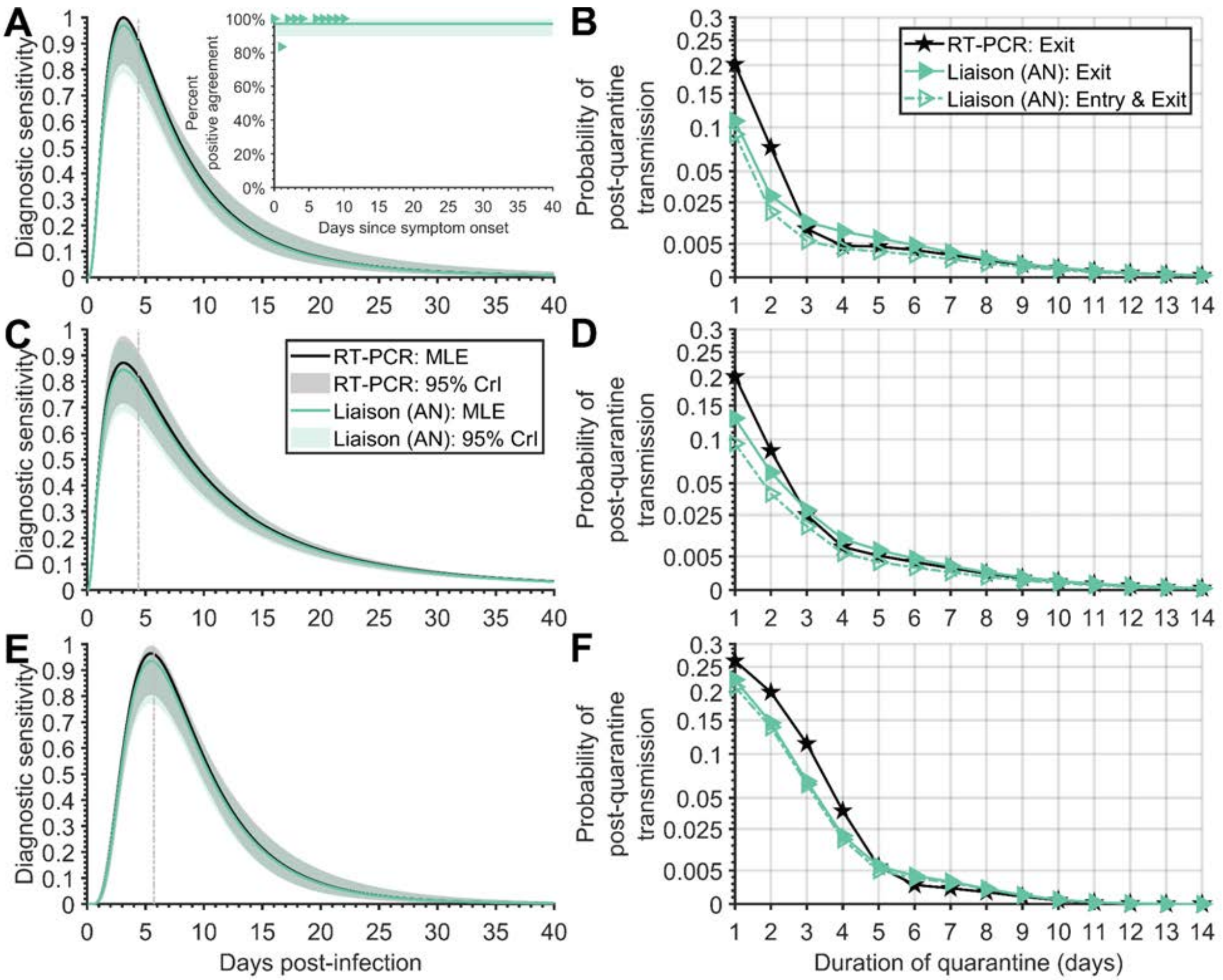


**Figure S12. The diagnostic sensitivity curve and probability of post-quarantine transmission for Liaison anterior nasal swab.** Specifying a 4.4-day incubation period, 35.1% of infections being asymptomatic, self-isolation upon symptom onset, (**A**) the RT-PCR diagnostic sensitivity curve (black) informed by data from Hellewell et al [9] using a log-Normal distribution and logistic regression model for the diagnostic sensitivity of the rapid antigen test (green, solid line) informed by percent positive agreement data (green triangles); (**B**) the probability of post-quarantine transmission for a RT-PCR test conducted 24 h before exit from quarantine (black stars), a rapid antigen test on exit (filled green triangles), as well as a rapid antigen test conducted on both entry to and exit from quarantine (open green triangles, dashed line); (**C**) the RT-PCR diagnostic sensitivity curve informed by data from Hellewell et al [9] using a log-Student's *t* distribution and logistic regression model for the diagnostic sensitivity of the rapid antigen test; (**D**) the probability of post-quarantine transmission for a RT-PCR test conducted 24 h before exit from quarantine, a rapid antigen test on exit, as well as a rapid antigen test conducted on both entry to and exit from quarantine. Specifying an 5.72-day incubation period, 35.1% of infections being asymptomatic, self-isolation upon symptom onset, (**E**) the RT-PCR diagnostic sensitivity curve informed by data from Hellewell et al [9] using a log-Normal distribution and logistic regression model for the diagnostic sensitivity of the rapid antigen test; and (**F**) the probability of post-quarantine transmission for a RT-PCR test conducted 24 h before exit from quarantine, a rapid antigen test on exit, as well as a rapid antigen test conducted on both entry to and exit from quarantine. The colored area denotes the 2.5 and 97.5 percentile constructed from 1,000 samples of the RT-PCR diagnostic sensitivity curve and the rapid antigen tests percent positive agreement curve.

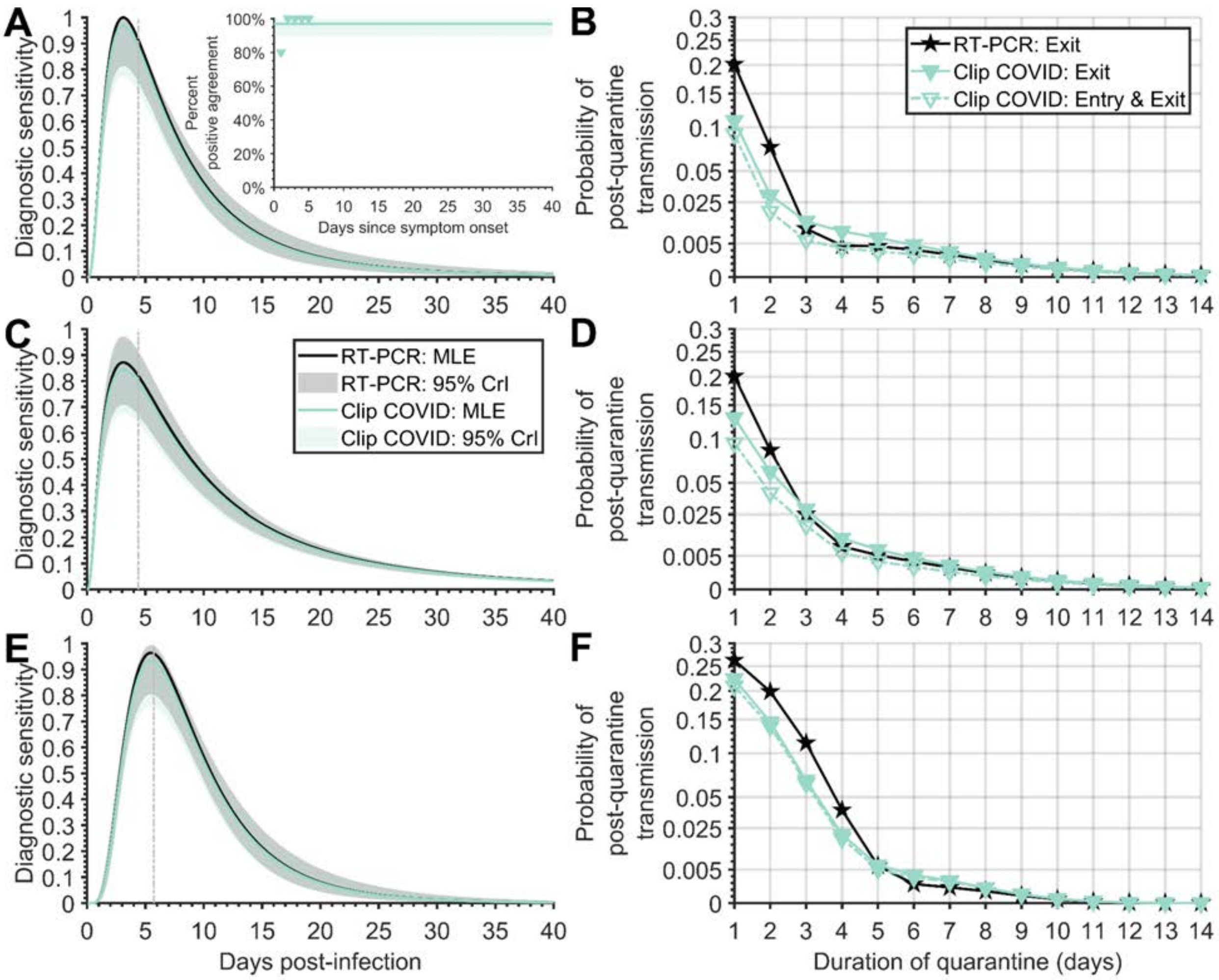


**Figure S13. The diagnostic sensitivity curve and probability of post-quarantine transmission for Clip COVID.** Specifying a 4.4-day incubation period, 35.1% of infections being asymptomatic, self-isolation upon symptom onset, (**A**) the RT-PCR diagnostic sensitivity curve (black) informed by data from Hellewell et al [9] using a log-Normal distribution and logistic regression model for the diagnostic sensitivity of the rapid antigen test (green, solid line) informed by percent positive agreement data (green triangles); (**B**) the probability of post-quarantine transmission for a RT-PCR test conducted 24 h before exit from quarantine (black stars), a rapid antigen test on exit (filled green triangles), as well as a rapid antigen test conducted on both entry to and exit from quarantine (open green triangles, dashed line); (**C**) the RT-PCR diagnostic sensitivity curve informed by data from Hellewell et al [9] using a log-Student's *t* distribution and logistic regression model for the diagnostic sensitivity of the rapid antigen test; (**D**) the probability of post-quarantine transmission for a RT-PCR test conducted 24 h before exit from quarantine, a rapid antigen test on exit, as well as a rapid antigen test conducted on both entry to and exit from quarantine. Specifying an 5.72-day incubation period, 35.1% of infections being asymptomatic, self-isolation upon symptom onset, (**E**) the RT-PCR diagnostic sensitivity curve informed by data from Hellewell et al [9] using a log-Normal distribution and logistic regression model for the diagnostic sensitivity of the rapid antigen test; and (**F**) the probability of post-quarantine transmission for a RT-PCR test conducted 24 h before exit from quarantine, a rapid antigen test on exit, as well as a rapid antigen test conducted on both entry to and exit from quarantine. The colored area denotes the 2.5 and 97.5 percentile constructed from 1,000 samples of the RT-PCR diagnostic sensitivity curve and the rapid antigen tests percent positive agreement curve.

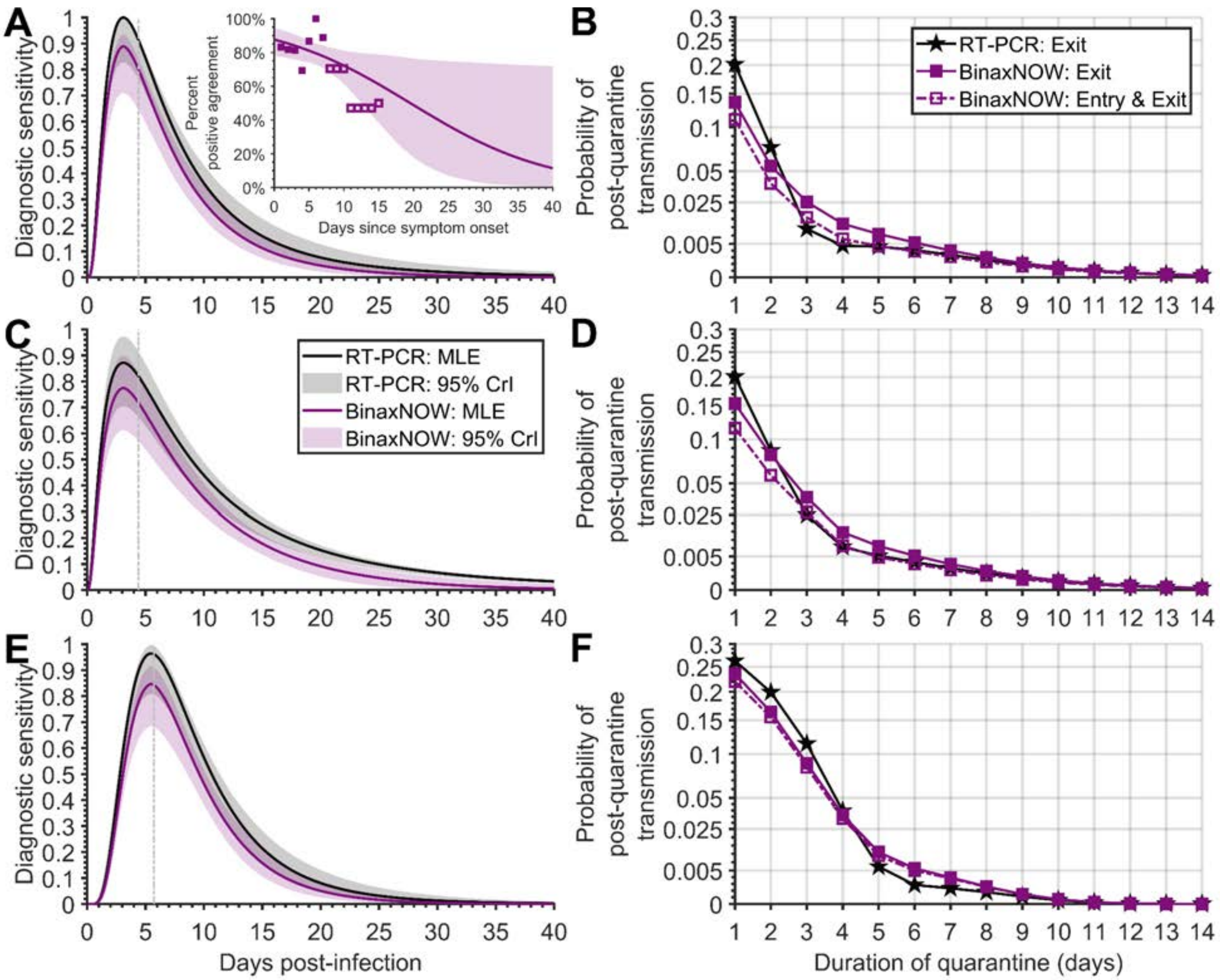


**Figure S14. The diagnostic sensitivity curve and probability of post-quarantine transmission for BinaxNOW.** Specifying a 4.4-day incubation period, 35.1% of infections being asymptomatic, self-isolation upon symptom onset, (**A**) the RT-PCR diagnostic sensitivity curve (black) informed by data from Hellewell et al [9] using a log-Normal distribution and logistic regression model for the diagnostic sensitivity of the rapid antigen test (purple, solid line) informed by percent positive agreement data (purple squares); (**B**) the probability of post-quarantine transmission for a RT-PCR test conducted 24 h before exit from quarantine (black stars), a rapid antigen test on exit (filled purple squares), as well as a rapid antigen test conducted on both entry to and exit from quarantine (open purple squares, dashed line); (**C**) the RT-PCR diagnostic sensitivity curve informed by data from Hellewell et al [9] using a log-Student's *t* distribution and logistic regression model for the diagnostic sensitivity of the rapid antigen test; (**D**) the probability of post-quarantine transmission for a RT-PCR test conducted 24 h before exit from quarantine, a rapid antigen test on exit, as well as a rapid antigen test conducted on both entry to and exit from quarantine. Specifying an 5.72-day incubation period, 35.1% of infections being asymptomatic, self-isolation upon symptom onset, (**E**) the RT-PCR diagnostic sensitivity curve informed by data from Hellewell et al [9] using a log-Normal distribution and logistic regression model for the diagnostic sensitivity of the rapid antigen test; and (**F**) the probability of post-quarantine transmission for a RT-PCR test conducted 24 h before exit from quarantine, a rapid antigen test on exit, as well as a rapid antigen test conducted on both entry to and exit from quarantine. The colored area denotes the 2.5 and 97.5 percentile constructed from 1,000 samples of the RT-PCR diagnostic sensitivity curve and the rapid antigen tests percent positive agreement curve.

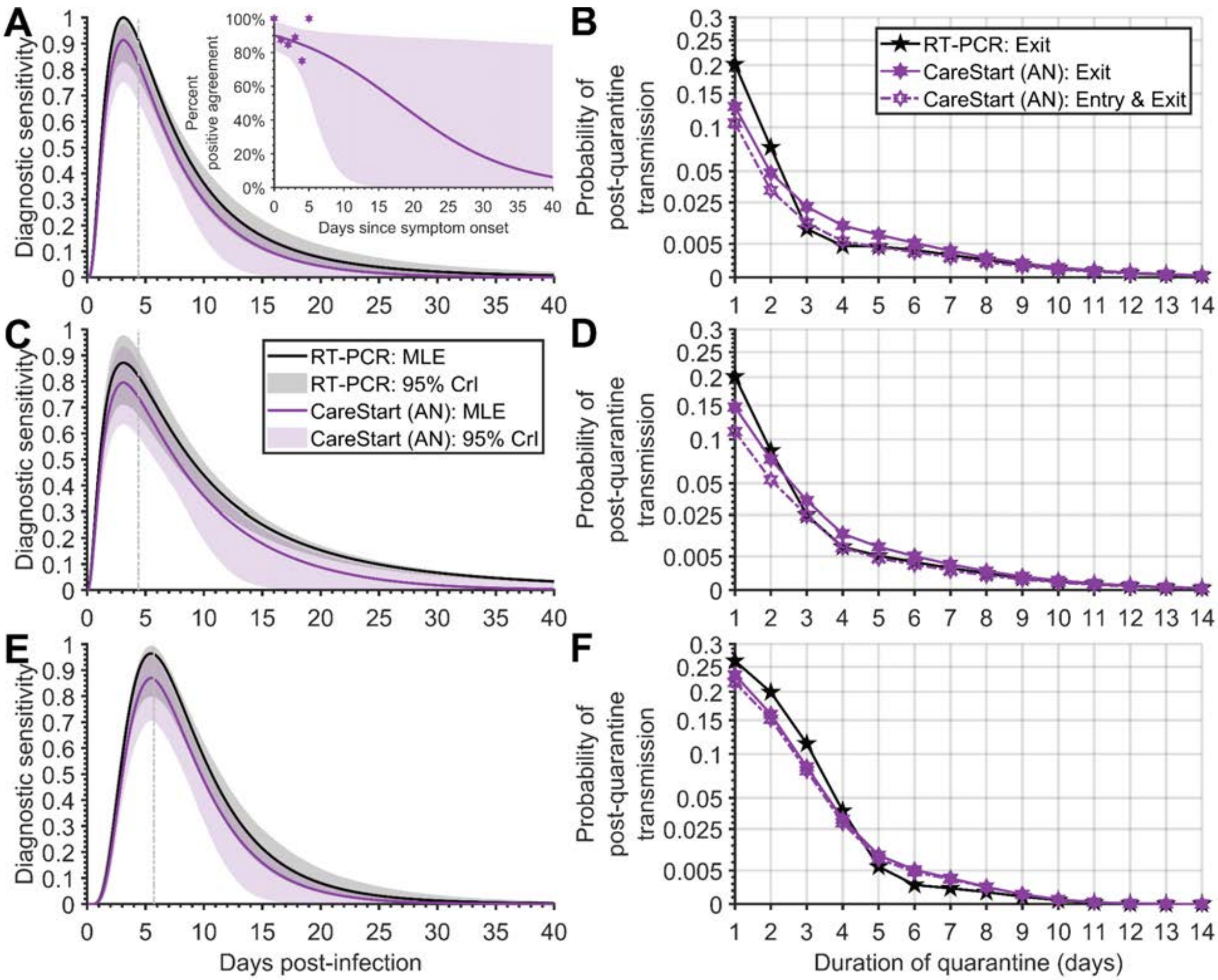


**Figure S15. The diagnostic sensitivity curve and probability of post-quarantine transmission for CareStart anterior nasal swab.** Specifying a 4.4-day incubation period, 35.1% of infections being asymptomatic, self-isolation upon symptom onset, (**A**) the RT-PCR diagnostic sensitivity curve (black) informed by data from Hellewell et al [9] using a log-Normal distribution and logistic regression model for the diagnostic sensitivity of the rapid antigen test (purple, solid line) informed by percent positive agreement data (purple hexagrams); (**B**) the probability of post-quarantine transmission for a RT-PCR test conducted 24 h before exit from quarantine (black stars), a rapid antigen test on exit (filled purple hexagrams), as well as a rapid antigen test conducted on both entry to and exit from quarantine (open purple hexagrams, dashed line); (**C**) the RT-PCR diagnostic sensitivity curve informed by data from Hellewell et al [9] using a log-Student's *t* distribution and logistic regression model for the diagnostic sensitivity of the rapid antigen test; (**D**) the probability of post-quarantine transmission for a RT-PCR test conducted 24 h before exit from quarantine, a rapid antigen test on exit, as well as a rapid antigen test conducted on both entry to and exit from quarantine. Specifying an 5.72-day incubation period, 35.1% of infections being asymptomatic, self-isolation upon symptom onset, (**E**) the RT-PCR diagnostic sensitivity curve informed by data from Hellewell et al [9] using a log-Normal distribution and logistic regression model for the diagnostic sensitivity of the rapid antigen test; and (**F**) the probability of post-quarantine transmission for a RT-PCR test conducted 24 h before exit from quarantine, a rapid antigen test on exit, as well as a rapid antigen test conducted on both entry to and exit from quarantine. The colored area denotes the 2.5 and 97.5 percentile constructed from 1,000 samples of the RT-PCR diagnostic sensitivity curve and the rapid antigen tests percent positive agreement curve.

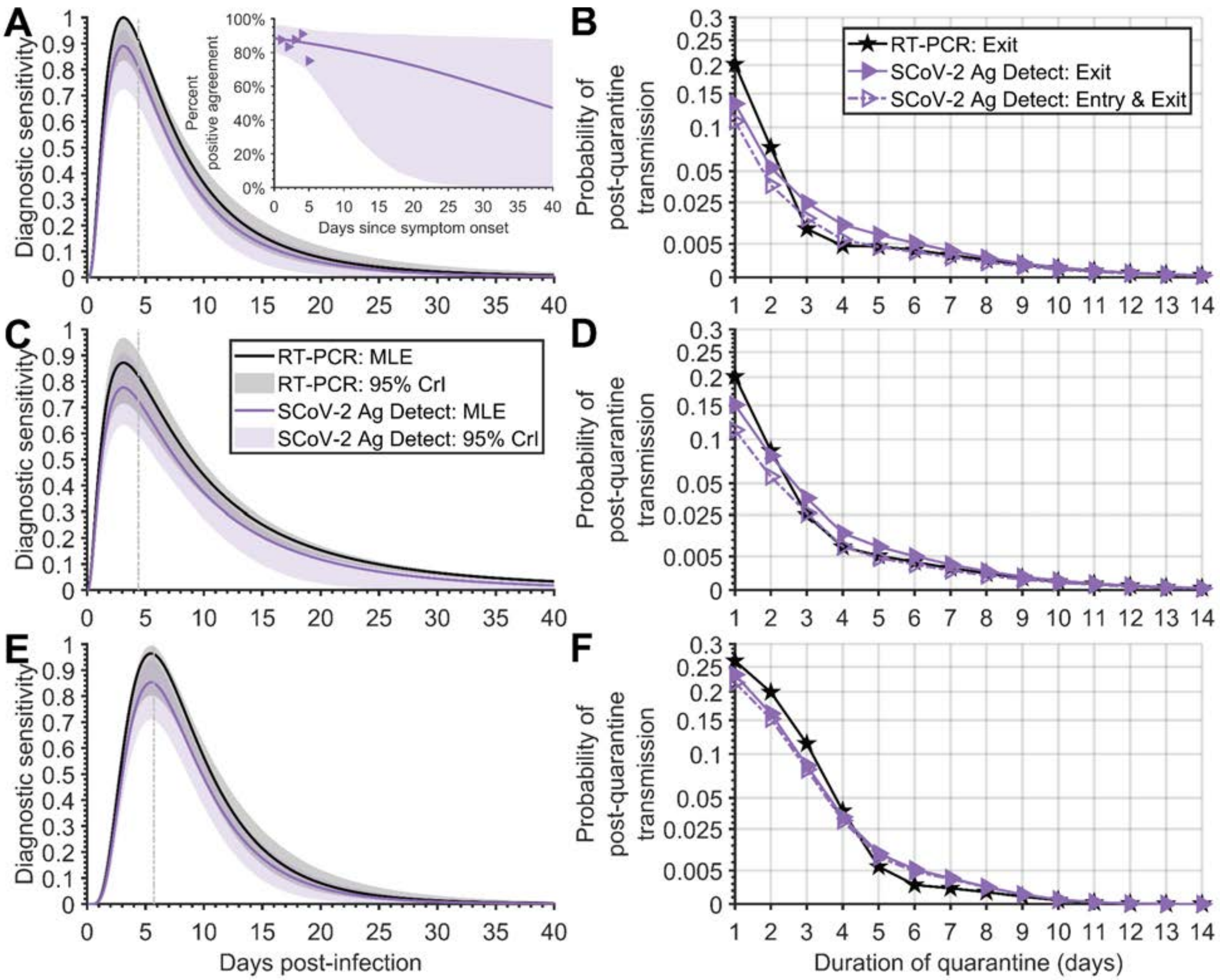


**Figure S16. The diagnostic sensitivity curve and probability of post-quarantine transmission for SCoV-2 Ag Detect.** Specifying a 4.4-day incubation period, 35.1% of infections being asymptomatic, self-isolation upon symptom onset, (**A**) the RT-PCR diagnostic sensitivity curve (black) informed by data from Hellewell et al [9] using a log-Normal distribution and logistic regression model for the diagnostic sensitivity of the rapid antigen test (purple, solid line) informed by percent positive agreement data (purple triangles); (**B**) the probability of post-quarantine transmission for a RT-PCR test conducted 24 h before exit from quarantine (black stars), a rapid antigen test on exit (filled purple triangles), as well as a rapid antigen test conducted on both entry to and exit from quarantine (open purple triangles, dashed line); (**C**) the RT-PCR diagnostic sensitivity curve informed by data from Hellewell et al [9] using a log-Student's *t* distribution and logistic regression model for the diagnostic sensitivity of the rapid antigen test; (**D**) the probability of post-quarantine transmission for a RT-PCR test conducted 24 h before exit from quarantine, a rapid antigen test on exit, as well as a rapid antigen test conducted on both entry to and exit from quarantine. Specifying an 5.72-day incubation period, 35.1% of infections being asymptomatic, self-isolation upon symptom onset, (**E**) the RT-PCR diagnostic sensitivity curve informed by data from Hellewell et al [9] using a log-Normal distribution and logistic regression model for the diagnostic sensitivity of the rapid antigen test; and (**F**) the probability of post-quarantine transmission for a RT-PCR test conducted 24 h before exit from quarantine, a rapid antigen test on exit, as well as a rapid antigen test conducted on both entry to and exit from quarantine. The colored area denotes the 2.5 and 97.5 percentile constructed from 1,000 samples of the RT-PCR diagnostic sensitivity curve and the rapid antigen tests percent positive agreement curve.

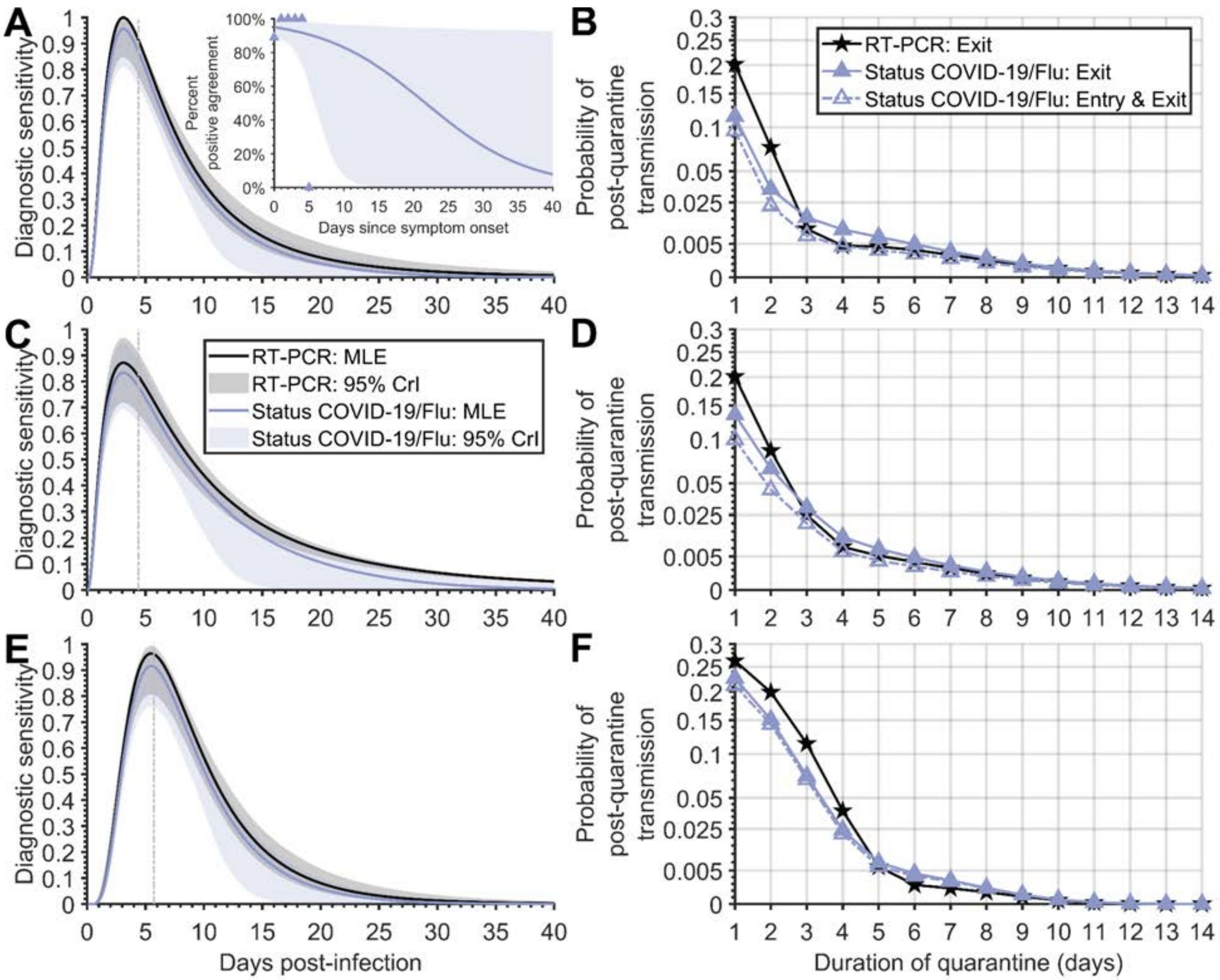


**Figure S17. The diagnostic sensitivity curve and probability of post-quarantine transmission for Status COVID-19/Flu.** Specifying a 4.4-day incubation period, 35.1% of infections being asymptomatic, self-isolation upon symptom onset, (**A**) the RT-PCR diagnostic sensitivity curve (black) informed by data from Hellewell et al [9] using a log-Normal distribution and logistic regression model for the diagnostic sensitivity of the rapid antigen test (purple, solid line) informed by percent positive agreement data (purple triangles); (**B**) the probability of post-quarantine transmission for a RT-PCR test conducted 24 h before exit from quarantine (black stars), a rapid antigen test on exit (filled purple triangles), as well as a rapid antigen test conducted on both entry to and exit from quarantine (open purple triangles, dashed line); (**C**) the RT-PCR diagnostic sensitivity curve informed by data from Hellewell et al [9] using a log-Student's *t* distribution and logistic regression model for the diagnostic sensitivity of the rapid antigen test; (**D**) the probability of post-quarantine transmission for a RT-PCR test conducted 24 h before exit from quarantine, a rapid antigen test on exit, as well as a rapid antigen test conducted on both entry to and exit from quarantine. Specifying an 5.72-day incubation period, 35.1% of infections being asymptomatic, self-isolation upon symptom onset, (**E**) the RT-PCR diagnostic sensitivity curve informed by data from Hellewell et al [9] using a log-Normal distribution and logistic regression model for the diagnostic sensitivity of the rapid antigen test; and (**F**) the probability of post-quarantine transmission for a RT-PCR test conducted 24 h before exit from quarantine, a rapid antigen test on exit, as well as a rapid antigen test conducted on both entry to and exit from quarantine. The colored area denotes the 2.5 and 97.5 percentile constructed from 1,000 samples of the RT-PCR diagnostic sensitivity curve and the rapid antigen tests percent positive agreement curve.

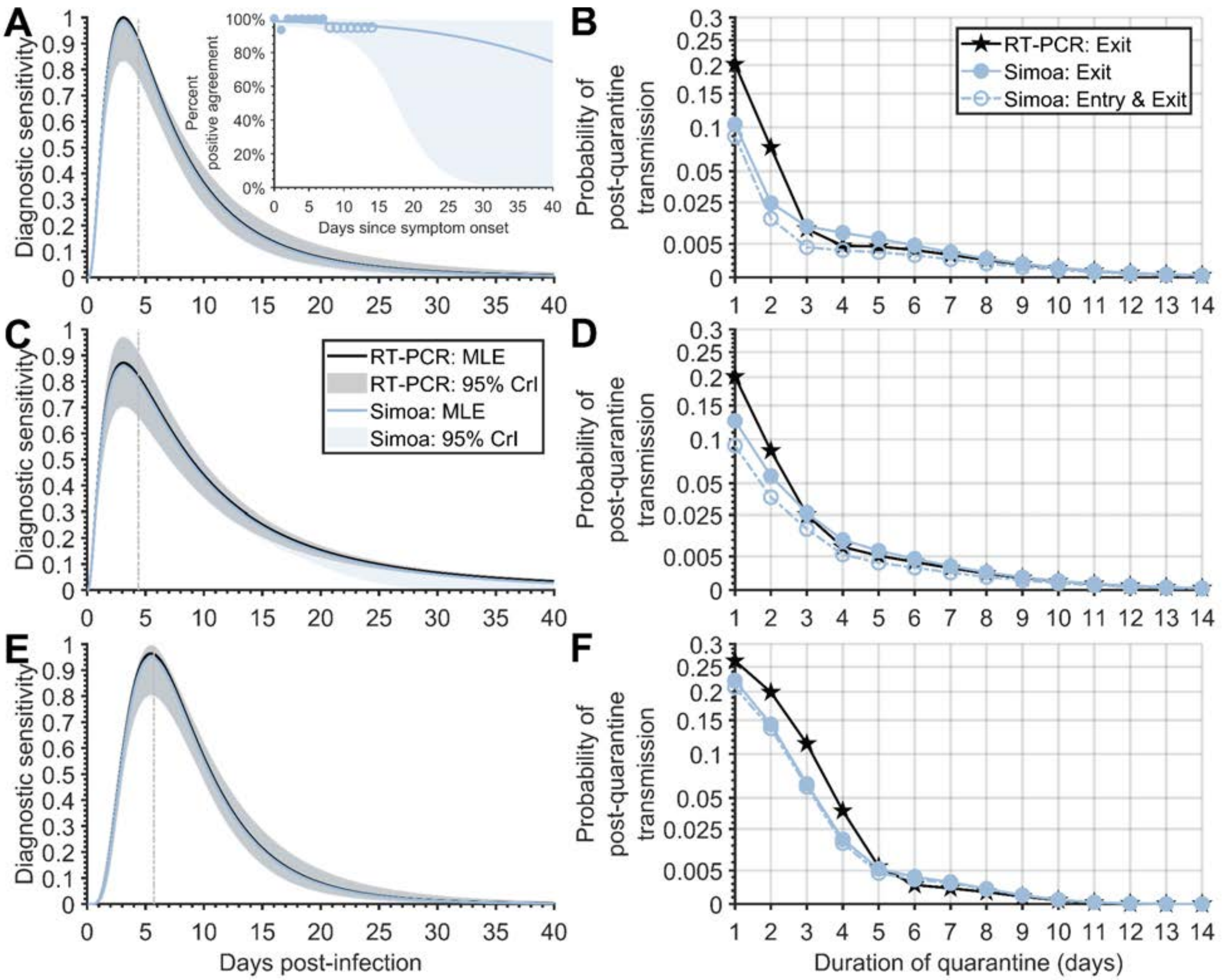


**Figure S18. The diagnostic sensitivity curve and probability of post-quarantine transmission for Simoa.** Specifying a 4.4-day incubation period, 35.1% of infections being asymptomatic, self-isolation upon symptom onset, (**A**) the RT-PCR diagnostic sensitivity curve (black) informed by data from Hellewell et al [9] using a log-Normal distribution and logistic regression model for the diagnostic sensitivity of the rapid antigen test (blue, solid line) informed by percent positive agreement data (blue circles); (**B**) the probability of post-quarantine transmission for a RT-PCR test conducted 24 h before exit from quarantine (black stars), a rapid antigen test on exit (filled blue circles), as well as a rapid antigen test conducted on both entry to and exit from quarantine (open blue circles, dashed line); (**C**) the RT-PCR diagnostic sensitivity curve informed by data from Hellewell et al [9] using a log-Student's *t* distribution and logistic regression model for the diagnostic sensitivity of the rapid antigen test; (**D**) the probability of post-quarantine transmission for a RT-PCR test conducted 24 h before exit from quarantine, a rapid antigen test on exit, as well as a rapid antigen test conducted on both entry to and exit from quarantine. Specifying an 5.72-day incubation period, 35.1% of infections being asymptomatic, self-isolation upon symptom onset, (**E**) the RT-PCR diagnostic sensitivity curve informed by data from Hellewell et al [9] using a log-Normal distribution and logistic regression model for the diagnostic sensitivity of the rapid antigen test; and (**F**) the probability of post-quarantine transmission for a RT-PCR test conducted 24 h before exit from quarantine, a rapid antigen test on exit, as well as a rapid antigen test conducted on both entry to and exit from quarantine. The colored area denotes the 2.5 and 97.5 percentile constructed from 1,000 samples of the RT-PCR diagnostic sensitivity curve and the rapid antigen tests percent positive agreement curve.

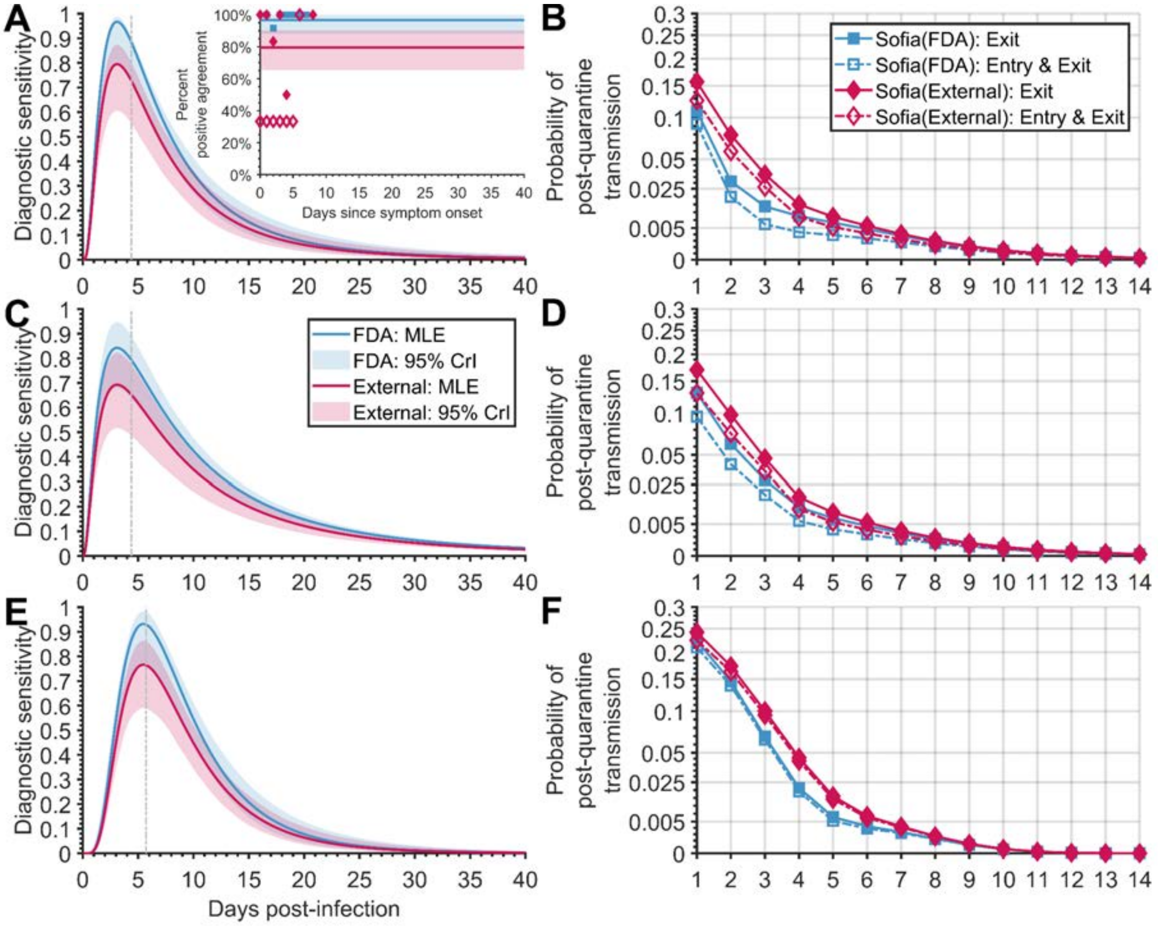


**Figure S19. The diagnostic sensitivity curve and probability of post-quarantine transmission for the internal and external percent positive agreement data for Sofia.** Specifying a 4.4-day incubation period, 35.1% of infections being asymptomatic, self-isolation upon symptom onset, and the RT-PCR diagnostic sensitivity curve informed by data from Hellewell et al [9] (**A**) using a log-Normal distribution to represent the RT-PCR diagnostic sensitivity curve and logistic regression model for the diagnostic sensitivity of the rapid antigen test (blue, solid line) informed by percent positive agreement data from the EUA FDA documentation (blue squares) and that informed by percent positive agreement data from community testing (red solid line; red diamonds); (**B**) the probability of post-quarantine transmission for a rapid antigen test on exit when informed by the FDA dataset (filled blue squares) or the external dataset (filled red diamonds), as well as a rapid antigen test conducted on both entry to and exit from quarantine when informed by the FDA dataset (open blue squares) or the external dataset (open red diamonds); (**C**) using a log-Student's *t* distribution to represent the RT-PCR diagnostic sensitivity curve and logistic regression model for the diagnostic sensitivity of the rapid antigen test informed by percent positive agreement data from the EUA FDA documentation and that informed by percent positive agreement data from community testing; (**C**) the probability of post-quarantine transmission for a rapid antigen test on exit when informed by the FDA dataset or the external dataset, as well as a rapid antigen test conducted on both entry to and exit from quarantine when informed by the FDA dataset or the external dataset. Specifying an 5.72-day incubation period, 35.1% of infections being asymptomatic, self-isolation upon symptom onset; (**E**) using a log-Normal distribution to represent the RT-PCR diagnostic sensitivity curve and logistic regression model for the diagnostic sensitivity of the rapid antigen test informed by percent positive agreement data from the EUA

FDA documentation and that informed by percent positive agreement data from community testing; and (**F**) the probability of post-quarantine transmission for a rapid antigen test on exit when informed by the FDA dataset or the external dataset, as well as a rapid antigen test conducted on both entry to and exit from quarantine when informed by the FDA dataset or the external dataset. The colored area denotes the 2.5 and 97.5 percentile constructed from 1,000 samples of the RT-PCR diagnostic sensitivity curve and the rapid antigen tests percent positive agreement curve.

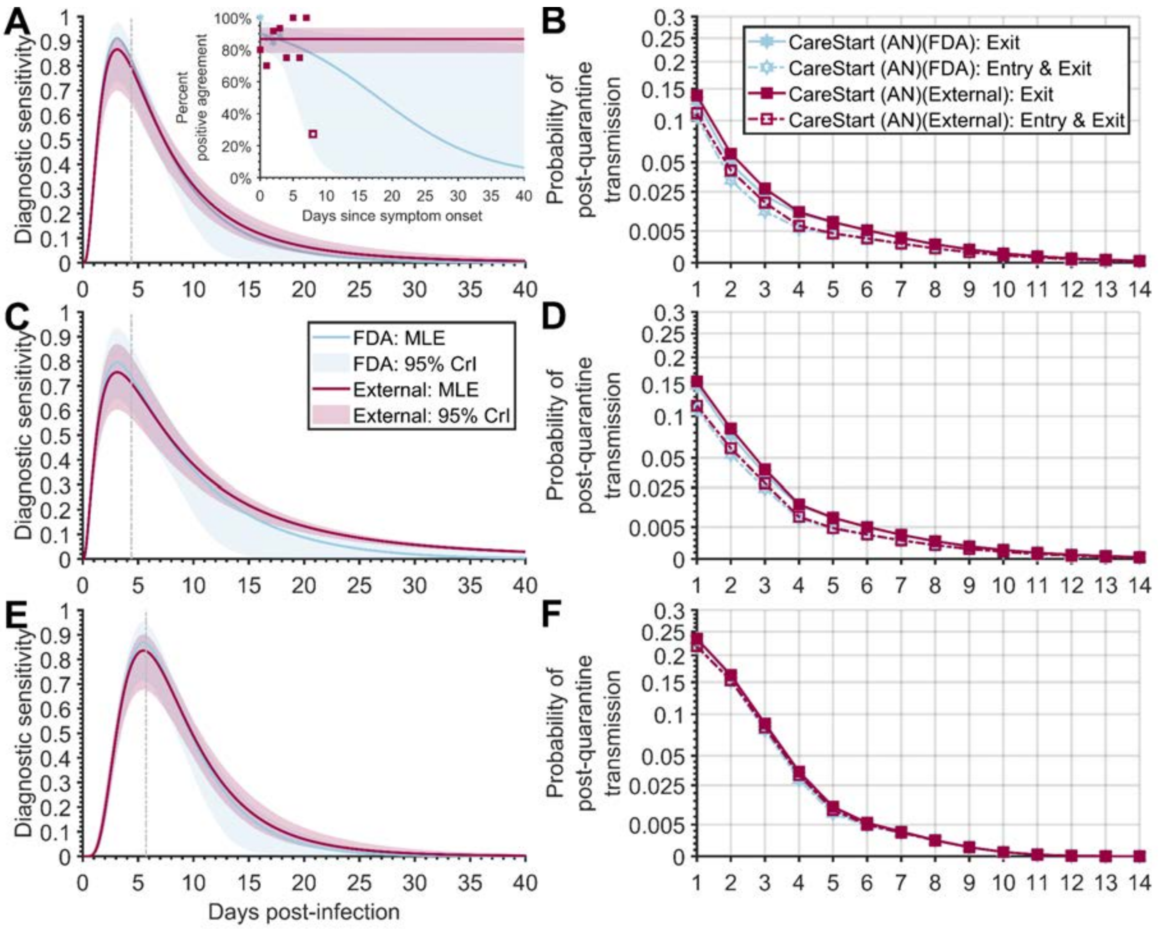


**Figure S20. The diagnostic sensitivity curve and probability of post-quarantine transmission for the industrial and external percent positive agreement data for CareStart.** Specifying a 4.4-day incubation period, 35.1% of infections being asymptomatic, self-isolation upon symptom onset, and the RT-PCR diagnostic sensitivity curve informed by data from Hellewell et al [9] (**A**) using a log-Normal distribution to represent the RT-PCR diagnostic sensitivity curve and logistic regression model for the diagnostic sensitivity of the rapid antigen test (blue, solid line) informed by percent positive agreement data from the EUA FDA documentation (blue hexagrams) and that informed by percent positive agreement data from community testing (red solid line; red squares); (**B**) the probability of post-quarantine transmission for a rapid antigen test on exit when informed by the FDA dataset (filled blue hexagrams) or the external dataset (filled red squares), as well as a rapid antigen test conducted on both entry to and exit from quarantine when informed by the FDA dataset (open blue hexagrams) or the external dataset (open red squares); (**C**) using a log-Student's *t* distribution to represent the RT-PCR diagnostic sensitivity curve and logistic regression model for the diagnostic sensitivity of the rapid antigen test informed by percent positive agreement data from the EUA FDA documentation and that informed by percent positive agreement data from community testing; (**C**) the probability of post-quarantine transmission for a rapid antigen test on exit when informed by the FDA dataset or the external dataset, as well as a rapid antigen test conducted on both entry to and exit from quarantine when informed by the FDA dataset or the external dataset. Specifying an 5.72-day incubation period, 35.1% of infections being asymptomatic, self-isolation upon symptom onset, (**E**) using a log-Normal distribution to represent the RT-PCR diagnostic sensitivity curve and logistic regression model for the diagnostic sensitivity of the rapid antigen test informed by percent positive agreement data from the EUA

FDA documentation and that informed by percent positive agreement data from community testing; and (**F**) the probability of post-quarantine transmission for a rapid antigen test on exit when informed by the FDA dataset or the external dataset, as well as a rapid antigen test conducted on both entry to and exit from quarantine when informed by the FDA dataset or the external dataset. The colored area denotes the 2.5 and 97.5 percentile constructed from 1,000 samples of the RT-PCR diagnostic sensitivity curve and the rapid antigen tests percent positive agreement curve.

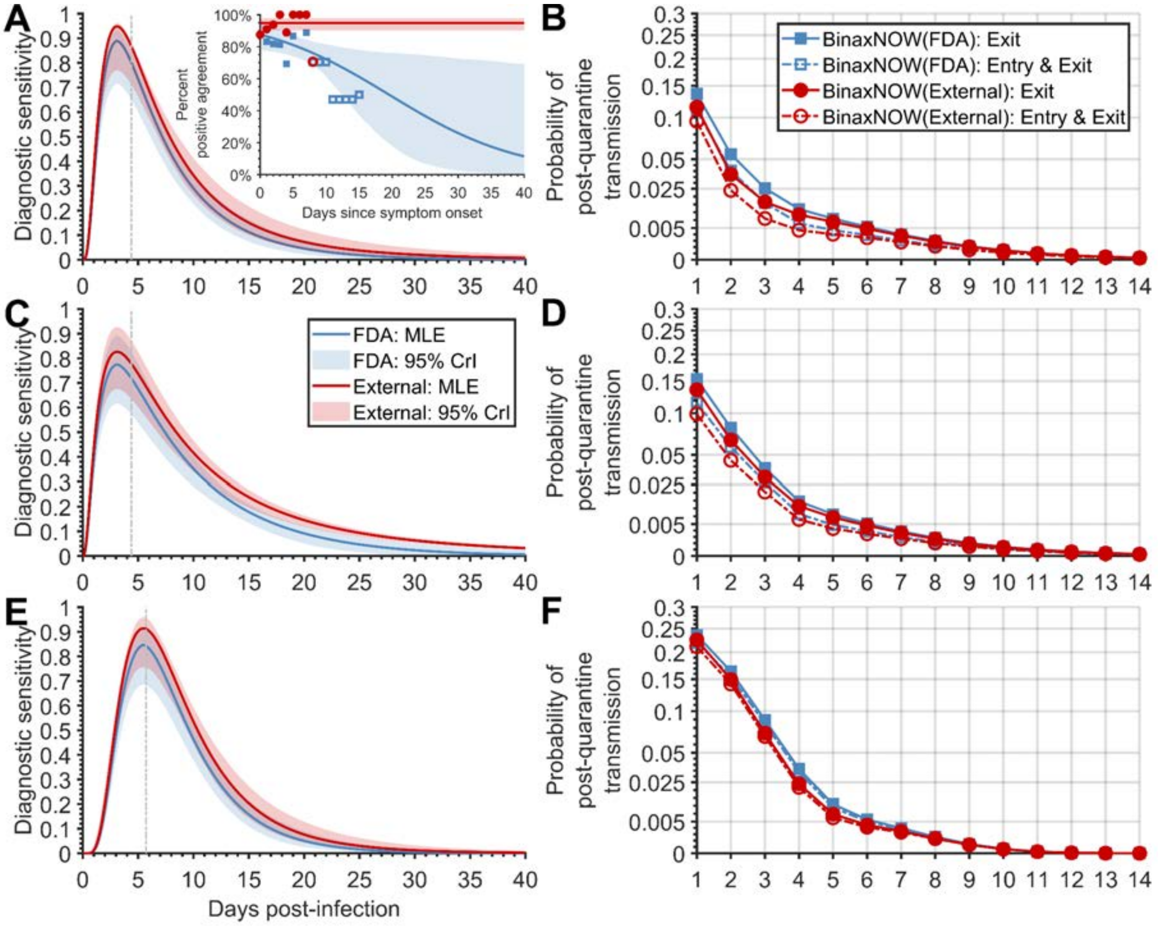


**Figure S21. The diagnostic sensitivity curve and probability of post-quarantine transmission for the industrial and external percent positive agreement data for BinaxNOW.** Specifying a 4.4-day incubation period, 35.1% of infections being asymptomatic, self-isolation upon symptom onset, and the RT-PCR diagnostic sensitivity curve informed by data from Hellewell et al [9] (**A**) using a log-Normal distribution to represent the RT-PCR diagnostic sensitivity curve and logistic regression model for the diagnostic sensitivity of the rapid antigen test (blue, solid line) informed by percent positive agreement data from the EUA FDA documentation (blue squares) and that informed by percent positive agreement data from community testing (red solid line; red circles); (**B**) the probability of post-quarantine transmission for a rapid antigen test on exit when informed by the FDA dataset (filled blue squares) or the external dataset (filled red circles), as well as a rapid antigen test conducted on both entry to and exit from quarantine when informed by the FDA dataset (open blue squares) or the external dataset (open red circles); (**C**) using a log-Student's *t* distribution to represent the RT-PCR diagnostic sensitivity curve and logistic regression model for the diagnostic sensitivity of the rapid antigen test informed by percent positive agreement data from the EUA FDA documentation and that informed by percent positive agreement data from community testing; (**D**) the probability of post-quarantine transmission for a rapid antigen test on exit when informed by the FDA dataset or the external dataset, as well as a rapid antigen test conducted on both entry to and exit from quarantine when informed by the FDA dataset or the external dataset. Specifying an 5.72-day incubation period, 35.1% of infections being asymptomatic, self-isolation upon symptom onset; (**E**) using a log-Normal distribution to represent the RT-PCR diagnostic sensitivity curve and logistic regression model for the diagnostic sensitivity of the rapid antigen test informed by percent positive agreement data from the EUA

FDA documentation and that informed by percent positive agreement data from community testing; and (**F**) the probability of post-quarantine transmission for a rapid antigen test on exit when informed by the FDA dataset or the external dataset, as well as a rapid antigen test conducted on both entry to and exit from quarantine when informed by the FDA dataset or the external dataset. The colored area denotes the 2.5 and 97.5 percentile constructed from 1,000 samples of the RT-PCR diagnostic sensitivity curve and the rapid antigen tests percent positive agreement curve.

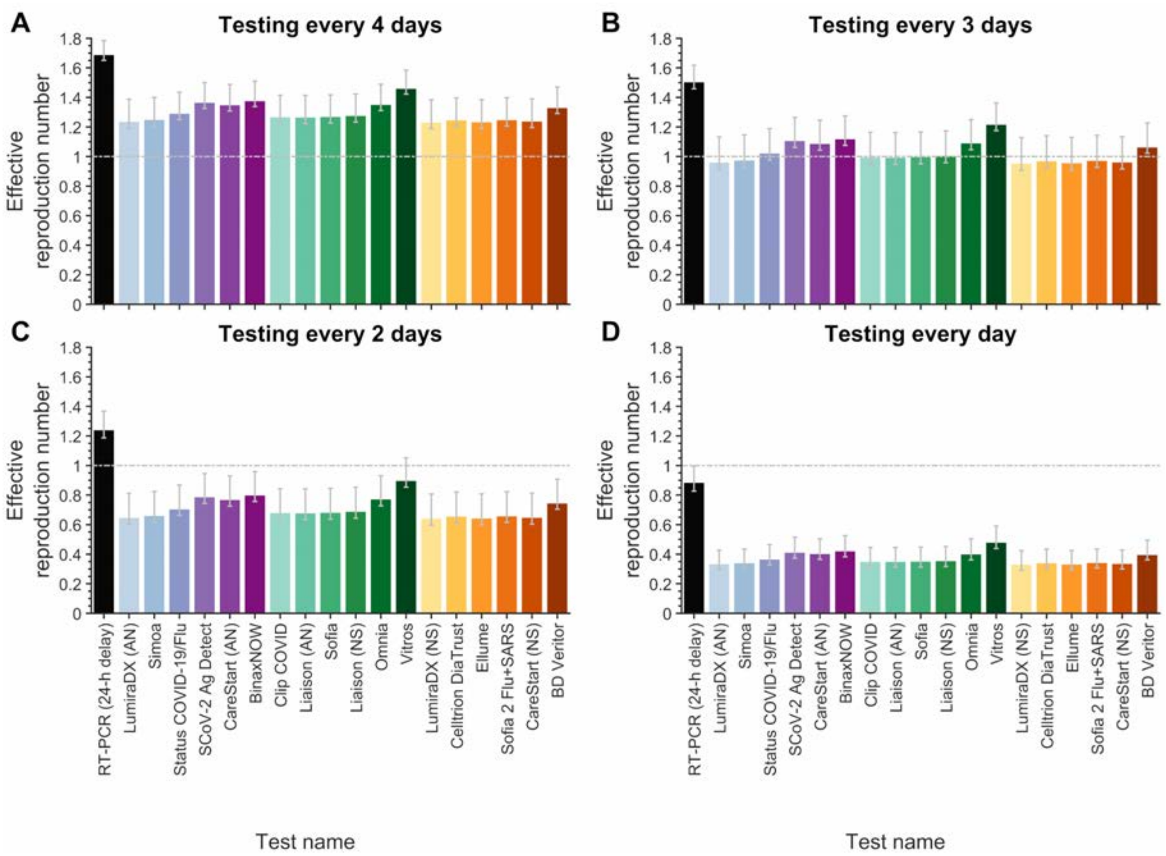


**Figure S22. The effective reproduction number when serial testing is conducted every day to one every four days with a rapid antigen test or RT-PCR.** Specifying a 4.4-day incubation period, 35.1% of infections as asymptomatic, a one-day delay in receiving RT-PCR, no delay rapid antigen test results, self-isolation upon symptom onset, and the RT-PCR diagnostic sensitivity curve (log-Normal) informed by data from Hellewell et al [9], the expected transmission with serial testing using an RT-PCR test (black) and the 18 rapid antigen tests (colors; *x* axis) when testing is conducted (**A**) every four days, (**B**) every three days, (**C**) every two days, and (**D**) every day.

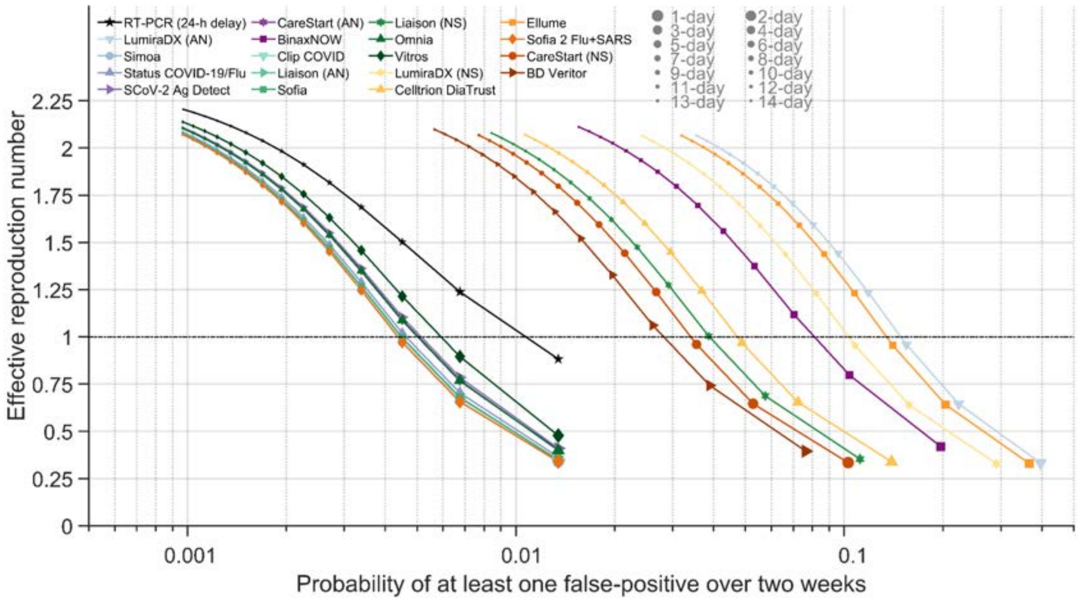


**Figure S23. The effective reproduction number, and probability of a false positive, for a range of frequencies of serial testing with RT-PCR and rapid antigen tests.** Specifying a 4.4-day incubation period, 35.1% of infections being asymptomatic, a 24-h delay in receiving RT-PCR test results, no-delay rapid antigen test results, self-isolation upon symptom onset, and the diagnostic sensitivity curve for the RT-PCR (log-Normal) was informed by data from Hellewell et al [9], the expected transmission with serial testing using an RT-PCR test with a one-day delay in obtaining test results (black stars) and the 18 rapid antigen tests (colors) for testing every day to every 14 days (small dots: longer time between tests; larger dots: shorter time between tests) and the corresponding probability of at least one false positive over a two-week period (*x* axis).

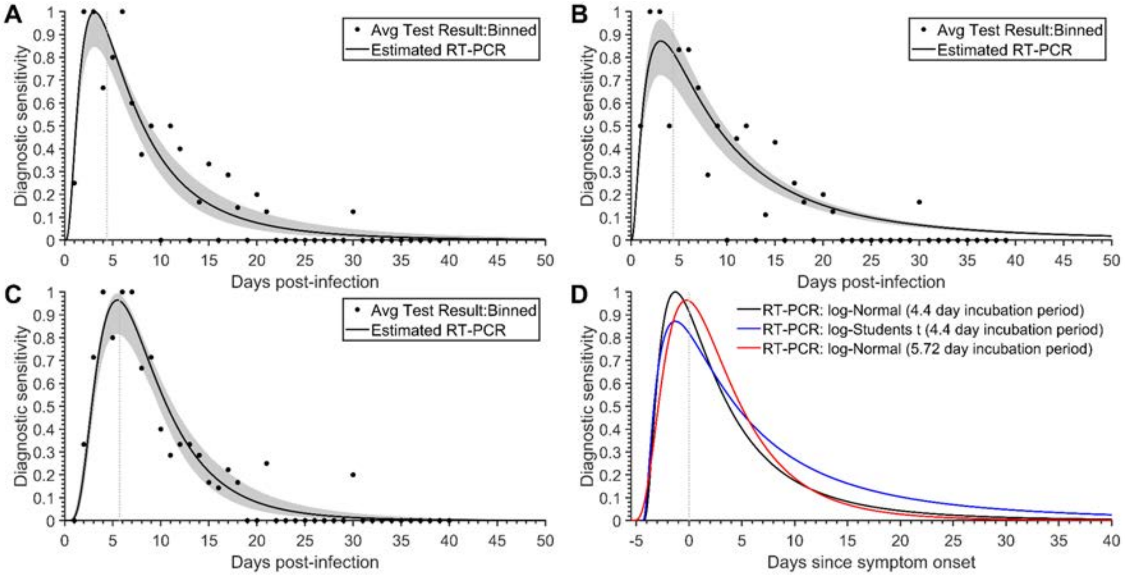


**Figure S24: Diagnostic sensitivity of RT-PCR.** The estimated diagnostic sensitivity of RT-PCR tests (black stars) informed by data from Hellewell et al [9] for (**A**) an incubation period of 4.4 days (dashed vertical line) and the temporal RT-PCR diagnostic sensitivity represented by a log-Normal distribution, (**B**) an incubation period of 4.4 days (dashed vertical line) and the temporal RT-PCR diagnostic sensitivity represented by a log-Student's *t* distribution, (**C**) an incubation period of 5.72 days (dashed vertical line) and the temporal RT-PCR diagnostic sensitivity represented by a log-Normal distribution and (**D**) a comparison of the three temporal RT-PCR diagnostic sensitivity curves relative to the time of symptom onset (dashed vertical line)

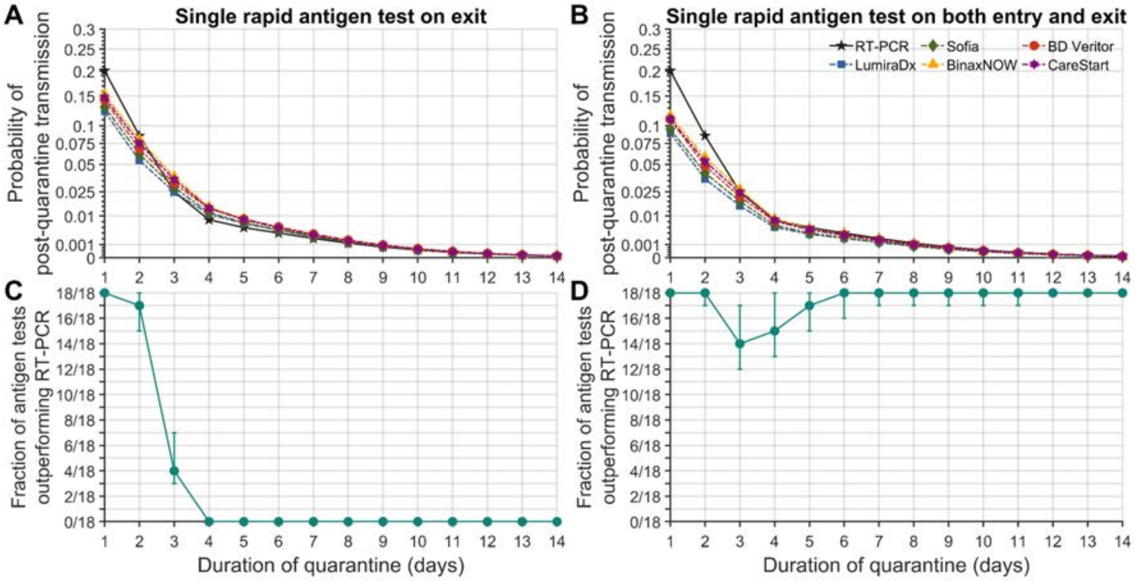


**Figure S25. Probability of post-quarantine transmission and fraction of antigen tests outperforming RT-PCR for quarantines of one to 14 days based on an alternative diagnostic sensitivity function.** Specifying a negative-binomial distribution for expected post-quarantine transmission, 35.1% of infections being asymptomatic, a 24-h delay in obtaining RT-PCR test results, no delay in receiving rapid antigen test results, an incubation period of 4.4 days, self-isolation upon symptom onset, and the diagnostic sensitivity curve represented by a log-Student's *t* distribution, the probability of post-quarantine transmission when conducting an RT-PCR test only on exit (solid line; black stars) and the rapid antigen tests (dashed lines) LumiraDx (blue squares); Sofia (green diamonds); BinaxNOW (yellow triangles); BD Veritor (red circles); and CareStart (purple hexagrams) performed on (**A**) exit and (**B**) both entry and exit; and the fraction of the 18 rapid antigen tests whose use conferred a lower probability of post-quarantine transmission than did an RT-PCR test conducted 24 h before exit from quarantine, when the rapid antigen test was conducted (**C**) on exit and **(D**) on both entry and exit.

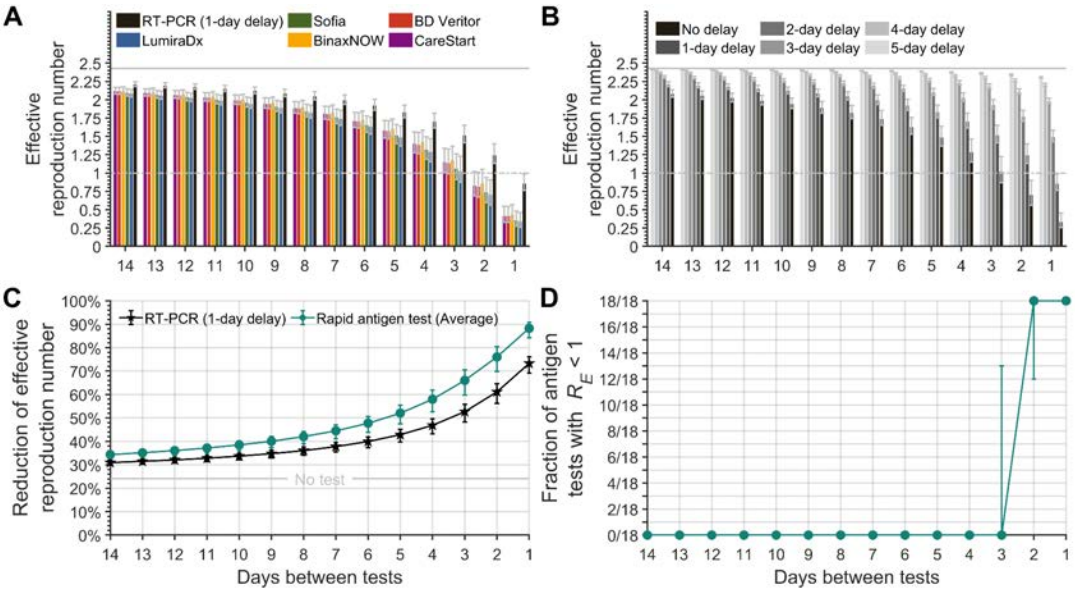


**Figure S26. Effective reproduction number with frequencies of serial testing from every day to every 14 days and isolation of positives, based on an alternative diagnostic sensitivity function.** Specifying 35.1% of infections as asymptomatic, a one-day delay in receiving RT-PCR and rapid antigen test results, an incubation period of 4.4 days, self-isolation upon symptom onset, and a RT-PCR diagnostic sensitivity curve represented by a log-Student's *t* distribution (**A**) the expected effective reproduction number with serial testing using an RT-PCR test (black) and the rapid antigen tests LumiraDx (blue); Sofia (green); BinaxNOW (yellow); BD Veritor (red); and CareStart (purple), and (**B**) for serial testing every day to every 14 days with a zero- to five-day delay (black to light gray) in obtaining the results for an RT-PCR test and isolation of positives in comparison to no testing (solid gray line). (**C**) The fraction of rapid antigen tests of the 18 tests that had a lower effective reproduction number than a RT-PCR test with a 24-h delay, and (**D**) that had an effective reproduction number ($R_E$) below one for testing frequencies ranging from every day to every two weeks and isolating positives.

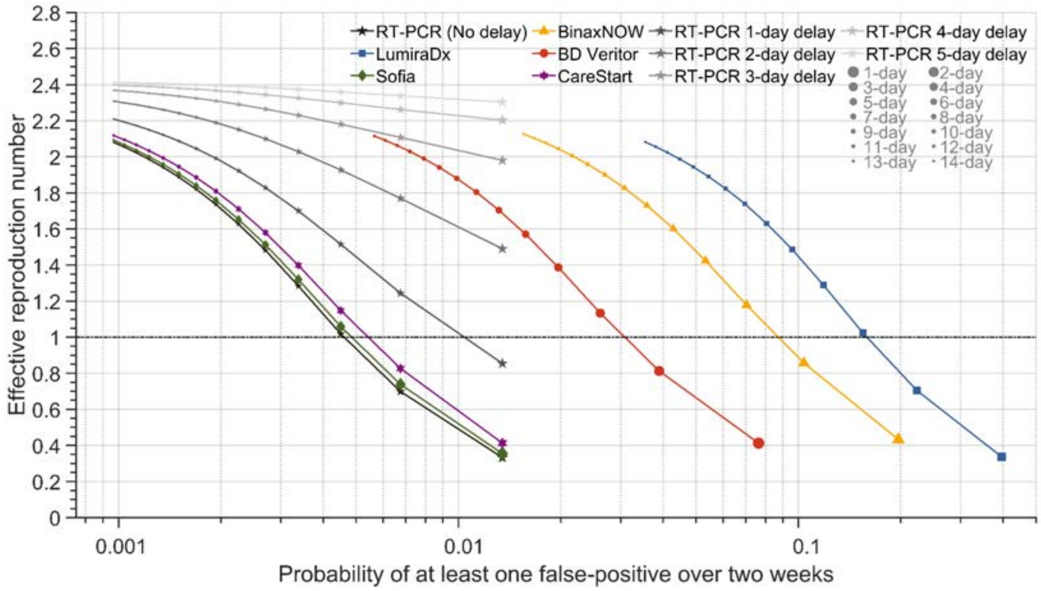


**Figure S27. Effective reproduction numbers and probabilities of false positives for varying frequencies of serial testing with RT-PCR and rapid antigen tests and isolation of positives, based on an alternative diagnostic sensitivity function.** Specifying 35.1% of infections being asymptomatic, no delay in receiving RT-PCR and rapid antigen test results, an incubation period of 4.4 days, self-isolation upon symptom onset, and the diagnostic sensitivity curve represented by a log-Student's *t* distribution, the expected transmission with serial RT-PCR testing with a zero- to five-day delay (black star gradient) in obtaining test results, and the rapid antigen test LumiraDx (blue square); Sofia (green diamond); BinaxNOW (yellow triangle); BD Veritor (red circle); and CareStart (purple hexagram) for testing every day to every 14 days (small dots: longer time between tests; larger dots: shorter time between tests) and the corresponding probability of at least one false positive over a two-week period (*x* axis).

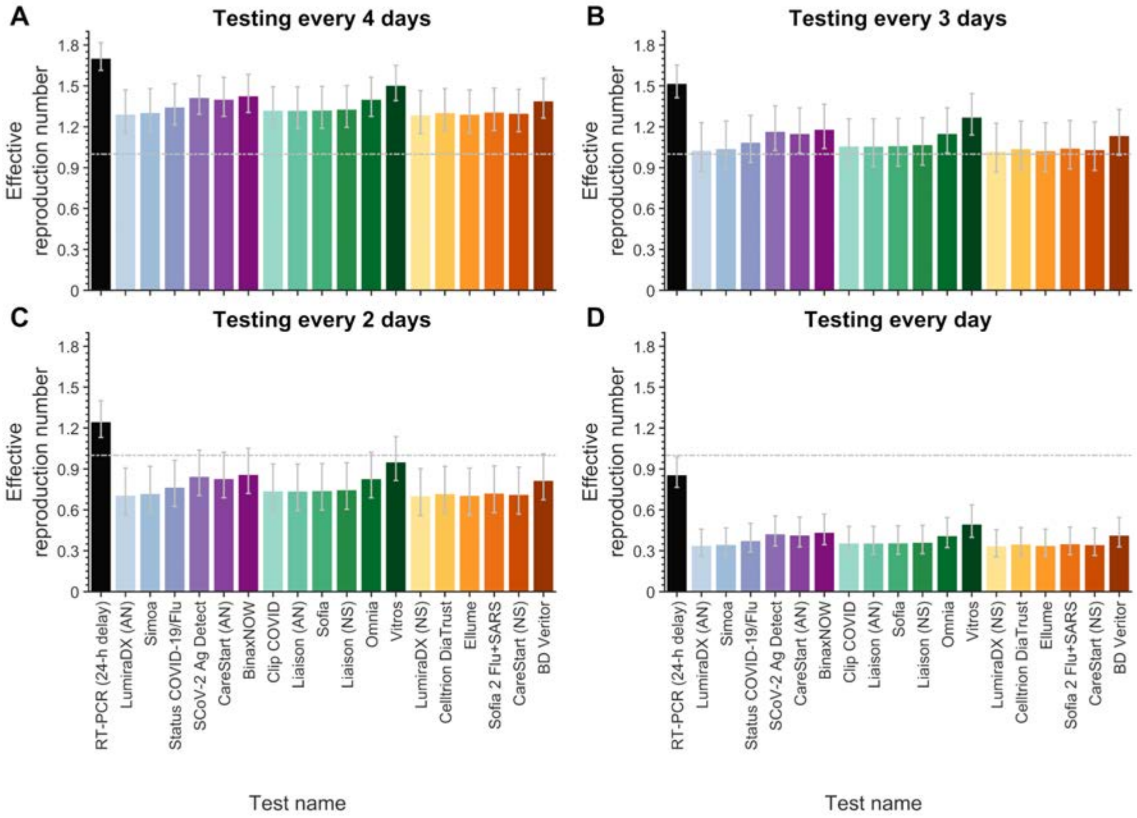


**Figure S28. The effective reproduction number when serial testing is conducted every day to every four days with a rapid antigen test or RT-PCR for an alternative diagnostic sensitivity function.** Specifying a 4.4-day incubation period, 35.1% of infections as asymptomatic, a one-day delay in receiving RT-PCR and rapid antigen test results, self-isolation upon symptom onset, and the RT-PCR diagnostic sensitivity curve represented by a log-Student's *t* distribution, the expected transmission with serial testing using an RT-PCR test (black) and the 18 rapid antigen tests (colors; *x* axis) when testing is conducted (**A**) every four days, (**B**) every three days, (**C**) every two days and (**F**) every day.

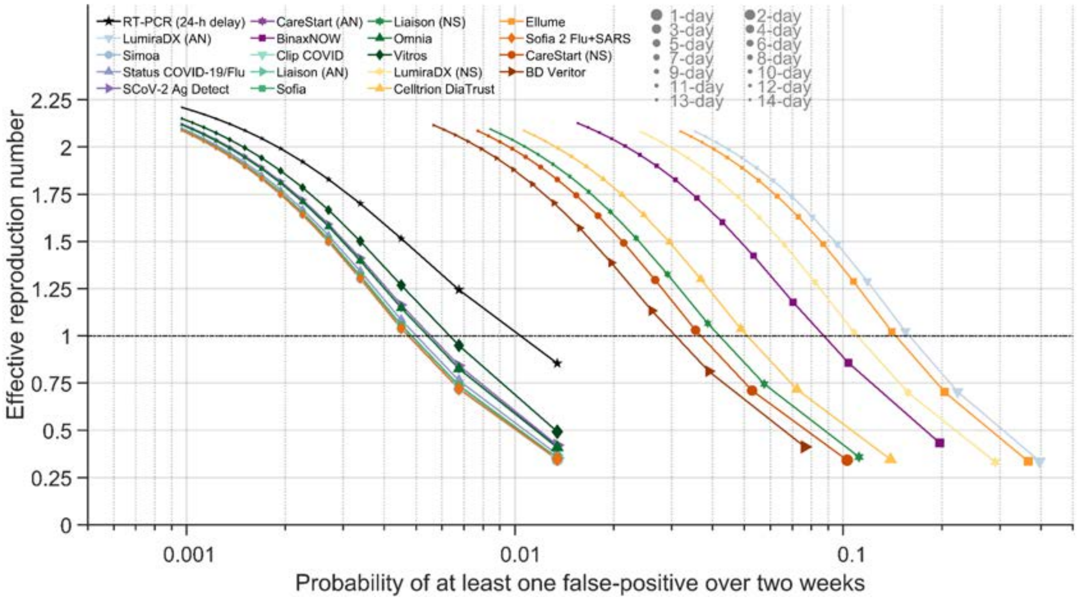


**Figure S29. The effective reproduction number, and probability of a false positive, for a range of frequencies of serial testing with RT-PCR and rapid antigen tests for an alternative diagnostic sensitivity function.** Specifying a 4.4-day incubation period, 35.1% of infections being asymptomatic, a 24-h delay in receiving RT-PCR test results, and no-delay rapid antigen test results, self-isolation upon symptom onset, and the diagnostic sensitivity curve for the RT-PCR represented by a log-Student's *t* distribution, the expected transmission with serial testing using an RT-PCR test with a one-day delay in obtaining test results (black stars) and the 18 rapid antigen tests (colors) for testing every day to every 14 days (small symbols: longer time between tests; larger symbols: shorter time between tests) and the corresponding probability of at least one false positive over a two-week period (*x* axis).

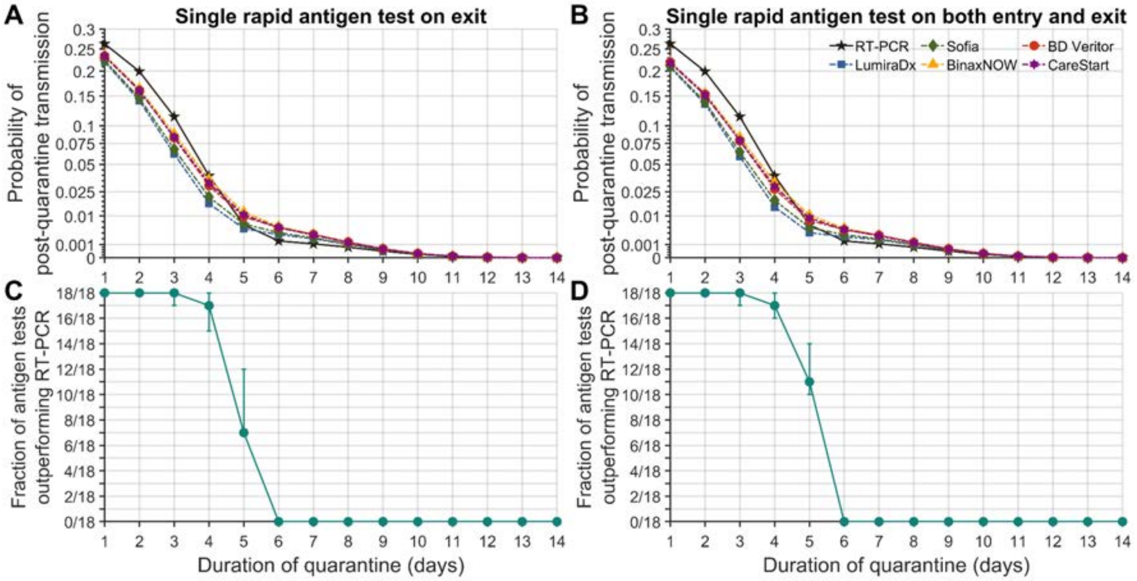


**Figure S30. Probability of post-quarantine transmission and fraction of antigen tests outperforming RT-PCR for quarantines of one to 14 days based on an alternative incubation period.** Specifying a negative-binomial distribution for expected post-quarantine transmission, 35.1% of infections being asymptomatic, a 24-h delay in obtaining RT-PCR test results, no delay in receiving rapid antigen test results, an incubation period of 5.72 days, self-isolation upon symptom onset, and the diagnostic sensitivity curve represented by a log-Normal distribution, the probability of post-quarantine transmission when conducting an RT-PCR test only on exit (solid line; black stars) and the rapid antigen tests (dashed lines) LumiraDx (blue squares); Sofia (green diamonds); BinaxNOW (yellow triangles); BD Veritor (red circles); and CareStart (purple hexagrams) performed on (**A**) exit and (**B**) both entry and exit; and the fraction of the 18 rapid antigen tests whose use conferred a lower probability of post-quarantine transmission than did an RT-PCR test conducted 24 h before exit from quarantine, when the rapid antigen test was conducted (**C**) on exit and **(D**) on both entry and exit.

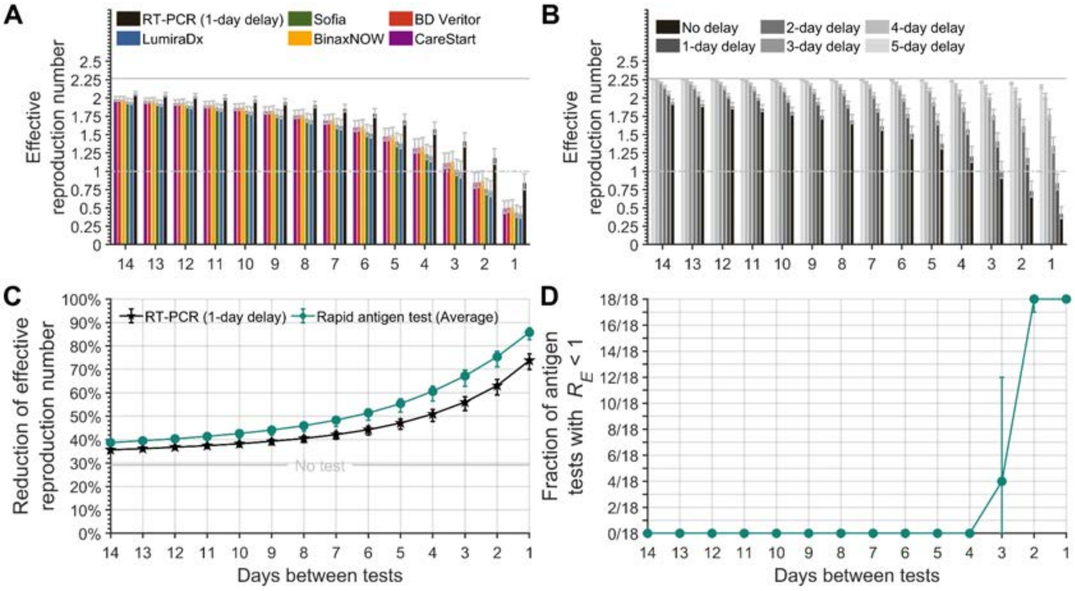


**Figure S31. Effective reproduction number with frequencies of serial testing from every day to every 14 days and isolation of positives, based on an alternative incubation period.** Specifying 35.1% of infections as asymptomatic, a one-day delay in receiving RT-PCR and rapid antigen test results, an incubation period of 5.72 days, self-isolation upon symptom onset, and a RT-PCR diagnostic sensitivity curve represented by a log-Normal distribution (**A**) the expected effective reproduction number with serial testing using an RT-PCR test (black) and the rapid antigen tests LumiraDx (blue); Sofia (green); BinaxNOW (yellow); BD Veritor (red); and CareStart (purple), and (**B**) for serial testing every day to every 14 days with a zero- to five-day delay (black to light gray) in obtaining the results for an RT-PCR test and isolation of positives in comparison to no testing (solid gray line). (**C**) The fraction of rapid antigen tests of the 18 tests that had a lower effective reproduction number than a RT-PCR test with a 24-h delay, and (**D**) that had an effective reproduction number ($R_E$) below one for testing frequencies ranging from every day to every two weeks and isolating positives.

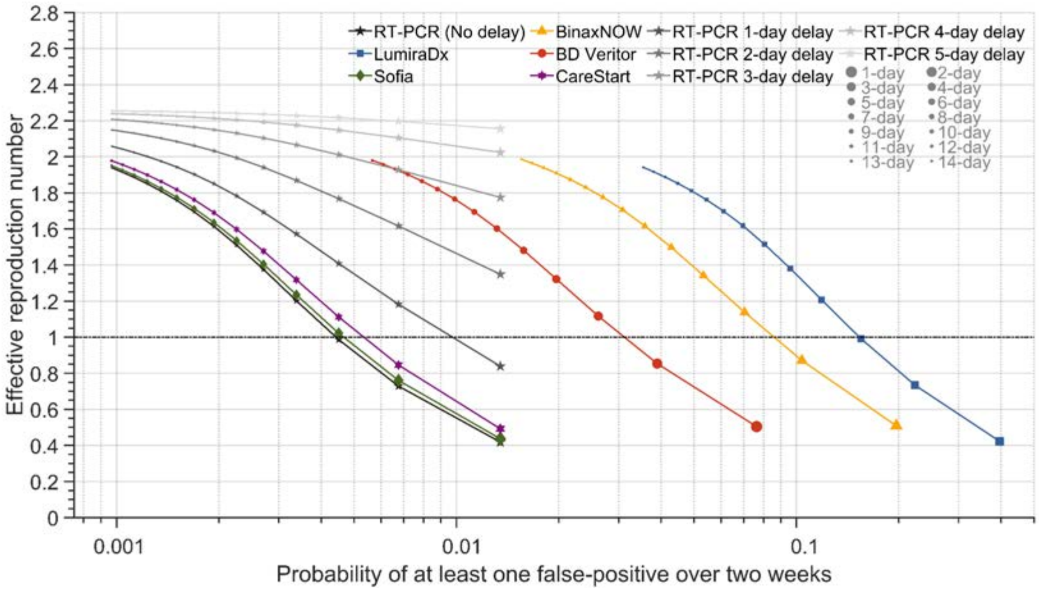


**Figure S32. Effective reproduction numbers and probabilities of false positives for varying frequencies of serial testing with RT-PCR and rapid antigen tests and isolation of positives, based on an alternative incubation period.** Specifying 35.1% of infections being asymptomatic, no delay in receiving RT-PCR and rapid antigen test results, an incubation period of 5.72 days, self-isolation upon symptom onset, and the diagnostic sensitivity curve represented by a log-Normal distribution, the expected transmission with serial RT-PCR testing with a zero- to five-day delay (black star gradient) in obtaining test results, and the rapid antigen test LumiraDx (blue square); Sofia (green diamond); BinaxNOW (yellow triangle); BD Veritor (red circle); and CareStart (purple hexagram) for testing every day to every 14 days (small dots: longer time between tests; larger dots: shorter time between tests) and the corresponding probability of at least one false positive over a two-week period (*x* axis).

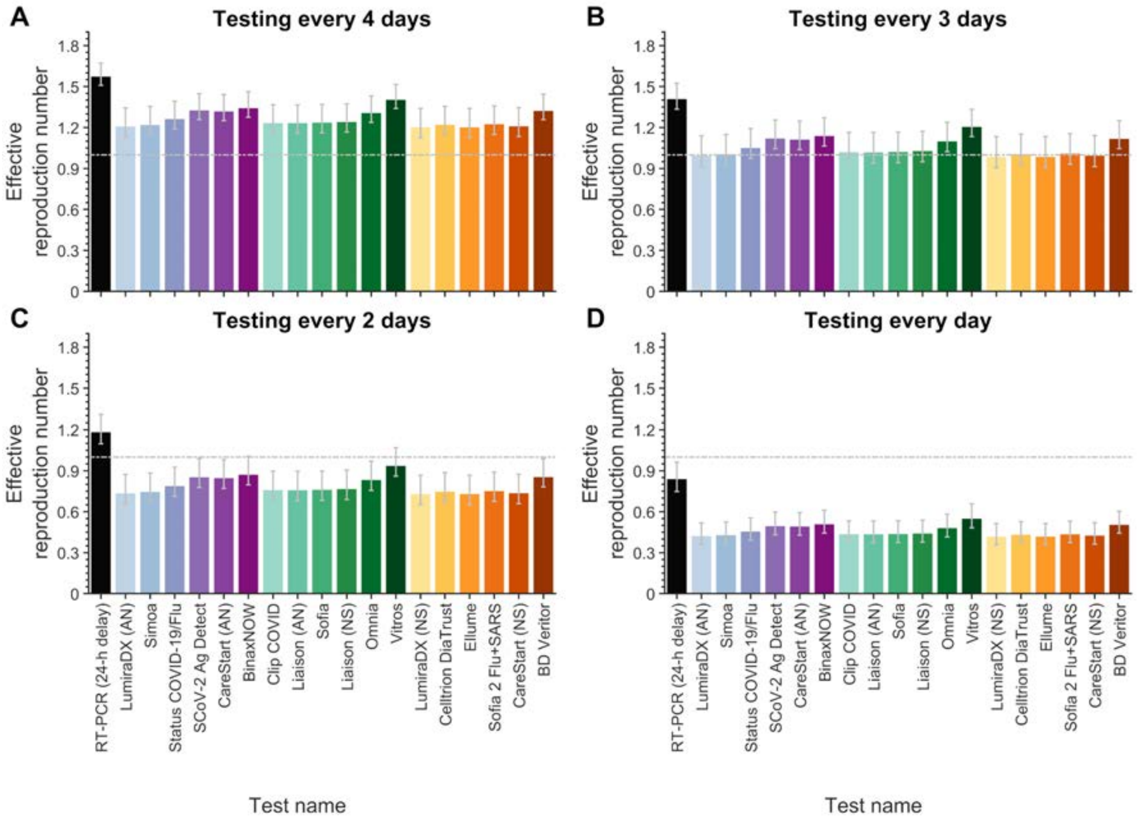


**Figure S33. The effective reproduction number when serial testing is conducted every day to every four days with a rapid antigen test or RT-PCR for an alternative incubation period.** Specifying a 5.72-day incubation period, 35.1% of infections as asymptomatic, a one-day delay in receiving RT-PCR and rapid antigen test results, self-isolation upon symptom onset, and the RT-PCR diagnostic sensitivity curve represented by a log-Normal distribution, the expected transmission with serial testing using an RT-PCR test (black) and the 18 rapid antigen tests (colors; *x* axis) when testing is conducted (**A**) every four days, (**B**) every three days, (**C**) every two days and (**F**) every day.

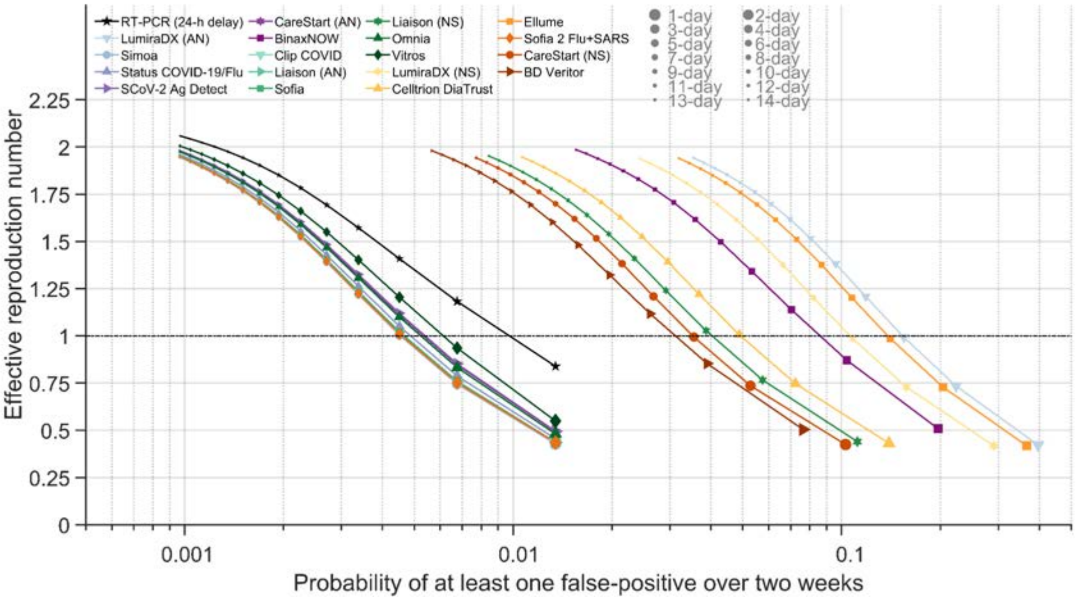


**Figure S34. The effective reproduction number, and probability of a false positive, for a range of frequencies of serial testing with RT-PCR and rapid antigen tests for an alternative incubation period.** Specifying a 5.72-day incubation period, 35.1% of infections being asymptomatic, a 24-h delay in receiving RT-PCR test results, and no-delay rapid antigen test results, self-isolation upon symptom onset, and the diagnostic sensitivity curve for the RT-PCR represented by a log-Normal distribution, the expected transmission with serial testing using an RT-PCR test with a one-day delay in obtaining test results (black stars) and the 18 rapid antigen tests (colors) for testing every day to every 14 days (small symbols: longer time between tests; larger symbols: shorter time between tests) and the corresponding probability of at least one false positive over a two-week period (*x* axis).

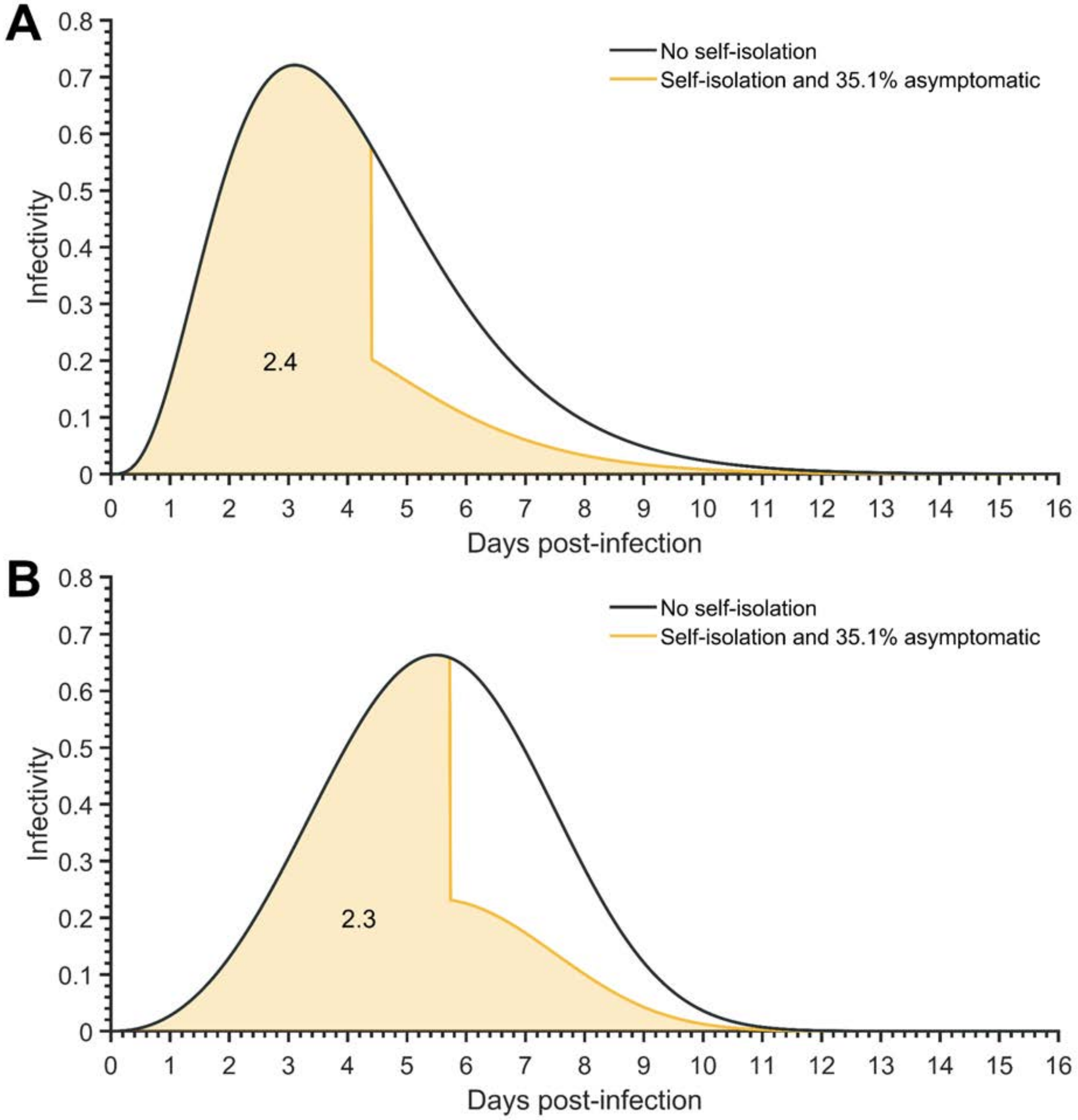


**Figure S35: The average temporal infectivity curve.** Specifying a basic reproduction number of 3.2 and 35.1% of infections being asymptomatic, the average infectivity curve for a known time of infection under no self-isolation upon symptom onset (black) and perfect isolation upon symptom onset (yellow line) for (**A**) an incubation period of 4.4 days (resulting in 2.4 secondary infections, yellow fill), and (**B**) an incubation period of 5.72 days (resulting in 2.3 secondary infections, yellow fill).

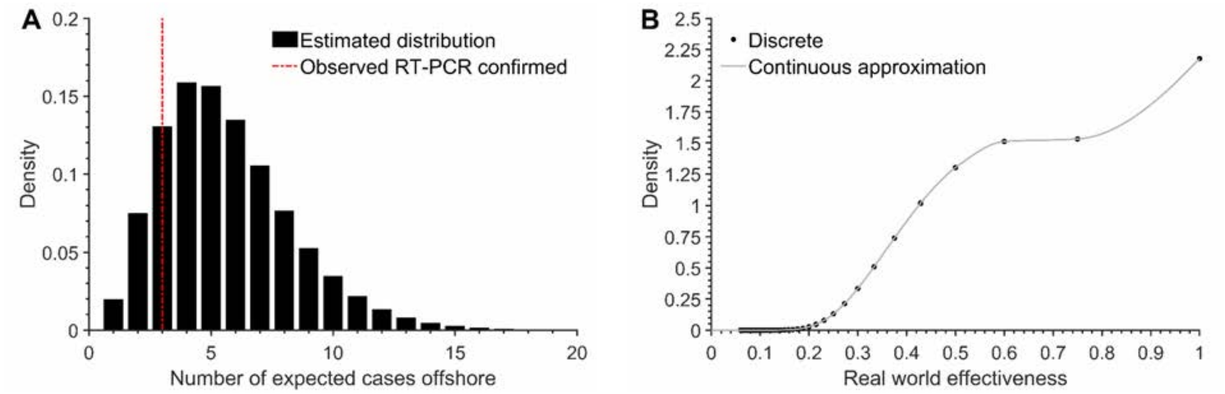


**Figure S36. Estimated infectious cases offshore.** Specifying a basic reproduction number of 3.2 and 100% infections being asymptomatic, an incubation period of 5.72 days, the distribution of (**A**) the estimated number of infectious individuals in the offshore environment (black bars) relative to the number of RT-PCR confirmed cases observed (red dashed line) and (**B**) the estimated distribution of the effectiveness of the serial testing strategy conducted offshore based on the three cases detected in the offshore environment (black circles) and the approximated continuous distribution of the effectiveness of the serial testing (gray line). The continuous distribution was approximated using an interpolation function.

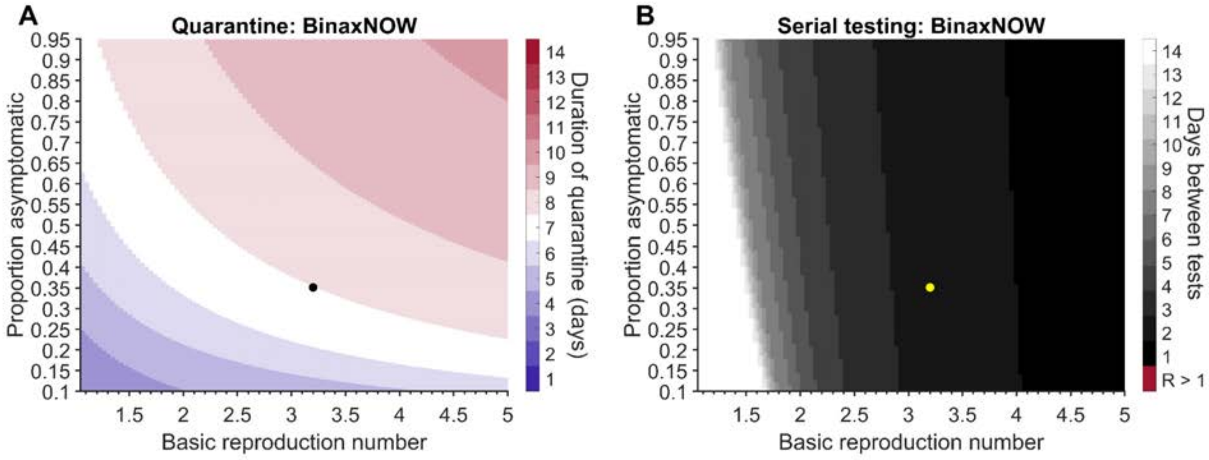


**Figure S37. The impact of asymptomatic infection and basic reproduction number on quarantine duration and frequency of serial testing for BinaxNOW.** Specifying an incubation period of 4.4 days and RT-PCR diagnostic sensitivity represented by a log-Normal distribution (**A**) the quarantine duration for a RA test on exit (blue and red gradient) that has an equivalent or lower probability of post-quarantine transmission than the baseline scenario (35.1% asymptomatic infection and a basic reproduction number of 3.2) for a 7-day quarantine (black dot) and (**B**) the frequency of serial testing required to maintain the effective reproduction number below one (black and white gradient; baseline indicated by yellow dot) for asymptomatic infections ranging from 10% to 95% and a basic reproduction number ranging from 1.05 to 5.

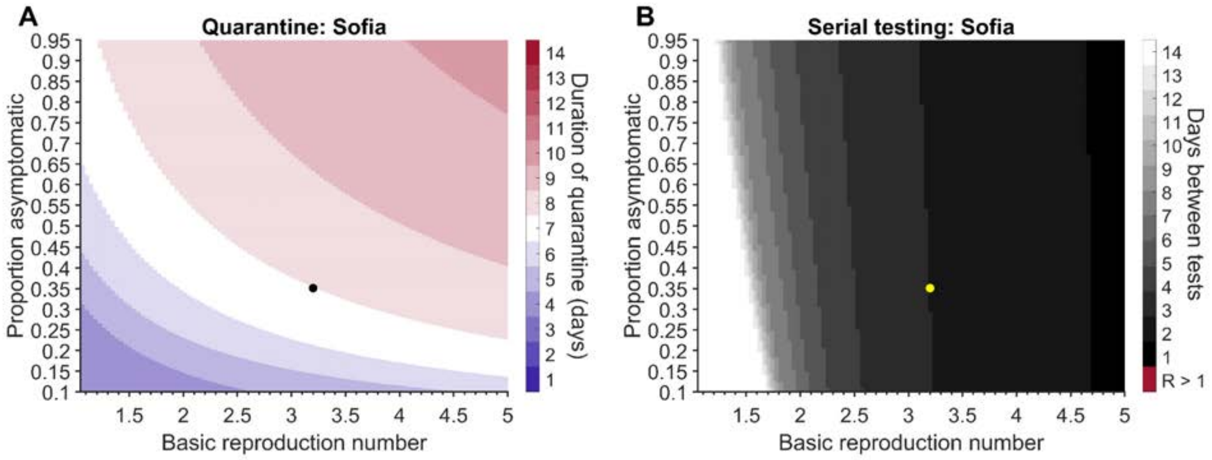


**Figure S38. The impact of asymptomatic infection and basic reproduction number on quarantine duration and frequency of serial testing for Sofia.** Specifying an incubation period of 4.4 days and RT-PCR diagnostic sensitivity represented by a log-Normal distribution (**A**) the quarantine duration for a RA test on exit (blue and red gradient) that has an equivalent or lower probability of post-quarantine transmission than the baseline scenario (35.1% asymptomatic infection and a basic reproduction number of 3.2) for a 7-day quarantine (black dot) and (**B**) the frequency of serial testing required to maintain the effective reproduction number below one (black and white gradient; baseline indicated by yellow dot) for asymptomatic infections ranging from 10% to 95% and a basic reproduction number ranging from 1.05 to 5.

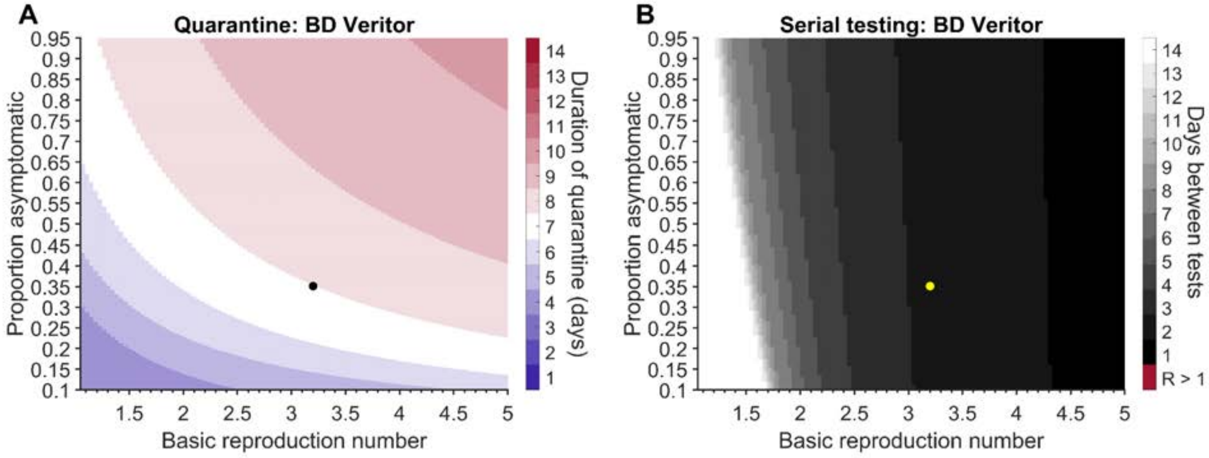


**Figure S39. The impact of asymptomatic infection and basic reproduction number on quarantine duration and frequency of serial testing for BD Veritor.** Specifying an incubation period of 4.4 days and RT-PCR diagnostic sensitivity represented by a log-Normal distribution (**A**) the quarantine duration for a RA test on exit (blue and red gradient) that has an equivalent or lower probability of post-quarantine transmission than the baseline scenario (35.1% asymptomatic infection and a basic reproduction number of 3.2) for a 7-day quarantine (black dot) and (**B**) the frequency of serial testing required to maintain the effective reproduction number below one (black and white gradient; baseline indicated by yellow dot) for asymptomatic infections ranging from 10% to 95% and a basic reproduction number ranging from 1.05 to 5.

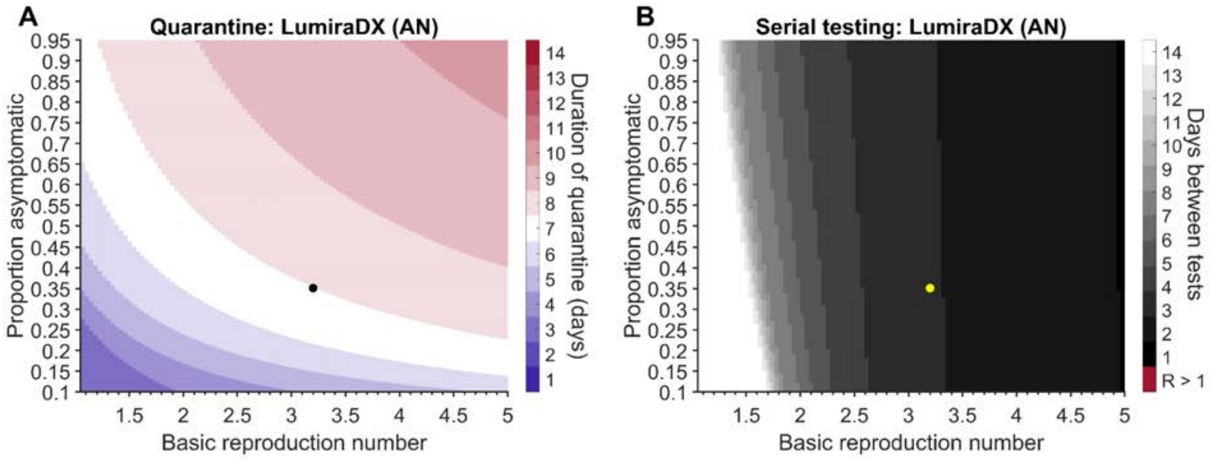


**Figure S40. The impact of asymptomatic infection and basic reproduction number on quarantine duration and frequency of serial testing for LumiraDx (AN).** Specifying an incubation period of 4.4 days and RT-PCR diagnostic sensitivity represented by a log-Normal distribution, (**A**) the quarantine duration for a RA test on exit (blue and red gradient) that has an equivalent or lower probability of post-quarantine transmission than the baseline scenario (35.1% asymptomatic infection and a basic reproduction number of 3.2) for a 7-day quarantine (black dot), and (**B**) the frequency of serial testing required to maintain the effective reproduction number below one (black and white gradient; baseline indicated by yellow dot) for asymptomatic infections ranging from 10% to 95% and a basic reproduction number ranging from 1.05 to 5.

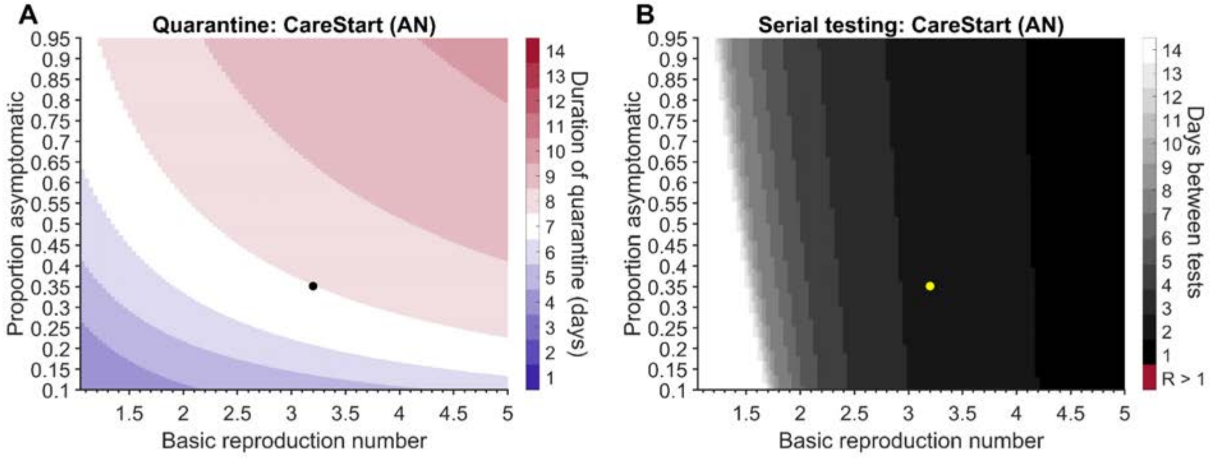


**Figure S41. The impact of asymptomatic infection and basic reproduction number on quarantine duration and frequency of serial testing for CareStart (AN).** Specifying an incubation period of 4.4 days and RT-PCR diagnostic sensitivity represented by a log-Normal distribution, (**A**) the quarantine duration for a RA test on exit (blue and red gradient) that has an equivalent or lower probability of post-quarantine transmission than the baseline scenario (35.1% asymptomatic infection and a basic reproduction number of 3.2) for a 7-day quarantine (black dot), and (**B**) the frequency of serial testing required to maintain the effective reproduction number below one (black and white gradient; baseline indicated by yellow dot) for asymptomatic infections ranging from 10% to 95% and a basic reproduction number ranging from 1.05 to 5.

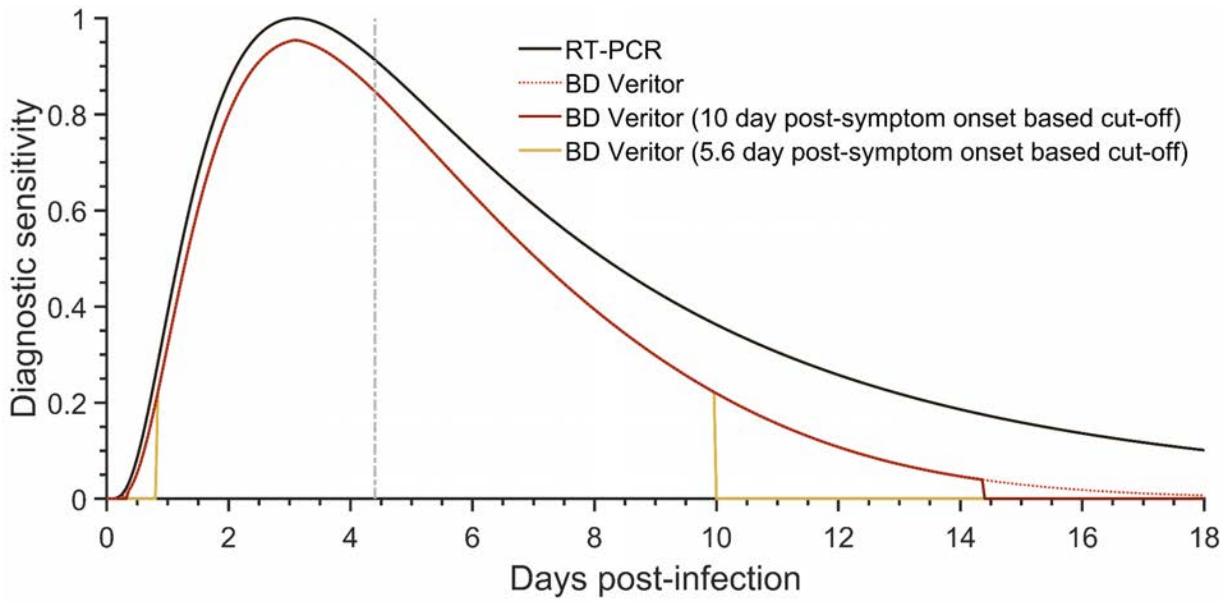


**Figure S42. Temporal diagnostic sensitivity of a rapid antigen test when specifying a threshold day for returning a positive test.** Specifying an incubation period of 4.4 days, the temporal diagnostic sensitivity of an RT-PCR test (black line), the BD Veritor rapid antigen test with no cut-off (red dashed line), the BD Veritor rapid antigen test with a threshold day specified by the level of infectivity 10 days after symptom onset (solid red line), and the BD Veritor rapid antigen test with a cut-off specified by the level of infectivity 5.6 days after symptom onset (solid yellow line).

### References cited in supplementary material


1. CDC. Overview of Testing for SARS-CoV-2 (COVID-19). https://www.cdc.gov/coronavirus/2019-ncov/hcp/testing-overview.html (2021).
2. Salvagno, G. L., Gianfilippi, G., Bragantini, D., Henry, B. M. & Lippi, G. Clinical assessment of the Roche SARS-CoV-2 rapid antigen test. *Acta Radiol. Diagn.* (2021) doi:10.1515/dx-2020-0154.
3. Guglielmi, G. Fast coronavirus tests: what they can and can't do. *Nature* **585**, 496–498 (2020).
4. Wells, C. R. *et al.* Optimal COVID-19 quarantine and testing strategies. *Nat. Commun.* **12**, 356 (2021).
5. Xiao, A. T. *et al.* Dynamic Profile of RT-PCR Findings from 301 COVID-19 Patients in Wuhan, China: A Descriptive Study. *SSRN Electronic Journal* doi:10.2139/ssrn.3548769.
6. CDC. Interim Guidance on Duration of Isolation and Precautions for Adults with COVID-19. https://www.cdc.gov/coronavirus/2019-ncov/hcp/duration-isolation.html (2021).
7. Cevik, M. *et al.* SARS-CoV-2, SARS-CoV, and MERS-CoV viral load dynamics, duration of viral shedding, and infectiousness: a systematic review and meta-analysis. *Lancet Microbe* **2**, e13–e22 (2021).
8. Hogan, C. A. *et al.* Large-Scale Testing of Asymptomatic Healthcare Personnel for Severe Acute Respiratory Syndrome Coronavirus 2 - Volume 27, Number 1—January 2021 - Emerging Infectious Diseases journal - CDC. doi:10.3201/eid2701.203892.
9. Hellewell, J. *et al.* Estimating the effectiveness of routine asymptomatic PCR testing at different frequencies for the detection of SARS-CoV-2 infections. *BMC Medicine* **19**, (2021).
10. Zhang, M. *et al.* Transmission Dynamics of an Outbreak of the COVID-19 Delta Variant B.1.617.2 — Guangdong Province, China, May–June 2021. *China CDC Weekly* vol. 3 584–586 (2021).
11. Lauer, S. A. *et al.* The Incubation Period of Coronavirus Disease 2019 (COVID-19) From Publicly Reported Confirmed Cases: Estimation and Application. *Ann. Intern. Med.* **172**, 577–582 (2020).
12. Ashcroft, P., Lehtinen, S., Angst, D. C., Low, N. & Bonhoeffer, S. Quantifying the impact of quarantine duration on COVID-19 transmission. *Elife* **10**, (2021).

13. Young, S. *et al.* Clinical Evaluation of BD Veritor SARS-CoV-2 Point-of-Care Test Performance Compared to PCR-Based Testing and versus the Sofia 2 SARS Antigen Point-of-Care Test. *Journal of Clinical Microbiology* **59**, e02338–20 (2021).
14. Abbott. *BinaxNOW COVID-19 Ag CARD*. https://www.fda.gov/media/141570/download.
15. Pollock, N. R. *et al.* Performance and Implementation Evaluation of the Abbott BinaxNOW Rapid Antigen Test in a High-throughput Drive-through Community Testing Site in Massachusetts. doi:10.1101/2021.01.09.21249499.
16. AccessBio. *CareStart COVID-19 Antigen test*. https://www.fda.gov/media/142919/download.
17. Pollock, N. R. *et al.* Performance and Operational Evaluation of the Access Bio CareStart Rapid Antigen Test in a High-throughput Drive-through Community Testing Site in Massachusetts. *Open Forum Infect Dis* (2021) doi:10.1093/ofid/ofab243.
18. *Celltrion DiaTrustTM COVID-19 Ag Rapid Test*. https://www.fda.gov/media/147694/download.
19. Luminostics. *Clip COVID Rapid Antigen Test*. https://www.fda.gov/media/144256/download.
20. Ellume. *Ellume COVID-19 Home Test*. https://www.fda.gov/media/144592/download.
21. DiaSorin. *Liaison SARS-CoV-2 Ag assay*. https://www.fda.gov/media/147311/download.
22. [No title]. https://www.fda.gov/media/141304/download.
23. [No title]. https://www.fda.gov/media/141304/download.
24. Qorvo Biotechnologies. *Omnia SARS-CoV-2 Antigen*. https://www.fda.gov/media/147578/download.
25. InBios. *SCoV-2 Ag Detect Rapid Test*. https://www.fda.gov/media/148353/download.
26. Quanterix. *Simoa SARS-CoV-2 N Protein Antigen Test*. https://www.fda.gov/media/144929/download.
27. Quidel. *Sofia SARS Antigen FIA*. https://www.fda.gov/media/137885/download.
28. Pray, I. W. Performance of an Antigen-Based Test for Asymptomatic and Symptomatic SARS-CoV-2 Testing at Two University Campuses — Wisconsin, September–October 2020. *MMWR Morb. Mortal. Wkly. Rep.* **69**, (2021).
29. Quidel. *Sofia 2 Flu + SARS Antigen*. https://www.fda.gov/media/142704/download.

30. LifeSign. *Status COVID-19/Flu*. https://www.fda.gov/media/145697/download.

31. VITROS. *VITROS Immunodiagnostic Products SARS-CoV-2 Antigen Reagent Pack*. https://www.fda.gov/media/145073/download.
